# A multimodal atlas of COVID-19 severity identifies hallmarks of dysregulated immunity

**DOI:** 10.1101/2025.09.19.25334095

**Authors:** Kamil Slowikowski, Pritha Sen, Christopher V Cosgriff, Jessica Tantivit, Tom Eisenhaure, Thomas J LaSalle, Kasidet Manakongtreecheep, Alice Tirard, Benjamin Y Arnold, Ana Pacheco-Navarro, Emily Yu-Ann Yang, Miguel Reyes, Anna Gonye, Irena Gushterova, Brian Russo, Maricarmen Rojas-Lopez, Nihaarika Sharma, Molly F Thomas, Tatyana Sharova, Dennie Frederick, Kendall Lavin-Parsons, Brendan Lilley, Brenna McKaig, Carl Lodenstein, Hargun Khanna, Kyle Kays, Nicole Charland, Neal Smith, Swetha Ramesh, Toni M Delorey, Devan Phillips, Liat Amir-Zilberstein, Eric M Brown, Maura Benson, Sung-Moo Park, Betsabeh K Tusi, Vladislav Pokatayev, Cody Hecht, Novalia Pishesha, Ann E Woolley, Lisa Cosimi, Orit Rozenblatt-Rosen, Lloyd Bod, Paul C Blainey, Aviv Regev, Jacques Deguine, Ramnik Xavier, Deborah Hung, Genevieve M Boland, Roby P Bhattacharyya, Paul J Utz, Marcia B Goldberg, Michael K Mansour, Michael R Filbin, Moshe Sade-Feldman, Nir Hacohen, Alexandra-Chloe Villani

## Abstract

The alpha-variant wave of the COVID-19 pandemic provided a unique opportunity to study, at single-cell resolution, how near-universal exposure to the same pathogen can lead to either effective or dysfunctional immune responses in humans. Although single-cell RNA-sequencing studies have characterized immune cellular features of COVID-19, they have not shown how tocilizumab treatment changes these features at single-cell resolution, or which features might persist into convalescence. In this study, we analyzed 2.5 million circulating immune cells from 428 patients across time points (840 PBMC samples), encompassing three contemporaneous SARS-CoV-2 cohorts: acutely infected patients across five WHO disease severity levels and three time points, patients from the first randomized control trial to study the efficacy of tocilizumab in the management of COVID-19, and convalescent patients three months after infection. We used linear modeling to integrate multiple data types – including single-cell RNA-seq, CITE-seq, TCR and BCR sequencing, viral load measurements, viral neutralization assays, detection of 75 autoantibodies, HLA genotype data, and serum proteomics covering 1,463 targets – to derive the most comprehensive view to-date of the biological features of COVID-19 disease severity. Our findings show that myeloid-derived suppressor cells (MDSCs) act as a key immunologic pivot point in severe COVID-19. Myeloid dysfunction, which is marked by impaired antigen presentation, drives a non-productive adaptive immune response, as reflected by reduced expression of B and T cell gene programs involved in antigen recognition, immune synapse formation, and cytotoxicity. Severe disease is also linked to autoantibodies targeting type I interferons, influenced by specific HLA-DQB1 allelic variants, and strongly correlated with serum IL-6 levels. Tocilizumab treatment eliminates *CLU*-expressing MDSCs and ISG-positive myeloid subsets, restores antigen presentation, and reactivates productive adaptive immunity. These changes align with improved clinical outcomes and better clinical laboratory measures, including reduced CRP. While many immunologic abnormalities in acute severe COVID-19 resolve during convalescence 3-months post-infection, we observed persistently high *ICOS* expression in regulatory T cells, potentially linking acute infection to chronic post-COVID syndromes. Overall, we define distinct innate and adaptive host immune responses associated with acute, IL-6–responsive, and convalescent SARS-CoV-2 infection. Our multimodal and high-dimensional dataset with curated clinical metadata provides a foundational and clinically relevant resource for modeling host immune response biology in health and disease.

## Introduction

While various biological facets of severe acute respiratory syndrome coronavirus 2 (SARS-CoV-2) infection have been investigated, the scale and focus of prior studies have made it difficult to establish clear and unifying mechanistic explanations underlying the range of dysfunctional immune responses observed in infection across the spectrum of disease severity. SARS-CoV-2 infection can be asymptomatic or cause a mild acute illness, but in some patients — more frequently among those infected with early strains before vaccines were available — it led to respiratory failure, multiorgan dysfunction, and death. Morbidity and mortality in COVID-19 is mediated by direct viral damage and host immune response^1–6^. When an impaired host immune response to infection leads to organ dysfunction, this condition is classified as sepsis. Understanding which biological determinants distinguish a host immune response that successfully clears an infection from one that leads to sepsis is a major challenge in medicine.

Despite significant research efforts, progress in understanding the underpinnings of sepsis has been hindered by the vast complexity of host and pathogen interactions shaping host immune response, which can vary in many ways. Recently, researchers have used advanced omics technologies to tackle this challenge by identifying more homogeneous groups of patients who may share similar host immune response dysfunctions^7,8^. Notably, these studies have uncovered common patterns of gene expression program activity across different infectious etiologies^9^. Additionally, single-cell genomics analyses have revealed novel immune cell states and gene programs associated with the development and severity of sepsis^10,11^.

During the first three months of the initial wave of the SARS-CoV-2 pandemic in Boston, MA, USA, prior to any significant viral evolution, widespread availability of anti-viral therapies, or vaccination, we conducted a longitudinal single-cell RNA sequencing (scRNA-seq) analysis of the peripheral blood immune response in 351 patients who presented to the emergency department with febrile respiratory illness. We integrated these data with clinical phenotype information, viral load measurements, serum proteomics, serum antibody levels, HLA genotype, and TCR/BCR repertoire characterization.

Our analyses revealed the hallmarks of host-immune response dysfunction across the full spectrum of SARS-CoV-2 disease severity as it evolves over time. We identify (1) the cellular and transcriptional features that distinguish an effective adaptive immune response from a dysfunctional one; (2) the interplay between antigen presentation, antigen receptor repertoire diversity, and host immune response effectiveness; (3) which cellular and transcriptional program changes in immune cells persist for months after infection; and (4) how IL-6-receptor inhibition through immunomodulatory tocilizumab therapy modulates immunity to reduce tissue damage and improve clinical outcomes in severe disease. We propose a model in which the initial failure to control the infection triggers a feedback loop, causing simultaneous immune overactivation and suppression. The immune overactivation is associated with tissue damage caused by failure to control the infection, and this suppresses the adaptive immune response that could help to control the infection. Our results suggest that disrupting this cycle is the key mechanism by which immunomodulatory therapy can benefit some patients with sepsis.

## Results

### Study Design

We assembled a single center cohort of patients that presented to the emergency department at the Massachusetts General Hospital (MGH) with a febrile respiratory illness during the initial alpha variant wave of the SARS-CoV-2 pandemic and collected blood samples from 351 patients across five acuity levels and at three time points, and subsequently adjudicated their clinical trajectory according to an established ordinal scale, in order to define immunological signatures of severity. We generated single-cell multiomics data from approximately 2.2 million peripheral blood mononuclear cells (PBMC) isolated from these patients (Figure 1 A). Of these patients, 302 had a positive PCR test from a nasal swab for SARS-CoV-2, and the remaining 49 patients were negative. We performed scRNA-seq, cellular indexing of transcriptomes and epitopes (CITE-seq), and TCR and BCR sequencing on 751 PBMC total samples from these patients collected across up to four time points, of which 705 samples passed QC and were included in downstream analysis (Methods; Figure S1). We integrated our cellular analysis with multiple additional datasets including serum autoantibody data, HLA genotyping inferred from bulk neutrophil transcriptomic sequencing^12^, as well as previously published serum proteomics^13^ and viral titer data^14^ measured in this same cohort.

**Figure 1.**
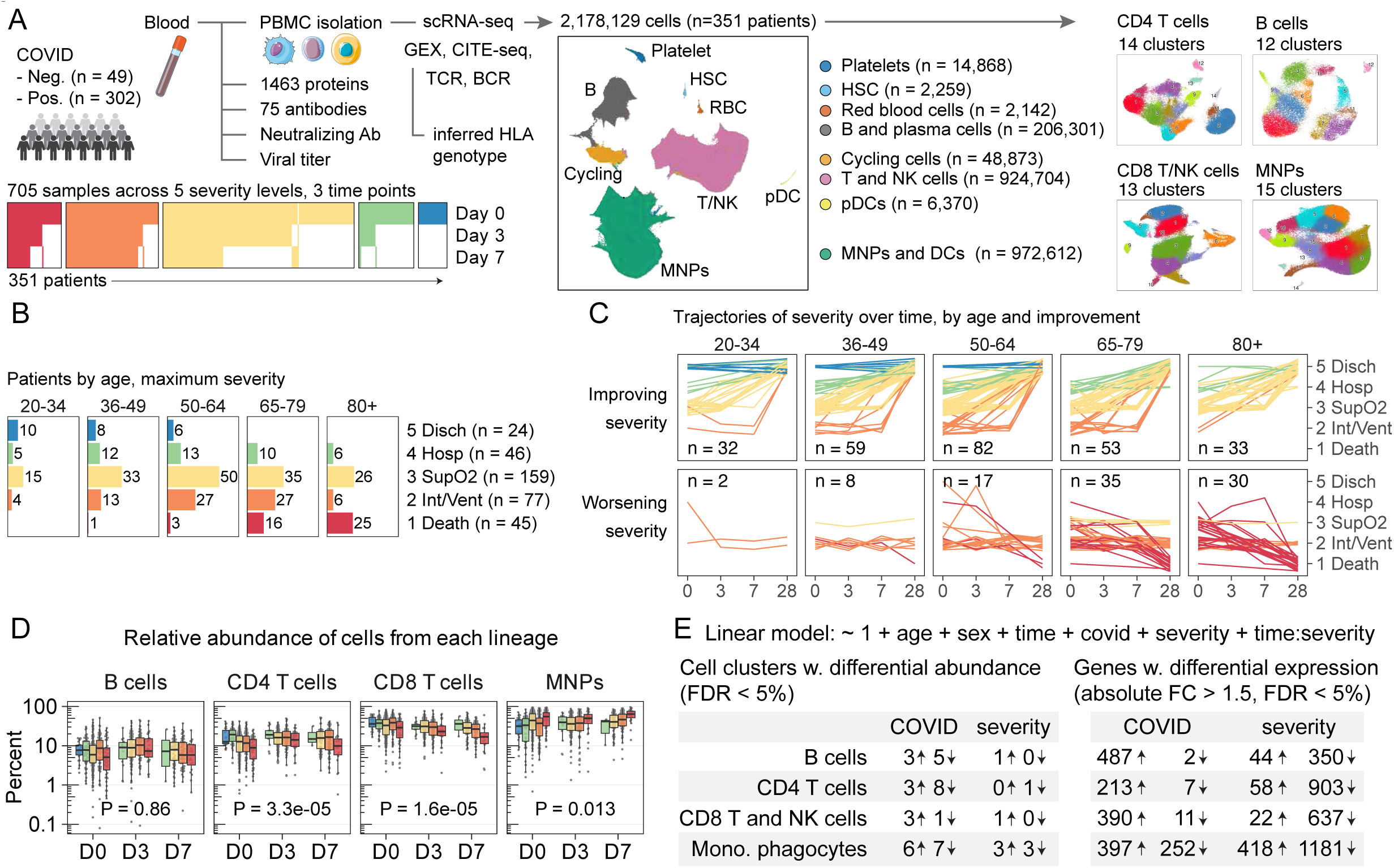
Overview of study design **A.** Overview of study design. 705 scRNA-seq samples from 351 patients across 3 time points. UMAP embedding of all cells where hue indicates cell lineage. UMAP embeddings of subclustering results for each cell lineage. **B.** Patients by age and maximum severity. **C.** Trajectories of COVID severity over time, by age and improvement. Hue indicates maximum COVID severity; y-axis indicates current severity with some jitter. Number of patients in each facet is shown. **D.** COVID-19 severity associations with relative abundance of B cells, CD4 T cells, CD8 T and NK cells, and Mononuclear Phagocytes (MNPs). Box plots show median and interquartile range, dots represent patient samples. Hue indicates COVID severity. x-axis indicates time point; y-axis indicates percent of cells from each sample in each cell lineage. P-values for the severity coefficient from multivariate linear regression models. **E.** Table summarizing the number of cell subsets in each lineage that are associated with infection or severity (left) and number of genes with differential expression (right).

Clinical features and biological samples were obtained and assessed on day 0 in the emergency department, and on days 3, 7, and 28 from presentation if the patient remained admitted to the hospital as an inpatient (Table S1; **Methods**). Patients had symptoms for a mean of 8.2 days before entering the emergency department (Figure S1 A). At each time point, clinical severity was manually annotated according to the initial WHO severity 5-point scale as follows: 1 Death (n=45), 2 Intubation/Ventilation (n=77), 3 Supplementary Oxygen (n=159), 4 Hospitalization (n=46), 5 Discharge (n=24)^13^. Consistent with previous reports^14,15^, age and viral load were higher in patients with worse acuity (Figure 1 B, C; Figure S1 B, D).

We also used a single-cell multi-omics approach to deeply characterize the host immune response in a separate cohort of patients after convalescent SARS-CoV-2 infection (defined as positive serology for IgG to the receptor binding domain (RBD) of the SARS-CoV-2 spike protein) Table S2, and another cohort of critically ill patients successfully treated with anti-IL-6 receptor therapy Table S3.

To distinguish the cellular subsets and transcriptional signals associated with different factors such as SARS-CoV-2 infection or COVID-19 disease severity, we fit linear models to the data. All analyses used the same linear model specification, where the feature of interest (e.g., gene expression) is estimated as a linear combination of age, sex, infection status, maximum severity level, time point, and the interaction between maximum severity and time (**Methods**). To integrate additional immunological measurements or modalities (e.g., serum protein levels, autoantibodies, HLA alleles), we included one additional variable in the model from the corresponding measurements. This approach provided the flexibility to extend the model with additional variables, enabling us to identify cellular subsets or transcriptional signatures associated with many different immunological measurements. We report our results at two levels of resolution: cell lineage (mononuclear phagocyte (MNP), CD4 T, CD8 T, B) and cell subset within each lineage (Figure 1 D, E).

Because our study included patients who tested negative for SARS-CoV-2 infection, we could distinguish which features were specific to SARS-CoV-2 infection or disease severity. Abundance analysis at the lineage level demonstrated myeloid expansion and T cell lymphopenia in association with worse severity (Figure 1 D), consistent with previous studies^16^. We found the greatest number of severity associations for cell subset abundance and gene expression in the MNP lineage (Figure 1 E), but all cell lineages had associations that characterize a dysfunctional immune state associated with severity.

### B cell subsets associated with COVID-19 severity

Within the B cell lineage, we identified 12 cell subsets: six naive populations (B-1, 2, 3, 6, 8, 9) expressing *FCER2*, *BACH2*, and *TCL1A*; five subsets with a memory or effector phenotype (B-4, 5, 7, 10, 11) expressing *CD27* and *TNFRSF13B*; and plasma cells (B-12) expressing *MZB1* (Figure 2 A, B, C) (Table S4). Infection with SARS-CoV-2 was associated with an increased abundance of the cell subsets expressing interferon-stimulated genes (ISGs) (e.g., *IFITM1*): naive B cells (B-3) and memory B cells (B-10) (Figure 2 C, D, E). Infection was also associated with the induction of over 500 genes in these two subsets (Figure S2 C). The elevated abundance of these ISG^hi^ subsets (B-3, 10) persisted over time in patients with worse severity (Figure 2 D, F), and decreased in proportion to the neutralization capacity of patient serum (Figure S3 A), but they were not associated with viral load (Figure S3 B). These results are consistent with the hypothesis that viral persistence in severe disease causes a persistent ISG^hi^ B cell state^17^.

**Figure 2.**
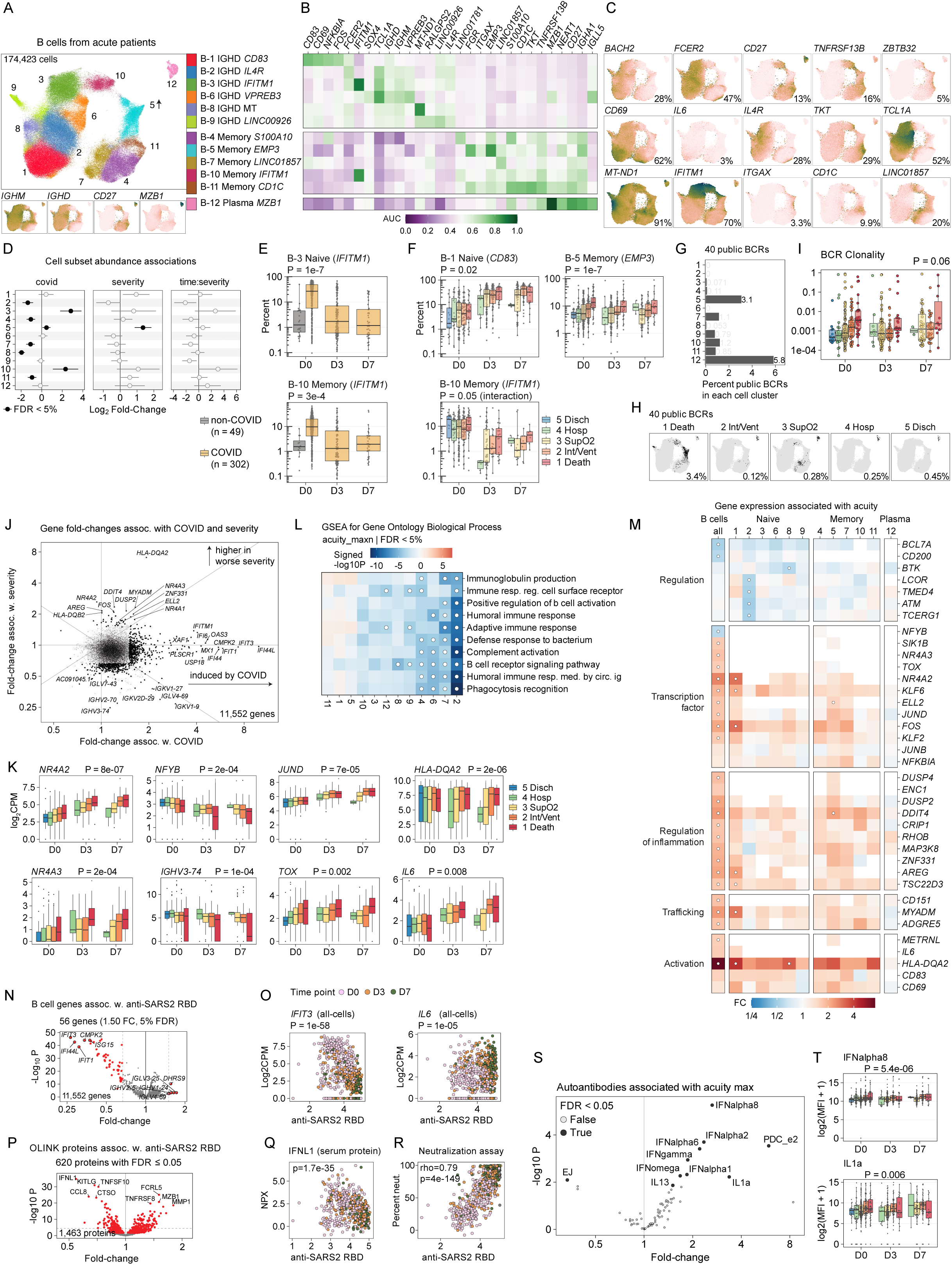
B cell subsets and genes associated with COVID-19 severity. **A.** UMAP embedding, hue indicates subset membership. *IGHM*, *IGHD*, *CD27*, *MZB1* are indicators of major types of B cells. **B.** Heatmap of marker genes selected for each cell subset, color indicates area under the receiver-operator curve (AUROC or AUC) for a one-versus-all comparison of cells in one subset versus other cells not in that subset. **C.** UMAP panels with log2CPM gene expression levels of selected marker genes for all B cells. **D.** Cell subset abundance associations for cell subsets (rows). Error bars indicate 95% CI for coefficients *covid*, *severity*, and *time:severity* from multivariate linear regression analysis. Black color indicates FDR < 5%. **E, F.** Cell subset abundance for subsets associated with COVID infection (**E**) or COVID severity (**F**). Dots represent patients, box plots indicate median and interquartile range, x-axis represents three time points (D0, D3, D7). Hue indicates COVID infection or severity. **G.** Bar plots showing percent of BCRs in each cell subset that are public. **H.** UMAP embeddings showing cells with public BCRs. Faceted by maximum COVID severity. **I.** Box plot of B cell BCR clonality in each sample. Hue indicates COVID severity. Dots represent patient samples, box plots indicate median and interquartile range. X-axis indicates time point, y-axis indicates clonality of BCRs in each sample. P-value indicates association with severity. **J.** Scatter plot of genes associated with COVID infection (x-axis) and COVID severity (y-axis). Axes represent log2 fold-change associations from multivariate linear regression. Dots represent genes, black color and size indicate absolute fold-change > 1.5 and FDR < 5%. **K.** B cell gene expression for genes associated with COVID severity. Dots represent patients, box plots indicate median and interquartile range, x-axis represents three time points (D0, D3, D7). Color indicates disease severity. **L.** Heatmap of gene set enrichment association results for COVID severity, gene sets (rows), cell subsets (columns). Hue indicates signed -log10 p-value. Dots indicate FDR < 5%. **M.** Heatmap of severity-associated fold-changes for selected genes (rows) and cell subsets (columns). Hue indicates fold-change. Dots indicate FDR < 5%. **N.** Volcano plot of B cell gene expression associations with antibody against SARS-CoV-2 RBD. Results from multivariate linear regression. Dots indicate genes, x-axis indicates fold-change (relative to a unit increase in anti-SARS-CoV-2-RBD on the MFI scale, see **Methods**), y-axis indicates negative log10 p-value. Red color indicates FDR < 5%. **O.** Gene expression of *IFIT3* and *IL6* (y-axis) versus antibody level of anti-SARS-CoV-2 RBD (x-axis). Hue indicates time point. P-value for antibody coefficient from multivariate linear regression. **P.** Volcano plot of OLINK serum protein associations with antibody against SARS-CoV-2 RBD. Results from multivariate linear regression. Dots indicate proteins, x-axis indicates fold-change (relative to a unit increase in anti-SARS-CoV-2-RBD on the MFI scale, see **Methods**), y-axis indicates negative log10 p-value. Red color indicates FDR < 5%. **Q.** Protein level of IFNL1 (y-axis) versus antibody against SARS-CoV-2 RBD (x-axis). Hue indicates time point. P-value for antibody coefficient from multivariate linear regression. **R.** Neutralization assay result (y-axis) versus antibody against SARS-CoV-2 RBD (x-axis). Hue indicates time point. Spearman correlation and p-value. **S.** 74 autoantibodies associated with SARS-CoV-2 severity, x-axis represents fold-change and y-axis represents negative log10 p-value. **T.** Two box plots show the measurements of antibodies against IFNalpha8 and IL1a, x-axis represents time points and y-axis represents the log2(MFI+1) measurement of the antibody. Hue indicates severity. Two-sided p-value for the coefficient in the linear model is shown.

Patients with more severe disease mainly had increased abundance of memory B cells (B-5) with *ITGAX*, *CD1C*, *EMP3* and naive B cells (B-1) with *CD83* (Figure 2 D,F; Table S5). The *ITGAX* (encoding CD11c) B-5 cells have previously been described as atypical memory B cells (also referred to as CD21^Lo^) found in association with chronic inflammatory processes, and have also been described as age-associated B cells^18–20^. Of note, these B-5 cells were associated with increasing age in our data (Figure S3 C).

COVID-19 disease severity was associated with higher expression of genes involved in regulation of inflammation and tissue repair such as *AREG*, *DDIT4*, *ZNF331*, and *MAP3K8* (Figure 2 J-M) within the B cell compartment across all cell subsets (Table S6). Severity was also associated with increased transcription of *JUND*, *TOX*, *NR4A2*, and *NR4A3*, encoding transcription factors implicated in regulation of effector function and cytokine production, and increased expression of *HLA-DQA2* and *IL6*, consistent with B cell activation (Figure 2 J, K, M). SARS-CoV-2 infection induced ISGs (*IFI44*, *IFI44L*, *IFIT1*, *IFIT3*) (Figure S2 A, D), especially in the atypical memory B cells (B-5) (Figure S2 F, G). However, ISGs were not associated with severity (Figure 2 J, Figure S2 B, E).

Examination of the BCR repertoire (Figure S3 E, F) revealed evidence of convergent selection with 5.8% of the plasma cell subset (B-12) expressing one of the 40 public clones (Figure 2 G; Figure S3 G; Table S7). Most public BCR clones localized to the atypical memory B cells (B-5) from patients who died (Figure 2 H). Further, increased BCR clonality was associated with worse severity (Figure 2 I) and age (Figure S3 D), but we did not identify specific BCR genes associated with acuity (Table S8). Since the abundance of atypical memory B cells (B-5) was not associated with neutralizing antibody levels (Figure S3 A; Table S9), these results may reflect a maladaptive B cell population that is more clonally expanded in severe COVID-19 disease. We also found that severity was associated with decreased expression of B cell receptor (BCR) genes (Figure 2 J, L, M; Figure S2 B, G; Figure S4 A; Table S10).

Additionally, we found repression of *CR2* and its encoded protein CD21 (the complement receptor 2, part of the B cell co-receptor) (Figure S5 A, B, C), which compounds the effect of lower transcription of BCR genes. We conclude that patients with more severe disease have increased abundance of atypical memory B cells and a B cell phenotype with lower expression of BCR machinery, which may impair the ability to recognize and respond to SARS-CoV-2 antigens.

### Aberrant humoral responses in COVID-19 severity

To further understand how changes in B cell subset abundance and gene expression are associated with the humoral response, we examined how our scRNA-seq findings correlate with serum levels of anti-SARS-CoV-2 antibodies, *in vitro* neutralization capacity, and serum proteins. Increased neutralizing capacity of a patient’s serum was associated with a proportional decrease of ISG transcription (e.g., *IFIT3*) (Figure 2 N,O) and serum IFNL1 levels (Figure 2 P,Q). In contrast, neutralization capacity was effectively a surrogate measure for the level of antibody targeting the receptor binding domain (RBD) of the SARS-CoV-2 spike protein (Figure 2 R). The level of antibody against SARS-CoV-2-RBD was associated with increased expression of genes associated with B cell activation and maturation (e.g., *IL6*) (Figure 2 O), and increased serum levels of protein markers of B cell maturation (e.g., MZB1, SDC1) (Figure 2 P).

Levels of antibodies against nucleocapsid (N) and RBD were negatively associated with age (Figure S6 A; Table S11), consistent with the increased abundance of atypical memory B cells (B-5) in older age and worse disease severity. These data suggest that the reduced production of antiviral antibodies may be one of the mechanisms by which advanced age contributes to increased disease severity.

Moreover, the integrative analysis of these multimodal observations supports the hypothesis that the optimality and clonality of the BCR repertoire influences how the host response progresses from the first phase of interferon production to the second phase of antiviral antibody production, and demonstrates how this progression is associated with changes in the transcriptional states of circulating cells. We conclude that patients with worse severity have plasma cells that fail to rapidly produce effective antiviral antibodies within the first two weeks of symptoms. The lack of antiviral antibodies impairs the productivity of the overall host immune response as reflected in the transcriptional states of other immune cell lineages.

### Autoantibodies are associated with COVID-19 severity

Autoantibodies against type I interferon have previously been associated with severe COVID-19^21,22^, and more recent studies have found additional autoantibodies suggesting a link between SARS-CoV-2 infection, autoantibody production, and the development of Long COVID or post-acute sequelae of COVID-19 (PASC) symptoms^23^. As such, we expanded our characterization of the humoral response by measuring 69 known autoantibodies (**Methods**; Figure S6; Table S11). Although we only detected auto-antibodies in a small subset of patients, we identified nine autoantibodies associated with disease severity: four types of interferon-alpha (1, 2, 6, 8), interferon gamma, interferon omega, IL1a, IL13, and pyruvate dehydrogenase E2 (PDC_e2) (Figure 2 S, T; Figure S6 A, B). As a positive control, samples positive for SARS-Cov-2 had significantly elevated levels of the anti-RBD and anti-N antibodies (Figure S6 C).

To reduce the dimensionality of the autoantibody data, we applied non-negative matrix factorization (NMF). A small subset of patients had a high score for NMF factor 7 (NMF-7), which has highest loading values for autoantibodies targeting six types of IFN-alpha (10, 8, 6, 2, 1, 7) and IFN omega (Figure S6 D). When we tested for NMF-7 association with cell subsets abundance and gene expression, we found the greatest number of associations within the myeloid lineage (Figure S6 E, F). The ISG^Hi^ cell subsets in the B cell, CD4 T cell, and myeloid lineages that were elevated after COVID infection were negatively associated with NMF-7, suggesting that autoantibodies against IFN alpha may inhibit the expansion of ISG^Hi^ cell states (Figure S6 F). This finding was further supported by the negative association between IFN alpha autoantibodies (NMF-7) and reduced expression of ISGs (e.g., *IFI27*, *IFI44*, *IFI44L*) within the myeloid lineage (Figure S6 G, H). Within the myeloid lineage, the pDC (MP-14) cells, which are the primary producers of IFNalpha^24^, had the greatest number of genes (over 300) repressed in association with IFN alpha autoantibodies, including *IFI27* and *SIGLEC1* (Figure S6 G, H, I; Table S12). Thus, although autoantibodies to type I interferon were found in a minority of patients, our results demonstrate that their presence affects the abundance of ISG^Hi^ cell subsets and the transcriptional phenotype of pDCs, both of which have critical functions in mediating anti-viral immunity.

### CD4 T cell subsets associated with COVID-19 severity

Within the CD4 T cell lineage, we identified 14 cell subsets composed of naive (CD4-1, 3), cytotoxic (CD4-5, 6), effector (CD4-5, 8, 9, 12), regulatory (CD4-13), and ISG^Hi^ (CD4-4, 10) phenotypes, as well as NK-like cells (CD4-2) and subsets that appear to be double-positive (CD4/CD8) cells (CD4-7, 11, 14) (Figure 3 A, B, C; Table S4).

**Figure 3.**
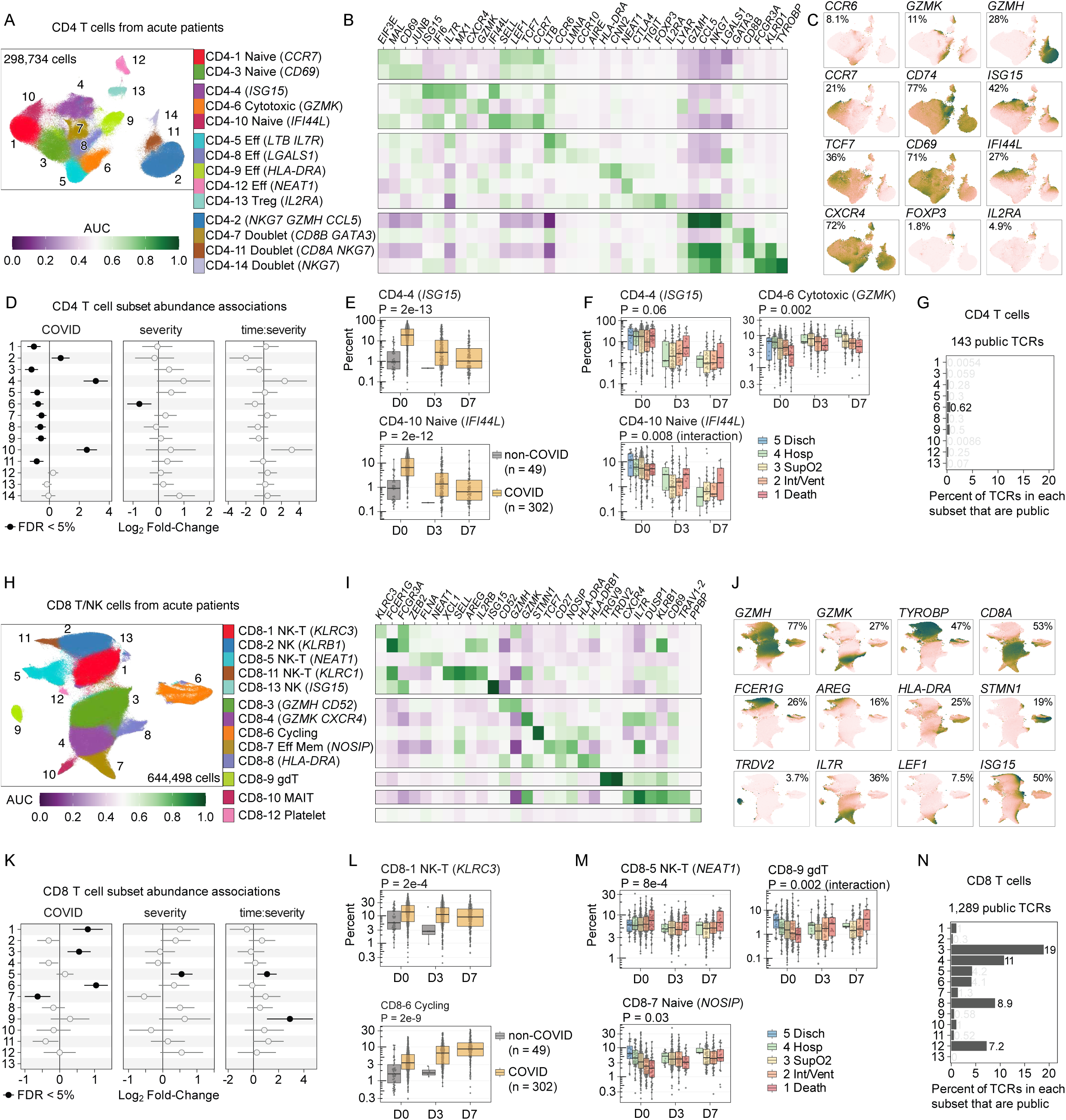
CD8 T/NK cell and CD4 T cell associations with COVID-19 severity. **A, H.** UMAP embedding of CD4 T cells (**A**) and CD8 T/NK cells (**H**), hue indicates subset membership. **B, I.** Heatmap of marker genes selected for each cell subset, hue indicates AUC. **C, J.** UMAP panels with log2CPM gene expression levels of selected marker genes. **D, K.** Cell subset abundance associations for CD4 T cell subsets (**D**) and CD8 T cell subsets (**K**). Error bars indicate 95% CI for coefficients *covid*, *severity*, and *time:severity* from multivariate linear regression analysis. Black circles indicate FDR < 5%. **E, F, L, M.** CD4 T cell and CD8 T/NK cell subset abundance for subsets associated with COVID infection (**E**, **L**) or COVID severity (**F**, **M**). Dots represent patients, box plots indicate median and interquartile range, x-axis represents three time points (D0, D3, D7). Hue indicates COVID infection or severity. **G, N.** Bar plot of the percent of TCRs from each cell subset that are public in CD4 or CD8 T cells.

SARS-CoV-2 infection was associated with significant increase of ISG^Hi^ cells (CD4-4, 10), increase in the cytotoxic CD4 T cell subset with *GZMH* and *CCL5* (CD4-2), and a corresponding relative decrease in most of the other cell subsets (CD4-1, 3, 5, 6, 7, 8, 9, 11) (Figure 3 D, E; Table S5). In patients with severe COVID-19, these ISG^Hi^ CD4 T subsets (CD4-4, 10) remained at a high abundance over seven days of hospitalization, but decreased over time in patients with less severe disease (Figure 3 F), consistent with our parallel observation in the B cell compartment. Worse disease severity was also associated with lower abundance of the cytotoxic *GZMK*^Hi^ *CXCR4*^Hi^ CD4-6 subset (Figure 3 D), which may be related to age-associated increase of CXCR4-mediated migration^25^. CXCR4 is a chemokine known for its role in T cell signaling and directing cells to bone marrow niches, and CXCR4 blockade improves survival in a mouse model of sepsis^26^.

### CD8 T cell subsets associated with COVID-19 severity

We identified five NK and NK T cell subsets (CD8-1, 2, 5, 11, 13) and seven CD8 T cell subsets (CD8-3, 4, 6, 7, 8, 9, 10), including gamma delta T cells (CD8-9) and MAIT cells (CD8-10) (Figure 3 H, I, J; Table S4). SARS-CoV-2 infection was associated with increased abundance of *KLRC3*^Hi^ NK cells (CD8-1), *GZMH*^Hi^ *CD52*^Hi^ cells (CD8-3), and *STMN1*^Hi^ cycling cells (CD8-6) (Figure 3 K, L). The proportion of *NOSIP*^Hi^ effector memory CD8 T cells (CD8-7) was decreased in SARS-CoV-2 infection (Figure 3 K) and also decreased in severe patients at time of admission (Figure 3 M; Table S5). Patients with severe COVID-19 had a relatively lower proportion of gamma delta T (gdT) cells (CD8-9) at admission (D0), but this cell subset subsequently increased over time (Figure 3 K, M), consistent with previous reports of a decrease in circulating proportions of gdT cells associated with disease severity and mortality in sepsis^27,28^. Greater abundance of *NEAT1*^Hi^ NK cells with (CD8-5) was associated with worse severity at all time points (Figure 3 K, M), consistent with previous reports^29,30^.

### Suboptimal TCR selection is a defining feature of COVID-19 severity

We examined the T cell receptor (TCR) repertoire in the CD4 T cell compartment (Figure S7 A, B), and found that patients with more severe disease or advanced age had higher TCR clonality, especially on the first day of hospitalization (Figure S7 C, D). We also found a lack of clonal expansion in the CD4 T cell compartment, where each CD4 T cell subset had less than 1% of cells with the 143 public TCRs we identified in our study (Figure 3 G; Figure S7 E, F; Table S13). Notably, public CD4 T cell TCRs were observed in very few donors (Figure S7 F, G). In contrast, we found 1,289 public TCRs in the CD8 T cells (Figure 3 N; Figure S8 A, B; Table S13), and the majority of these were observed in the cytotoxic subsets, with 19% of the GZMH^Hi^ (CD8-3) cells and 11% of the GZMK^Hi^ (CD8-4) cells containing public TCRs (Figure 3 N; Figure S8 C). Public CD8 TCR clones were observed in a greater number of donors than the public CD4 TCRs, and some specific V and J genes were associated with worse disease severity in both CD4 and CD8 T cell compartments (Figure S7 F, G, H, I, J, K; Figure S8 D, E, F, G, H). Therefore, we focused subsequent TCR repertoire analysis within CD8 T cells.

While severity was associated with higher clonality within the CD8 TCR repertoire at time of admission, patients with less severe disease had an increase in CD8 TCR clonality over the first week of hospitalization (Figure 4 A). Therefore, we hypothesize that individuals with a more limited initial repertoire may be predisposed to the selection of suboptimal clones over time, whereas patients with a more diverse initial repertoire may be more likely to expand effective clones over time. Since age was significantly associated with higher clonality at all time points (Figure 4 A), these observations further reflect that age-dependent lack of T cell diversity may predispose patients toward mounting an initially suboptimal adaptive immune response.

**Figure 4.**
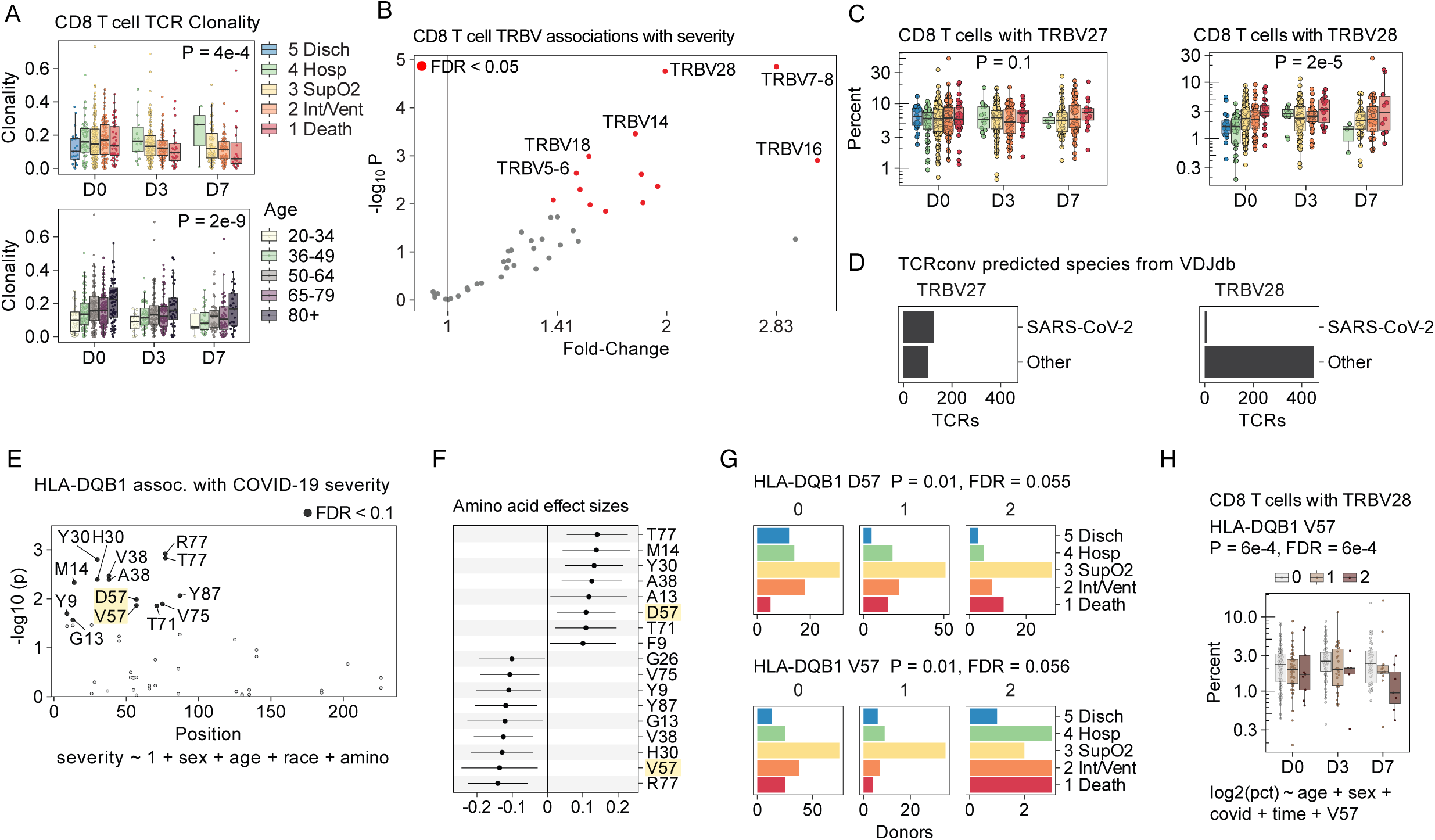
Association of COVID severity with CD8 T cell TCRs and HLA-DQB1 amino acid positions. *HLA and TCR associations with COVID-19 severity*. **A.** Box plot of CD8 T cell TCR clonality by COVID severity (top) and age (bottom). Dots represent patient samples, box plots indicate median and interquartile range, x-axis represents three time points (D0, D3, D7). Hue indicates COVID severity or age. P-value from multivariate linear regression. **B.** Volcano plot of TRBV associations with COVID severity. x-axis represents fold-change (corresponds to a 1-unit increase the percentage of CD8 T cells that have a given TRBV gene). y-axis represents negative log10 p-value from multivariate linear regression. Red color indicates FDR < 5%. **C.** Box plots of CD8 T cell abundance with TRBV27 (left) or TRBV28 (right). Dots represent patient samples, box plots indicate median and interquartile range, x-axis represents three time points (D0, D3, D7). Hue indicates COVID severity. P-value from multivariate linear regression. **D.** Bar plots of TCRconv predicted species for TCRs with TRBV27 (left) or TRBV28 (right). **E.** Amino acid position associations with COVID severity for HLA-DQB1. x-axis indicates position along the gene. y-axis indicates negative log10 p-value from multivariate linear regression. Black color indicates FDR < 5%. **F.** Coefficients from the multivariate linear regression for the effect size of each amino acid position on COVID severity. Amino acid positions D57 and V57 are highlighted. Error bars indicate 95% CI. **G.** Bar plots of the number of patients with each genotype. Facets indicate genotype (0, 1, 2) for amino acid position D57 (top) and V57 (bottom). x-axis indicates number of patients. Hue indicates COVID severity. **H.** Box plot of abundance of CD8 T cells with TRBV28, x-axis indicates time point and hue indicates genotype of HLA-DQB1 V57 (0, 1, 2). P-value from multivariate linear regression.

To investigate this TCR selection hypothesis further, we assessed whether the abundance of CD8 T cell subsets with different TCR V and J genes was associated with severity. We found that many V genes of the TCR beta chain (e.g., *TRBV28*, *TRBV7-8*, *TRBV5-6*) were more abundant in CD8 T cells from patients with more severe disease (Figure 4 B; Table S14). Previous *in vitro* peptide stimulation assays reported expansion of T cells expressing *TRBV27* in response to the peptide present in SARS-CoV-1 and SARS-CoV-2, but *TRBV28* in response to a nearly identical peptide from common coronavirus^31^. In our patients infected with SARS-CoV-2, we observed an expansion of *TRBV28* expressing CD8 T cells in association with more severe disease, but we found no association for *TRBV27* (Figure 4 C). Therefore, we hypothesized that expansion of CD8 T cells with *TRBV28* in more severely ill patients might indicate TCR repertoire bias toward TCRs with a lower affinity for SARS-CoV-2 peptides.

To test this hypothesis, we trained a previously reported deep learning model (TCRconv; **Methods**) that predicts epitope binding to TCR sequences on the publicly available VDJ database^32,33^, and generated epitope affinity predictions for the CD8 TCRs in our cohort (Figure S8 I). We found that many *TRBV27* TCRs were predicted to bind epitopes from SARS-CoV-2 (primarily SPR from the SARS-CoV-2 nucleoprotein), but nearly none of the *TRBV28* TCRs were predicted to bind known SARS-CoV-2 epitopes (Figure 4 D), consistent with the published *in vitro* results^31^. These data suggest that some patients with severe COVID-19 may have expanded CD8 T cells with TCRs that have low specificity for SARS-CoV-2 epitopes.

### HLA alleles are a genetic risk factor for disease severity

To understand if the molecular structure of the antigen presentation complex is associated with more severe disease, we tested for an association between disease severity and each amino acid position in each human leukocyte antigen (HLA) gene (Figure S9 A; Table S15; **Methods**). These analyses revealed several *HLA-DQB1* amino acid positions associated with disease severity (Figure 4 E), with a subset of positions having opposing effects depending on the amino acid residue. Aspartic acid (D) at position 57 was associated with worse disease severity and higher viral load (Figure S9 B, D; Table S16), while valine (V) at position 57 was protective against severe disease (Figure 4 F, G). Serine (S) at position 57 was associated with increased neutralization (Figure S9 C, E; Table S17). Notably, DQB1 D57 is strongly protective against type 1 diabetes^34^ but nearly ubiquitous in patients with narcolepsy^35^.

Next, we investigated how these COVID-19 severity-associated amino acids in the antigen presentation proteins might influence TCR selection. We tested if the abundance of CD8 T cells expressing each TCR beta chain V (TRBV) gene is associated with each amino acid position (**Methods**). The protective DQB1 V57 allele was associated with a lower proportion of CD8 T cells with severity-associated TRBV genes including *TRBV28* (Figure 4 H), *TRBV7-8* and *TRBV5-6* (Figure S9 F; Table S18). Since class II HLA presentation to CD4 T cells has been shown to be important for calibrating the CD8 T cell response^36^, our results suggest that *HLA-DQB1* genotype and CD4 T cell helper cells may drive the selection of suboptimal CD8 T cell TCRs and therefore, COVID-19 severity.

### Severity-associated gene expression changes in T lymphocytes

In both the CD4 and CD8/NK T cell lineages, SARS-CoV-2 infection induced transcription of ISGs (e.g., *IFI44L*, *IFIT3*, *OAS1*), but, as with the B cell lineage, expression of these genes was not associated with disease severity (Figure 5 A, B; Figure S10 A, B, D, E; Figure S11 A, B, D, E). In both the CD8/NK and CD4 T cell lineages, we also found hundreds of genes repressed in severe patients, including TCR genes (e.g., *TRBV5-5*) and genes responsible for modulating the immune synapse and TCR activation (*SPACA9*, *CETN2*, *ANXA1*) (Figure 5 A, B, C, D). Gene set enrichment analysis confirmed this observation, demonstrating that COVID-19 severity is associated with repression of genes in the T cell receptor complex in CD8/NK and CD4 T cells (Figure 5 E; Figure S4 A; Table S10). In the CITE-seq data, we found that CD101 was negatively associated with severity in CD4 T cells (Figure S5 D, E), and CD27 and CD28 were negatively associated with severity in CD8 T cells (Figure S5 F, G). These results are consistent with the downregulation of TCR signaling in severe patients.

**Figure 5.**
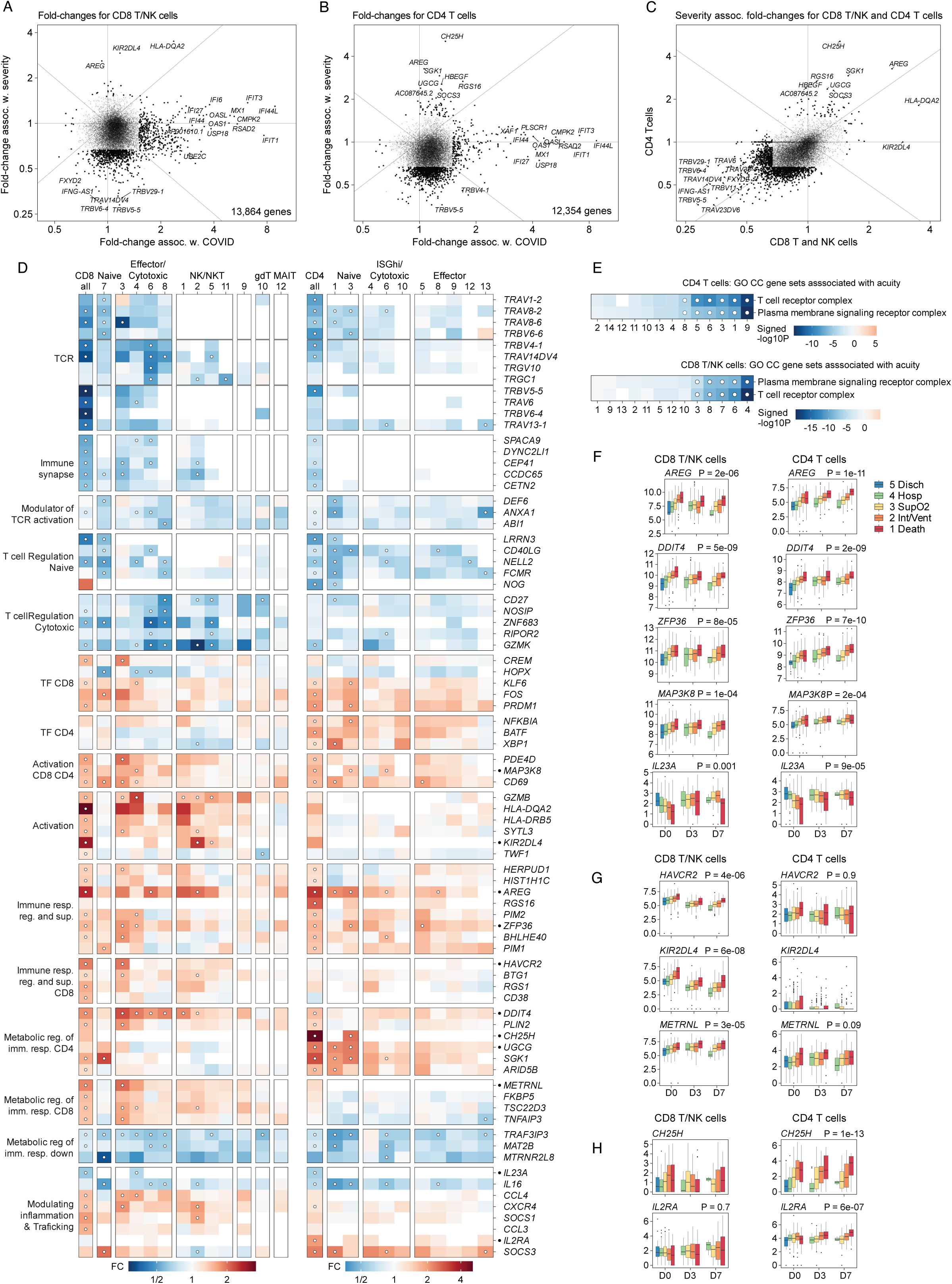
CD8 T/NK and CD4 T cell gene expression associations with COVID-19 severity. **A, B.** Scatter plot of genes associated with COVID infection (x-axis) and COVID severity (y-axis) for CD8 T/NK cells (**A**) and CD4 T cells (**B**). Axes represent fold-change associations from multivariate linear regression. Dots represent genes, black color and size indicate absolute fold-change > 1.5 and FDR < 5%. **C.** Scatter plot of genes comparing severity associations between CD8 T/NK cells (x-axis) and CD4 T cells (y-axis). Axes represent fold-change associations from multivariate linear regression. Dots represent genes, black color and size indicate absolute fold-change > 1.5 and FDR < 5%. **D.** Heatmap of fold-changes for selected genes in multiple categories (rows) and cell subsets (columns). Left panel shows CD8 T cells, right panel shows CD4 T cells. Hue indicates fold-change. Dots indicate FDR < 5%. **E.** Heatmap of signed negative log10 P-values for gene set enrichment results showing association between severity fold-changes in each cell subset. Gene Ontology Cellular Compartment gene sets are shown for CD4 T cells (top panel) and CD8 T cells (bottom panel). Dots indicate FDR < 5%. **F, G, H.** CD8 T cell (left column) and CD4 T cell (right column) gene expression for genes associated with COVID severity. **F** shows genes associated with both CD8 and CD4 T cells. **G** shows genes associated with CD8 T cells, but not CD4 T cells. **H** shows genes associated with CD4 T cells, but not CD8 T cells. Dots represent patients, box plots indicate median and interquartile range, x-axis represents three time points (D0, D3, D7). Hue indicates COVID severity. P-value from multivariate linear regression.

Although severity-associated transcriptional repression was observed across all of the T cell subsets, it was most pronounced in the NK-like CD4 cell subset (CD4-2) (Figure S11 C) and the *STMN1*^Hi^ cycling CD8 subset (CD8-6) (Figure S10 C). However, many genes across both lineages had increased transcription in association with disease severity, including *AREG*, *DDIT4*, *ZFP35*, *MAP3K8*, and *IL23A* (Figure 5 D, F).

Moreover, despite decreased expression of TCR related genes, several transcription factors and genes associated with T cell activation (e.g., *KLF6*, *FOS*, *PRDM1*, *BATF*, *XBP1*, *CD69*) were upregulated in CD4 and CD8 T cells. This increase in T cell activation markers was not accompanied by an increase in T cell proliferation signals (i.e., no severity association with the cycling CD8-6 subset, Figure 3 K). Moreover, while cell cycling gene sets were upregulated after infection, they were downregulated in worse severity (Figure S10 F, G). These results suggest TCR-independent, non-specific, and potentially defective T cell activation.

We also identified severity-associated genes that were specific to one T cell lineage. Within the CD8 lineage, we identified *HAVCR2* and *METRNL* (Figure 5 D, G; Table S6), which encode the inhibitory proteins TIM-3^37^ and Meteorin-like^38^ respectively. We also observed altered cytotoxicity within the CD8 lineage in association with increased disease severity: CD8 T and NK cells repressed *GZMK* and induced *GZMB* (Figure 5 D), consistent with findings in other forms of sepsis^39^. In contrast, the CD4 T lineage, but not the CD8 T lineage, had severity-associated upregulation of *CH25H*, which encodes a metabolic switch that constrains activation in response to interleukin-27 (IL-27) signaling, and *IL2RA*, which encodes a subunit of the IL-2 receptor complex and is implicated in T cell activation and proliferation, and maintenance of immune tolerance (Figure 5 D, H).

We also identified lineage-specific transcriptional changes with potential relevance to viral entry. *KIR2DL4* was higher in NK cells from severe patients, and encodes an inhibitory killer immunoglobulin-like receptor that binds heparan sulfate^40^, a host cell surface glycosaminoglycan that facilitates SARS-CoV-2 entry^41^. *KIR2DL4* is the only member of the KIR family known to bind FcepsilonRI-gamma^42^, which negatively regulates pDC function^43^, suggesting that transcriptional modulation of *KIR2DL4* in NK cells may contribute to altered dendritic cell regulation in the context of severe COVID-19. *UGCG* in CD4 T cells, but not CD8 T cells, was associated with severity (Figure 5 D), and encodes an enzyme responsible for the synthesis of glucosylceramide, a sphingolipid reported to promote viral entry^44^. Additionally, within the *ISG15*^Hi^ CD4 subset (CD4-4), we found that the gene set previously implicated as essential for viral entry was enriched with severity-associated genes (Figure S11 F, G)^45^, but we did not find this association in CD8 T cells (Table S10).

In summary, these transcriptional changes in more severe COVID-19 suggest an activated T lymphocyte phenotype marked by the repression of genes involved in antigen recognition, metabolic activity, immune synapse formation, and cytotoxicity, and simultaneous induction of genes involved in responding to tissue injury and repair. This shift toward T cell transcriptional states involved in tissue injury and repair, and away from viral clearance, may be a necessary response given the extent to which patients with severe disease have been found to have profound end-organ damage and tissue injury^13,46^.

### Sepsis-associated monocytes are a cornerstone of COVID-19 severity

We next characterized the role of the myeloid cell compartment in driving the dysregulated host response in severe SARS-CoV-2 infection. We identified 14 distinct mononuclear phagocytes (MNP) subsets (Figure 6 A, B, C), including nine classical CD14 monocyte subsets (MP-1-8, 15), three non-classical CD16 monocyte subsets (MP-9, 10, 12), cDCs (MP-11), pDCs (MP-14), and MNP-T cell doublets (MP-13) (Table S4). These myeloid subsets fall into four states: a progenitor state (MP-1, 4, 7 with *AHNAK*, *VIM*, *CD99*), a previously reported sepsis-associated state (MP-3 with *ALOX5AP*, *RETN*, *MCEMP1*, *PADI4*, *CLU*, *SELL*)^10^, an ISG^Hi^ state (MP-2, 5, 6, 10), and ISG^Lo^ effector state (MP-8, 9, 12) spanning both CD14 and CD16 MNPs.

**Figure 6.**
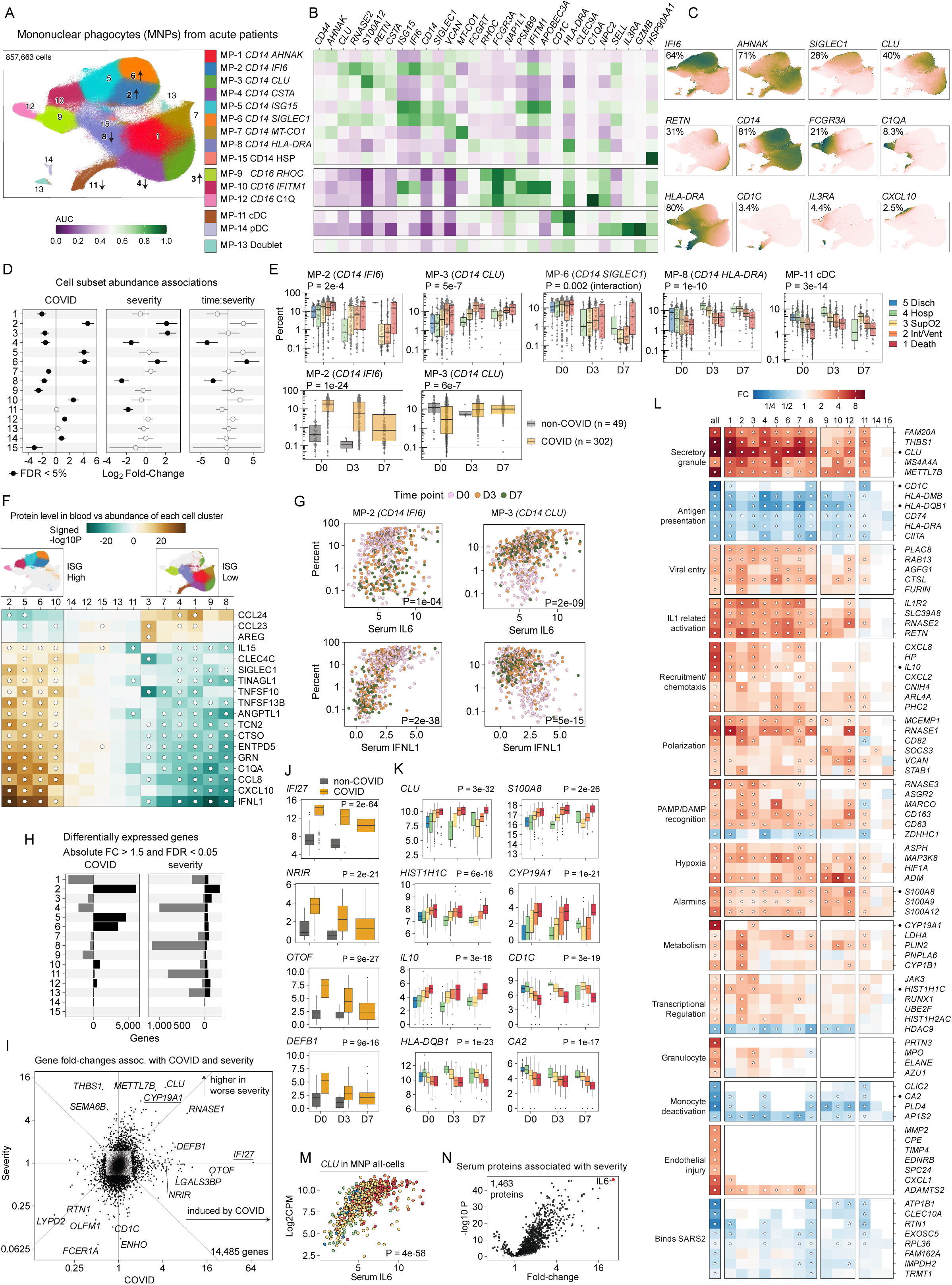
Mononuclear phagocyte cell subsets associated with COVID-19 severity. **A.** UMAP embedding of 857,663 MNP cells, color indicates membership to 15 subsets. Arrows indicate relative increased or decreased abundance association with COVID-19 severity. **B.** Heatmap of marker genes for each cell subset, hue indicates AUC. **C.** UMAP panels with log2CPM gene expression levels for selected marker genes. **D.** Cell subset abundance associations. Error bars indicate 95% CI for coefficients *covid* and *severity* and *time:severity* from multivariate linear regression analysis. Black circles indicate FDR < 5%. **E.** MNP cell subset abundance for subsets associated with COVID severity: MP-2, MP-3, MP-6, MP-8, MP-11. Dots represent patients, box plots indicate median and interquartile range, x-axis represents three time points (D0, D3, D7). Hue indicates COVID severity (top panels) or COVID infection status (bottom panels). **F.** Heatmap of OLINK proteins (rows) associated with abundance of each MNP cell subset (columns). Color indicates signed -log10 p-value (two-sided) for the protein coefficient from multivariate linear regression. UMAP embeddings show ISG High subsets (2,5,6,10) and ISG Low subsets (3,7,4,1,9,8). **G.** Scatter plots of blood protein (x-axis) versus MNP cell subset abundance (y-axis). Dots indicate patient samples, y-axis indicates log10 percent abundance of cell subset, x-axis indicates protein level in blood, hue indicates time point (D0, D3, D7). Two-sided p-value for the protein coefficient from multivariate linear regression. **H.** Bar plot of the number of differentially expressed genes for each cell subset, associated with COVID infection (left) or COVID severity (right). **I.** Scatter plot of genes associated with COVID infection (x-axis) and COVID severity (y-axis). Axes represent fold-change associations from multivariate linear regression. Dots represent genes, black color and size indicate absolute fold-change > 1.5 and FDR < 5%. **J, K.** Gene expression for genes associated with COVID infection (**J**: *IFI27*, *NRIR*, *OTOF*, *DEFB1*) or severity (**K**: *CLU*, *S100A8*, *HIST1H1C*, *CYP19A1*, *IL10*, *CD1C*, *HLA-DQB1*, *CA2*). Dots represent patients, box plots indicate median and interquartile range, x-axis represents time point. Hue indicates COVID infection (**J**) or disease severity (**K**). **L.** Heatmap of gene fold-changes for each all MNP cells (first column) and each MNP cell subset (columns) for genes grouped into categories. Hue indicates fold-change, dot indicates FDR < 5%. **M.** Scatter plot of IL-6 blood protein (x-axis) versus *CLU* gene expression in MNP cells (y-axis). Dots indicate patient samples, y-axis indicates log2 CPM gene expression, x-axis indicates protein level in blood, hue indicates COVID severity. Two-sided p-value for the protein coefficient from multivariate linear regression. **N.** Volcano plot for OLINK proteins associated with maximum COVID severity. Multivariate linear regression, two-sided p-value for the coefficient for severity. X-axis represents fold-change for each protein, and y-axis represents -log10 p-value.

SARS-CoV-2 infection is associated with increased abundance of *CD14* ISG^Hi^ subsets (MP-2, 5, 6), *CD16* ISG^Hi^ (MP-10), *CD16 C1QA*^Hi^ (MP-12), and pDC cells (MP-14) (Figure 6 D, E; Table S5), consistent with previous reports^1,4^. Patients with more severe disease had decreased relative abundance of the *HLA-DRA*^Hi^ subset (MP-8), cDCs (MP-11), and *CD14 CSTA*^Hi^ subset (MP-4) (Figure 6 D, E), also consistent with prior studies in COVID-19 and sepsis more broadly^1,2,4,21,47^. In contrast, patients with more severe disease had an increased relative abundance of two *CD14* ISG^Hi^ subsets (MP-2, MP-6), as well as the *CD14* ISG^Lo^ subset (MP-3) (Figure 6 D, E).

Notably, we identify MP-3 as a previously reported CD14 HLA-DR^Lo^ sepsis-associated monocyte state originally referred to as CD14 monocyte state 1 (CD14 MS1) that was identified in patients with bacterial sepsis^10^, which has also been described as myeloid-derived suppressor cells (MDSCs)^2,48,49^. The gene program defining this cell state (*CLU*, *RETN, ALOX5AP,* HLA-DR^Lo^) (Figure 6 B, C) has been reported in other studies of patients with COVID-19, and experimental work has both characterized them as immunosuppressive, and demonstrated that plasma from patients with either bacterial sepsis or severe COVID-19 can lead to their induction^2,4^. We also found the severity-associated ISG^Hi^ MP-2 subset to be transcriptionally similar to the MS-1 MP-3 subset, reflecting an ISG^Hi^ variant of the sepsis-associated monocyte state (Figure S12 A).

Next, we tested for association between the abundance of MNP subsets and serum proteins. We identified two distinct sets of serum proteins associated with the ISG^Hi^ or the ISG^Lo^ MNP cell subsets (Table S19). Abundance of ISG^Hi^ MNP subsets was associated with proteins involved in the early phase of acute response to viral infection (e.g., IFNL1, CXCL10, CCL8, IL15), whereas ISG^Lo^ subsets were associated with a distinct set of proteins, including monocyte chemoattractants associated with chronic inflammatory conditions, myelopoiesis and an immunosuppressive tumor microenvironment (e.g., AREG, CCL23, CCL24) (Figure 6 F)^50^. This bimodal pattern suggested that the ISG^Lo^ subsets, including the sepsis-associated MP-3 subset, are associated with serum markers of tissue damage and persistent inflammation, not to the markers of acute anti-viral response.

We examined myeloid cell subset associations with the serum level of IL-6, as previous reports have demonstrated correlation between plasma concentrations of IL-6 and the sepsis-associated monocyte state^2^. Concordant with these prior findings, we found that the relative abundance of the MP-3 subset increased in proportion to serum IL-6 levels, and observed that this relationship persisted over the seven day time course of hospitalization (Figure 6 G; Table S19). Although the transcriptionally similar MP-2 subset was also associated with serum IL-6 levels, its abundance decreased over time in strong association with the level of serum interferon (e.g., IFNL1) (Figure 6 G). These longitudinal data demonstrate the shift from the early interferon phase to a later IL-6 phase of disease associated with the expansion of specific MNP subsets. These data are also consistent with a model in which IL-6 promotes the development of the sepsis-associated subset (MP-3), and demonstrate that persistent high levels of IL-6 are present in patients with ongoing inflammation, tissue injury, and more severe illness.

### Myeloid cell gene expression associations with severity

COVID-19 severity was associated with a different pattern of gene expression than infection (Figure 6 H, I; Figure S13 A, B). Infection with SARS-CoV-2 induced gene expression in ISG^Hi^ myeloid subsets (MP-2, 5, 6, 10), *CD16 C1Q* subset (MP-12), and cDC (MP-11), but repressed gene expression in the other subsets (Figure 6 H, I; Table S6). Gene set enrichment analysis of infection-associated genes shows increased expression of genes in the RIG-I-like and Toll-like receptor signaling pathways across all MNP subsets (Figure S4 B; Table S10). Within some MNP subsets, infection is associated with the innate immune response and interferon signaling responses (Figure S13 C).

Most genes induced by infection, such as ISGs (e.g., *IFI27*, *OTOF*, *DEFB1*) and ISG regulators (e.g., *NRIR*) (Figure 6 J) were not associated with disease severity (Figure 6 I), with the exception of a few genes like *RNASE1* that were associated with both.

Severity was associated with repression of genes involved in antigen presentation (*CD1C*, *HLA-DRA*, *HLA-DQB1*, *CA2*) (Figure 6 L), specifically in *CD14* subset (MP-8), *CD14* progenitor-like (MP-4), and cDCs (MP-11) (Figure 6 L; Figure S13 D), which are the same subsets that are less abundant in severe patients (Figure 6 D). These results are complementary to the repression of TCR genes in the T cell compartment (Figure 5 D, E) — repressed gene expression on both sides of the immune synapse was associated with worse disease severity. This shows that repressed gene expression across both sides of the immune synapse is associated with worse disease severity.

Among the genes upregulated with increasing disease severity, we identified genes involved in immune activation and its negative feedback (*IL10*), as well as alarmins (*S100A8*, *S100A9*, *S100A12*), tissue damage and repair (*CLU*, *MMP2*), and transcriptional regulation (*HIST1H1C*, *JAK3*) (Figure 6 K, L; Figure S13 B). Increased MNP expression of *CYP19A1* was also associated with worse severity, consistent with a previous report of this aromatase-encoding gene mediating worse COVID-19 severity in males^51^.

Some severity-associated genes were increased or decreased among all MNP subsets (Figure 6 L). For example, all myeloid subsets had increased severity-associated expression of genes implicated in the secretory granule (*MS4A4A, METTL7B, FAM20A*, *THBS1*, *CLU*), which appeared in our gene set enrichment analyses (Figure S13 D).

Other key biological processes that were upregulated across all MNP subsets included viral entry (*PLAC8*, *FURIN*, *CTSL*, *AGFG1*, *RAB13*), IL-1 related pathway genes (*IL1R2*, *RETN*, *SLC39A8*), mononuclear phagocyte polarization (*VCAN*, *SOCS3*), scavenger receptors and PAMP/DAMP signaling (*CD163*, *MARCO*, *STAB1*), tissue hypoxia (*MAP3K8, ADM*, *HIF1A),* alarmins (*S100A8*, *S100A9*, *S100A12*) and endothelial injury (*ADAMTS2*) (Figure 6 L, Figure S13 D). In parallel, we found that all myeloid subsets had severity-associated downregulation of genes implicated in monocyte deactivation (*PLD4*, *AP1S2*), histone deacetylase genes (*HDAC9*, *HDAC2*), and genes that encode proteins known to bind SARS-CoV-2 (*ATP1B1*, *CLEC10A*, *RTN1*) (Figure 6 L, Figure S13 D).

We note that most severity-induced genes were in the sepsis-associated subsets (MP-2, MP-3) (Figure 6 H), and the induced genes unique to these subsets were related to metabolism (*LDHA*, *PLIN2*, *FABP5*, *CYP1B1*), transcriptional regulation (*HIST3H2A*, *HIST1H2AC*, *RUNX1*) and cytokine signaling (*JAK3*) (Figure 6 L). Considering the association between serum IL-6 levels and the abundance of the sepsis-associated subsets (MP-2, MP-3) (Figure 6 G), this severity-associated *JAK3* expression in both subsets is consistent with published reports demonstrating that IL-6 mediated JAK3-STAT3 activation results in the repression of HLA-DR gene expression that is characteristic of the sepsis-associated monocyte state^52^.

Among myeloid-expressed genes associated with serum IL-6 level, *CLU* had the strongest association (Figure S12 B; Table S20), and this gene was associated with severity in all myeloid subsets. *CLU* encodes clusterin, which has been reported as crucial for sepsis survival with observational and experimental data demonstrating it to be an anti-inflammatory, histone-neutralizing protein upregulated in response to extensive cell death^53^. These data suggest that IL-6 shifts the transcriptional state of myeloid cells away from antigen presentation and toward a clearance and tissue injury repair.

Of the 1,463 serum proteins measured in our dataset, the serum level of IL-6 was the most strongly associated with COVID-19 severity (22-fold higher in 1-Death than 5-Disch, 95% CI 17-28, P = 6e-47, Figure 6 N; Table S21), and it was consistently elevated across all time points (Figure S12 C). Since IL-6 receptor (IL6R) blockade has been shown to impact clinical outcomes in a subset of critically ill patients with COVID-19^6^, we hypothesized that IL-6 dependent myeloid dysregulation reflects a central hallmark of severity biology that might be improved by IL-6R blockade. To investigate this hypothesis, we characterized the transcriptional effects of IL-6R blockade on the host immune response at the single-cell level in patients hospitalized with COVID-19 during the alpha wave.

### Immunologic hallmarks of COVID-19 severity are reversed with tocilizumab

We screened the 243 patients from the BACC Bay randomized controlled trial of tocilizumab in patients with severe COVID-19 that was conducted at our institution (MGH) and others^54^. We focused our single-cell analysis on four patients who were identified, retrospectively, to be in the treatment arm and demonstrate clinical improvement following administration of tocilizumab after study unblinding (Table S3). In addition to clinical improvement, these patients had marked decay in serum C reactive protein (CRP) (measured for clinical care) (Table S22) and IL-6 levels (measured for research purposes) (Table S23) following tocilizumab administration (Figure 7 A; **Methods**). We performed scRNA-seq, CITE-seq, TCR and BCR sequencing on specimens collected immediately prior to tocilizumab administration and 4-, 7-, 14-, and 28-days post therapy (Figure 7 B, C, D).

**Figure 7.**
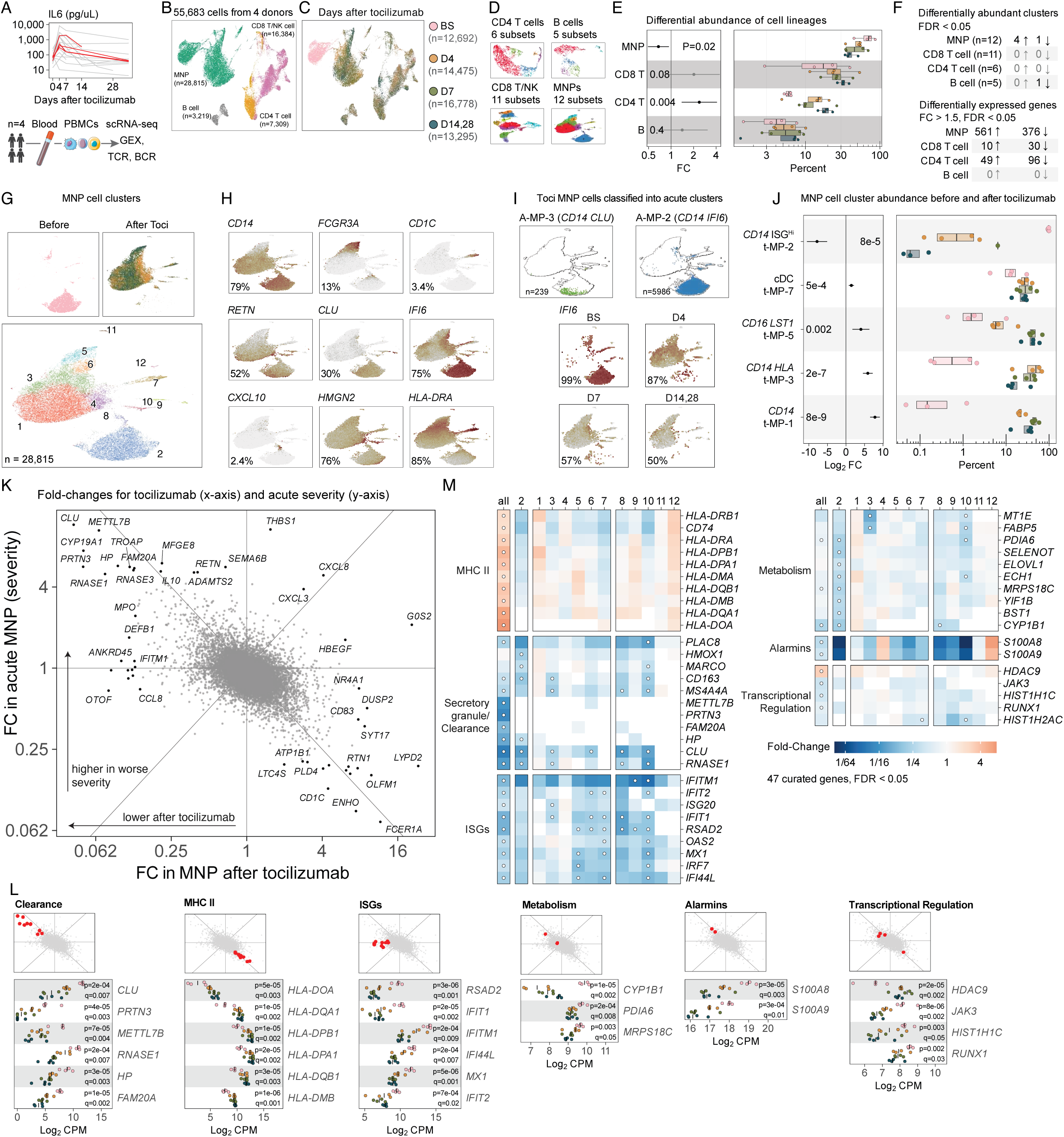
PBMC cellular and transcriptional associations in COVID-19 patients who received tocilizumab. **A.** IL-6 level over time points for multiple patients, red color indicates 4 selected patients for downstream profiling and analysis. Overview of blood sampling and data collection. **B.** UMAP embedding of all PBMCs. Hue indicates cell lineage. **C.** UMAP embedding of all PBMCs, hue indicates time after tocilizumab treatment (Baseline (BS), D4, D7, D14/28). **D.** UMAP embeddings for each cell lineage. Hue indicates cell subset. **E.** Differential abundance of cell lineages after tocilizumab. Dots represent patients, box plots indicate median and interquartile range, x-axis represents percent of cells from each cell lineage. Color indicates time point. Linear regression, two-sided p-value for the coefficient for severity. Error bars indicate 95% CI. **F.** Table summarizing the number of differentially abundant cell subsets in each cell lineage (left). Table summarizing the number of differentially expressed genes in each cell lineage (right). **G.** Top panels show UMAP embedding of tocilizumab MNP cells, top panels colored by time point, bottom panel colored by cell subset membership. **H.** UMAP embedding of tocilizumab MNP cells, color indicates gene expression for selected genes: *CD14*, *FCGR3A*, *CD1C*, *RETN*, *CLU*, *IFI6*, *CXCL10*, *HMGN2*, *HLA-DRA*. **I.** Top panels show classification of tocilizumab cells into acute cell subsets (A-MP-3 and A-MP-2). Bottom panels show log2CPM gene expression of *IFI6* faceted by time point after tocilizumab. Percent of cells with expression is shown. **J.** MNP cell subset abundance after tocilizumab. Linear regression, two-sided p-value for coefficient *after*, error bars indicate 95% CI. Dots represent patients, box plots indicate median and interquartile range, x-axis represents percent of cells from each cell lineage. Color indicates time point. **K.** Scatter plot of MNP fold-changes for genes associated with tocilizumab treatment (x-axis) and acute COVID severity (y-axis). Axes represent log2 fold-change associations from multivariate linear regression. Dots represent genes, selected genes are labeled. **L.** MNP gene expression levels for selected genes in each category from panel **K**. Small top panels indicate where genes fall in panel **K**. Bottom panels show log2CPM (x-axis), and color indicates time point after tocilizumab. **M.** Heatmap of fold-changes for selected genes in 6 categories: MHC II, Secretory granules/clearance, ISGs, Metabolism, Alarmins, Transcriptional Regulation. Color indicates fold-change. Dots indicate FDR < 5%. Columns indicate MNP cell subset, leftmost column indicates all MNP cells.

Tocilizumab treatment in this subset of recipients was associated with reversal of CD4 T and CD8 T cell lymphopenia, and a relatively lower abundance of all myeloid cells (Figure 7 E). The MNP compartment had the greatest number of changes in cell subset abundances and differentially expressed genes (Figure 7 F), in contrast to the minimal changes observed in the CD4 T, CD8 T, or B cell compartments (Figure S14).

In the myeloid lineage, we identified 12 MNP subsets with a distinct distribution pre- and post-treatment with tocilizumab (Figure 7 G). To map these cell subsets to our acute MNP subsets, we trained a linear discriminant analysis (LDA) classifier on the acute MNP data and then used it to classify each cell from the tocilizumab data (Figure 7 I; Figure S15). Notably, myeloid subset 2 in the tocilizumab data (t-MP-2) was no longer detectable after after treatment with tocilizumab (Figure 7 J; Figure S16 A, B, C, D, E). Cells from this subset mapped primarily to the sepsis-associated monocyte state subsets in the acute data (MP-2, MP-3) (Figure 7 I; Figure S15).

Upon closer examination of the cells within each myeloid cell subset over time, we noted that several clusters were divided into two transcriptionally distinct parts: one ISG^Hi^ (e.g., *IFI6*) and one ISG^Lo^ part. The ISG^Hi^ parts completely disappeared by day 14 within the classical CD14 monocytes (t-MP2, t-MP4, t-MP8), non-classical CD16 monocytes (t-MP6) and cDC (t-MP7) (Figure S16 C, D). We also identified a transient population of cycling classical MNPs (t-MP4) at day 4, which resolved by day 28 (Figure S16 E). By day 28, we observed the return of classical and non-classical monocytes to normal baseline (Figure S16 C, D, E), as well as cDCs (t-MP7) that no longer expressed markers of activation and ISGs (Figure 7 J; Figure S16 D).

Many of the severity-associated genes that were upregulated in the myeloid cells from severely ill patients were significantly repressed after tocilizumab treatment (Figure 7 K, L). These included genes related to secretory granules (*CLU*, *MS4A4*, *METTL7B*, *FAM20A*), genes encoding PAMP/DAMP receptors (*CD163*, *MARCO*, *STAB1*, *CD151*), metabolism pathways (*CYP1B1*, *FABP5*), alarmins (*S100A8*, *S100A9*), and ISG (*IFIT1*, *IFITM1*) (Figure 7 L, M; Figure S4 C; Figure S16 F, G; Table S24; Table S25).

Tocilizumab treatment led to repression of *JAK3* and induction of MHC II genes (e.g., *HLA-DQA1*) (Figure 7 L, M). IL-6 is known to suppress HLA-DR expression through JAK3-STAT3 signaling^52^, and our data demonstrates restoration of HLA-DR expression following tocilizumab administration. Our results establish a mechanistic link between the suppression of the sepsis-associated myeloid gene program and the activation of the antigen presentation gene program.

Since the most severely ill patients in our primary acute SARS-CoV-2 cohort effectively serve as a comparator for those reated with IL-6R blockade, these results provide a cellular and molecular basis for the immunomodulatory effects of tocilizumab in patients who had clinical improvement concomitant with pharmacologic therapy. Our results demonstrate that the IL-6/sepsis-associated monocyte state axis is a hallmark of severe COVID-19. Tocilizumab treatment eradicates the IL-6 dependent, immunosuppressive, sepsis-associated monocyte states (MP-2, MP-3), suggesting that these subsets contribute to the persistence of the dysregulated immune response. Additionally, the disappearance of the ISG^Hi^ myeloid subsets following treatment implies that viral clearance is occurring in parallel with reversal of immunosuppression and the restoration of antigen presentation capacity.

### Immunologic hallmarks in convalescent COVID-19

After identifying the central immunologic features of severe COVID-19 and demonstrating reversal of this dysregulation in the setting of IL-6R inhibition, we next examined if less severely ill patients — who achieved convalescence without antivirals or immunomodulatory treatment — had persistent hallmarks of acute disease. To assess the cellular and molecular changes associated with SARS-CoV-2 convalescence after non-severe infection, we performed scRNA-seq, CITE-seq, TCR, and BCR sequencing on more than 200,000 cells from 73 donors from a second cohort, independent of the previously described cohort of patients with acute SARS-CoV-2 infection (Table S2). Of these 73 donors, 43 had confirmed SARS-CoV-2 infection three months before sample collection, while the remaining 30 had serological studies consistent with absence of prior infection (Figure 8 A; **Methods**).

**Figure 8.**
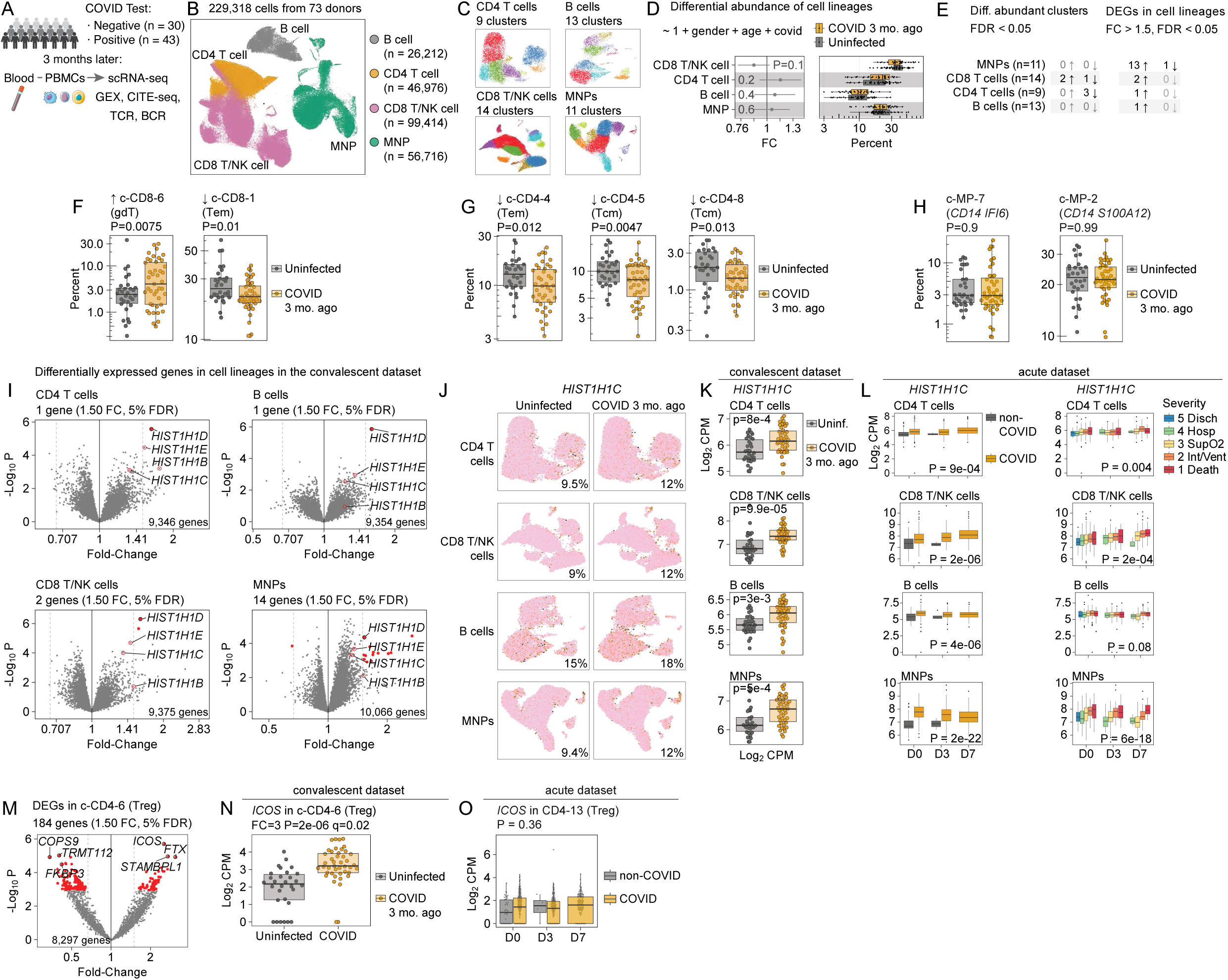
PBMC cellular and transcriptional associations in convalescent COVID-19 patients. **A.** Overview of patient selection and blood sampling 3 months after COVID infection. **B.** UMAP embedding of all PBMCs, hue indicates cell lineage. **C.** UMAP embeddings of cells from each lineage, hue indicates cell subset. **D.** Differential abundance of cell lineages, comparing patients who had COVID 3 months ago and healthy controls. Multivariate linear regression, two-sided p-value for the coefficient for COVID. Dots represent patients, box plots indicate median and interquartile range, x-axis represents percent of each donor’s cells that belong to each cell lineage. **E.** Table summarizing the number of differentially abundant cell subsets (left) and the number of differentially expressed genes (right) between convalescent COVID patients and healthy controls. **F, G, H.** Box plots of cell subset abundances, for cell subsets associated with convalescent COVID in CD8 T cells (**F**) and CD4 T cells (**G**) and MNPs (**H**). Dots represent patients, box plots indicate median and interquartile range, x-axis and hue represent convalescent COVID infection. p-values from multivariate linear regression. **I.** Volcano plots for genes associated with convalescent COVID infection in each cell lineage (facets). HIST genes are labeled. x-axis indicates fold-change, y-axis indicates negative log10 p-value. Red color indicates FDR < 5%. **J.** UMAP panels showing gene expression of *HIST1H1C* in cell lineages (rows) faceted across convalescent COVID infection (columns). Percent of cells with expression is shown. **K.** Box plots of *HIST1H1C* gene expression across cell lineages (rows). Dots represent patients, box plots indicate median and interquartile range, x-axis and hue represent convalescent COVID infection. P-value for coefficient for COVID infection from multivariate linear regression. **L.** Box plots of *HIST1H1C* gene expression across cell lineages (rows) in the acute dataset. Dots represent patients, box plots indicate median and interquartile range, x-axis represents time point, and hue represents COVID infection (left column) and COVID severity (right column). P-values from multivariate linear regression. **M.** Volcano plot for convalescent CD4 T cell subset 6 (Treg). Dots indicate genes, x-axis indicates fold-change, y-axis indicates negative log10 p-value from multivariate linear regression. Red color indicates FDR < 5%. **N.** Box plot of *ICOS* gene expression in CD4 T cell subset 6 (Treg) in the convalescent data. Fold-change and p-value is shown. Dots represent patients, box plots indicate median and interquartile range, x-axis and hue represent convalescent COVID infection. **O.** Box plot of *ICOS* gene expression in CD4 T cell subset 13 (Treg) in the acute data. Dots represent patients, box plots indicate median and interquartile range, x-axis represents time point, and hue represents COVID infection.

After clustering to cell lineage and subset resolution (Figure 8 B, C), we found that CD8 T/NK cells were slightly less abundant than other immune lineages in individuals with convalescent SARS-CoV-2 infection (Figure 8 D; Table S5). Within the CD8 T/NK lineage, convalescent donors had greater relative abundance of gdT cells (c-CD8-6) and lower abundance of effector memory T cells (Tem) (c-CD8-1) (Figure 8 F, Figure S17). Within the CD4 T cell lineage, convalescent donors had lower abundance of Tem (c-CD4-4) and two distinct subsets of central memory T cells (Tcm; c-CD4-5 and c-CD4-8) (Figure 8 G, Figure S18; Table S5). In contrast, B cells had significant differences in cell subset abundance or gene expression between uninfected and convalescent patients (Figure S19; Table S5). Similarly, within the myeloid lineage, cell subset abundances did not differ between the two groups (Figure S20; Table S5). For instance, convalescent and uninfected patients had similar levels of the CD14 subset most similar to our acute ISG^Hi^ classical monocytes (MP-2) (Figure 8 H left panel) as well as the sepsis-associated subset (MP-3) (Figure 8 H right panel). In contrast, both severe and non-severe acute patients had immediate changes in the abundance of these subsets (Figure 6 E). We conclude that many of the cellular changes present during acute infection return to baseline within three months of convalescence after infection.

When analyzed at the cell lineage level, only 18 genes were differentially expressed, 14 of which were in the MNP lineage (Figure 8 E), and most of the genes were related (Table S26). Several histone genes (*HIST1H1D*, *HIST1H1C*, *HIST1H1E*, *HIST2H2AC*) were elevated three months after infection in all four lineages (CD8 T cells, CD4 T cells, B cells, MNPs) (Figure 8 I, J, K). Some of these genes were also associated with severe disease in acute infection (Figure 8 L). While certain histone genes were uniquely associated with acute infection or convalescence, *HIST1H1C* and *HIST2H2AC* were induced in both (Figure S21 A, B, C). Notably, *HIST1H1C* transcription was also elevated in the blood in a study of 48 patients with PASC (Post-Acute Sequelae of COVID-19)^55^, and the encoded H1C protein has been shown to inhibit influenza virus replication^56^. Our results show that specific histone genes are transcriptionally induced during acute infection, associated with disease severity, and remain elevated in circulating immune cells during convalescence.

In the Treg cells from convalescent donors (c-CD4-6), we found higher expression of *ICOS* (FC = 3, P = 2e-6, FDR = 0.02) (Figure 8 M, N), but we did not find this in the Tregs from acute donors (Figure 8 O). However, *ICOS* was associated with worse severity in all CD4 T cells from acute patients (FC = 1.75, P = 2e-6, FDR = 2e-4, Table S6). *ICOS* is known to regulate immune tolerance and autoimmunity, and it may play a role in both post-acute sequelae and the immune response during recurrent infection^57^. In contrast, the ISGs induced during acute SARS-CoV-2 infection (e.g., *IFI27*, *IFI44L*, *IFIT3*, *IFIT1*) were no longer elevated in the convalescent individuals three months after infection (Figure S21 D, E).

Patients who were sampled three months after clearing SARS-CoV-2 infection had relatively subtle cellular abundance changes compared to what was observed in acute infection. The major sepsis-associated transcriptional hallmarks (e.g., repressed HLA, TCR, and BCR genes) were not observed in convalescent patients, indicating that these are molecular signals specific to acute infection and severe illness. However, we found that convalescent patients have persistent elevated expression of histone genes (e.g., *HIST1H1C*) in all immune cell types and elevated expression of *ICOS* in CD4 Tregs.

### A conceptual model of the biology of COVID-19 severity

We integrated our findings to propose a conceptual model of the host-response in acute SARS-CoV-2 infection (Figure 9). Our model is based on analysis of multiple modalities in acute disease across the spectrum of acute COVID-19 severity, the effect of immunomodulatory treatment, and persistent transcriptional changes that remain in convalescence. In this model, the effectiveness of the adaptive immune response is the primary bifurcation point between patients with severe and non-severe clinical illness.

**Figure 9.**
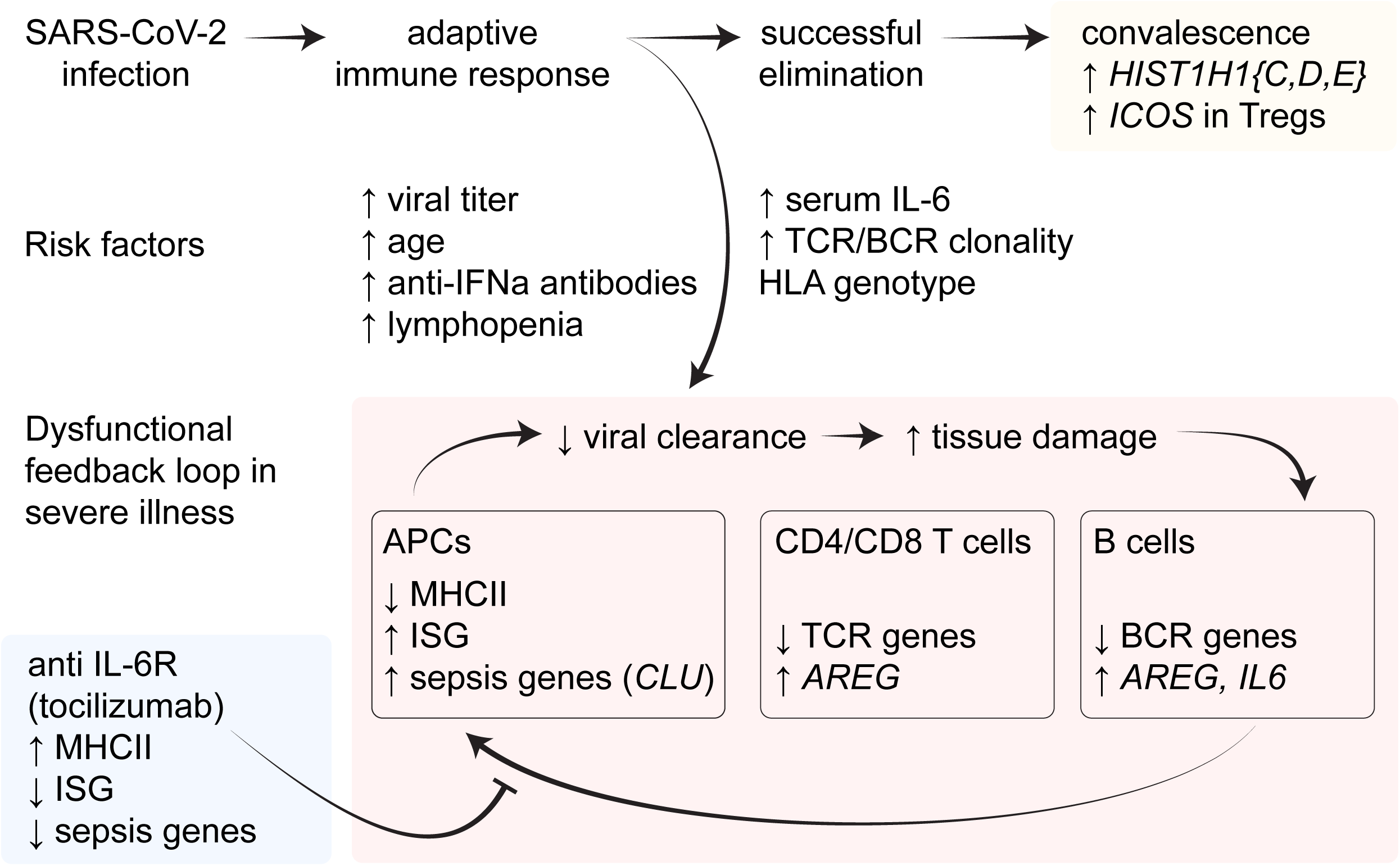
A schematic diagram of the factors that influence COVID severity.

Suboptimal T and B cell receptor repertoire selection and multiple etiologies (e.g., age-associated and age-independent increase in clonality, existing repertoire bias, HLA genotype, auto-antibody production, lymphopenia) lead to impaired viral clearance and consequent ongoing tissue injury and inflammation, including excess production of IL6. This leads to an IL-6 affected myeloid phenotype characterized by immunosuppressive sepsis-associated monocyte states and downregulation of antigen presentation which in turn promotes maladaptive lymphocyte behavior with a shared emphasis on tissue repair as opposed to viral clearance (Figure S22)^48^. In some patients, interruption of this cycle with IL-6 receptor blockade reverses the immunosuppressive sepsis-associated monocyte cell state and restores antigen presentation, which gives the adaptive immune system a second chance to promote viral clearance. Individuals infected with SARS-CoV-2 may have persistent transcriptional changes for months after infection, but these changes are different from the the changes in the acute phase of disease.

## Discussion

Our multimodal analysis of the immune response to acute SARS-CoV-2 infection in 351 patients reveals associations between disease severity and cell populations as well as molecular programs defined by scRNA-seq, and includes integrative analysis of CITE-seq, TCR and BCR sequencing, serum proteomics, auto-antibodies, viral load, anti-viral antibodies, and HLA genotypes. We show how immunologic intervention affects the hallmarks of disease severity and propose a conceptual model of immune dysfunction with maladaptive myeloid phenotype at the center of SARS-CoV-2 pathophysiology, providing insights into both COVID-19 severity and sepsis as a model of immune dysregulation.

Previous studies have identified myeloid dysfunction, T cell overactivation with exhaustion, extrafollicular B cell activation and atypical memory B cell phenotypes, TCR and BCR repertoire dynamics, autoantibody production, and direct viral cytotoxicity in the development of severe COVID-19^1,4,13,58–64^. We propose three central themes that integrate these previously reported observations with our findings: (1) impaired antigen recognition early in disease leads to severe COVID-19 later on; (2) severe disease is characterized by a shift toward responses to persistent inflammation and tissue injury, with reduced antigen presentation and effector function despite clear evidence of ongoing viral infection; (3) this dysfunctional immune state is sustained by IL-6 and specific subsets of myeloid-derived suppressor cells (MDSCs)^48,65^ previously associated with sepsis^66^, which contribute to the immunosuppressive environment and are associated with disease severity. Importantly, these MDSCs have also been referred to as MS1 cell state in the context of acute SARS-CoV-2 and sepsis^1–4^.

With respect to the first theme, we identify CD21^Lo^ atypical memory B cells – characterized by *ITGAX*, *CD1C*, *EMP3* – as the only B cell subset expanded in more severe disease, consistent with previous studies^62,67^. Higher proportions of CD21^Lo^ B cells have been associated with poorer COVID-19 vaccine response^68^. CD21 expression is typically reduced in apoptotic B cells, and an increased abundance of CD21^Lo^ B cells has been reported in individuals with systemic lupus erythematosus, chronic variable immunodeficiency, rheumatoid arthritis, human immunodeficiency virus (HIV) viremia, and exposure to *Plasmodium falciparum*^69^. Notably, several marker genes of the B-5 cell subset (*SOX5*, *FGR*, *ITGAX*) in our dataset are shared with a previously described population of expanded anergic B-cell subset in hepatitis C virus-associated mixed cryoglobulinemia^69^. Although some studies suggest these CD21^Lo^ atypical memory B cells can produce antibodies and autoantibodies, we did not observe a correlation between their abundance and neutralizing antibody titers in our dataset; only a small proportion of our patients had evidence of autoantibody production.

However, the majority of expanded public BCR clones in our data were localized within the severity-associated CD21^Lo^ atypical memory B cell subset (B-5), and increased B cell clonality was associated with worse disease severity. These findings demonstrate the dysfunctional B cell immune response associated with COVID-19 severity.

We found that increased CD8 TCR clonality early in disease was associated with worse severity, consistent with previous studies^70,71^, and consistent with known age-dependent decline in CD8 T cell numbers and narrowing of the TCR repertoire^72^. We identified specific severity-associated TCR genes, the majority of which were predicted not to bind SARS-CoV-2 epitopes, indicating that patients with severe disease may be selecting inefficient or suboptimal T cell clones. Our data suggests that a constrained repertoire may increase the likelihood of selecting suboptimal clones. Conversely, patients with less severe disease showed increasing clonality over time, consistent with the selection and expansion of clones that recognize SARS-CoV-2 epitopes.

In our data, aspartic acid (D) at position 57 in *HLA-DQB1* was associated with greater disease severity and higher viral load following SARS-CoV-2 infection. In contrast, the absence of aspartic acid at position 57 was associated with lower abundance of CD8 T cells expressing severity-associated TCR genes. Moreover, the *HLA-DQB1* locus has also been associated with PASC^73^ and with failure to seroconvert after COVID-19 vaccination^74^. Class II HLA proteins, such as HLA-DQB1, present antigens to CD4 T-helper cells, which in turn modulates cytotoxic CD8 T cell development via DC cross-priming^75,76^. In COVID-19, delayed generation of circulating CD4 T follicular helper cells (cTfh) has been associated with increased disease severity^36^. Therefore, our data suggest that COVID-19 severity is associated with antigen presentation by HLA-DQB1 to CD4 T helper cells, which influences development of anti-viral CD8 T cells and antibodies.

With respect to the second theme, we propose that multiple features of adaptive immune response, occurring either independently or in combination, can impair viral clearance, ultimately leading to a maladaptive immune state characterized by persistent tissue injury and inflammation. Despite persistent ISG signatures in both the T and B cell lineages, we observed transcriptional repression of genes involved in effector function, TCR and BCR signaling, and the immune synapse formation. Concurrently, genes associated with tissue injury and repair (e.g., *AREG*, *CH25H*, *DDIT4*) were upregulated. Viruses are known to inhibit TCR signaling^77^, and some hypothesize that SARS-CoV-2 may directly inhibit TCR signaling^78^. These adaptive immune alterations are paralleled in the myeloid compartment by broad repression of antigen presentation gene and protein expression. Since NFKB is required for MHC class II-mediated antigen presentation^79^ — and SARS-CoV-2 requires NFKB for viral replication^80^ — viral hijacking of NFKB may contribute to the observed repression of antigen presentation in MNPs from patients with acute COVID-19.

Our results highlight that COVID-19 severity is associated with extensive transcriptional changes across myeloid lineage subsets, and increased abundance of myeloid subsets with immunosuppressive profiles characteristic of MDSCs. Previous studies using electron microscopy and flow cytometry to characterize circulating immune cells showed that SARS-CoV-1 infects approximately 50% of lymphocytes and 30% of monocytes^16,81–83^, and that monocytes exhibit anisocytosis (extreme intracellular vacuolization)^81,84^. In the context of SARS-CoV-2, increased monocyte distribution width (MDW), which is a potential clinical indicator of anisocytosis and monocyte activation, has been observed in several studies^85–91^. In our dataset, severity-associated gene expression programs, such as those related to secretory granules, support the idea that increased vacuolization and heightened secretory behavior in myeloid cells is a hallmark of severe COVID-19. These findings suggest that MDW could offer a low-cost, clinically accessible marker of myeloid dysregulation, but additional studies are needed to directly connect this phenotype to the more granular molecular changes identified here and in other single-cell studies.

With respect to the third theme, MDSC phenotypes, as well as broader repression of HLA-DR expression, have been shown to be driven by IL-6/JAK3/STAT3 signaling. Previous studies showed that IL6R blockade can restore HLA-DR expression in the myeloid compartment and reverse bulk-transcriptomic signatures of myeloid dysfunction^2,92,93^. We demonstrate that IL6R blockade through tocilizumab results in the loss of MDSC cell state, restoration of HLA-DR expression, and re-establishment of MNP subset homeostasis, as well as reversal of T cell lymphopenia. Notably, these changes were associated with the disappearance of ISG signatures in all immune cell lineages, suggesting that resolution of the MDSC cellular state facilitated viral clearance. In contrast, transcriptional changes within the B and T cell lineages were minimal following tocilizumab treatment. Our data support a central biological role for MDSC subsets in maintaining a dysfunctional immune state that impairs viral clearance, and suggest that MDSC may represent a rational therapeutic target in a subset of patients with sepsis.

The notable difference between some randomized clinical trial (RCT) data showing limited benefits of IL-6 antagonism in COVID-19 and the clear immunologic effect our tocilizumab cohort single-cell multiomic analyses identified deserves special mention. Indeed, RCT conducted from April 2020 through 2021 during the alpha wave of the pandemic produced mixed results for IL-6 antagonist therapy in COVID-19. Some trials found no mortality benefit^54,94–96^, while others (or subgroup analyses of negative trials) showed improvements such as reduced progression to mechanical ventilation and end-organ support, lower 28-day mortality, and higher discharge rate at 28 days with tocilizumab therapy^6,97,98^. These differences in results stem from trials enrolling patients across a spectrum of COVID-19 disease severity, CRP levels, and co-morbidities — a reflection of the unprecedented challenges and need to rapidly study immunomodulatory treatment for a new, severely morbid viral infection. Careful review of subgroup analyses and individual RCT data suggests that IL-6 inhibition is most beneficial for patients with moderate to severe and/or critical COVID-19 disease^97,99^, especially those with high CRP^95^, and for those who received tocilizumab (as opposed to other IL-6 antagonists)^100^. Working within the limitations of the study design of our institution’s BACC BAY RCT — where PBMC were collected prospectively to study the immunology of IL-6 inhibition, and where inclusion criteria study design targeted hospitalized individuals with moderate COVID-19 disease necessitating supplemental O2 — we analyzed PBMCs from four individuals who had the most severe disease, highest pre-tocilizumab CRP and IL-6 levels, largest drop in these laboratory markers post-treatment, no other immunosuppressive or remdesivir therapy, and who ultimately were discharged home from the hospital without the need for mechanical ventilation.

Given this rigorous approach marrying subgroup analyses of published RCT data and our carefully selected patient cohort, we believe that the reversal of the MDSC/MS-1 myeloid state following tocilizumab we have identified in this study is a critical component of the mechanism underlying IL-6R blockade benefit in severe SARS-CoV-2 infection. This conclusion is further reinforced by our findings that two immunological hallmarks of severe COVID-19 (i.e., abundance of MNP subsets ISG^hi^ MP-2 and MDSC MP-3) and transcription of severity-associated genes (i.e. *CLU*) resolved by day 4 post-tocilizumab (Figure 7 I, L), much faster than in the untreated patients (Figure 6 E, K).

In convalescent patients who recovered from COVID-19 three months after infection, we did not find evidence of the MDSC cell state. This suggests that this immunosuppressive phenotype is a feature of acute disease and does not persist during recovery. Importantly, MDSCs can regulate inflammation and limit tissue injury. One example of the protective role of MDSC is their expression of *CLU*, which encodes clusterin, an enzyme that binds pro-inflammatory histones released by neutrophils and dying cells^53^. In mouse models of sepsis, *CLU* deficiency leads to increased mortality^53^, underscoring its protective role in mitigating inflammatory tissue injury. As such, immunomodulatory therapies that reduce *CLU* expression, such as IL-6 blockade as observed in our data, could be harmful in patients who are mounting an appropriate host immune response to tissue injury. A key limitation of our tocilizumab response analysis is that it focused exclusively on patients who clinically improved, potentially skewing the interpretation towards beneficial effects of MDSC suppression. Future studies will be needed to determine whether markers of myeloid dysregulation, such as *CLU* expression, can guide personalized treatment decisions and identify which patients will likely benefit or be harmed by immunomodulatory therapy.

In convalescent patients, we also observed a persistent elevation of *HIST1H1C* expression across all immune cell lineages, which was also found to be elevated in patients with PASC^55^. Eukaryotic histones have ancient evolutionary history with viruses^101^, and the SARS-CoV-2 protein ORF8 mimics a histone H3 motif^102^, suggesting a possible mechanism for viral interference with host chromatin regulation. Furthermore, H1C has been shown to bind dendritic cells and induce cytokine storm^103^, as well as mediate immune evasion by down-regulating MHC-I expression^104^. We also found that Tregs from convalescent patients have persistently higher *ICOS* expression, a co-stimulatory molecule known to be elevated in autoimmune diseases^57^. Although we saw most acuity-associated immune alterations resolve three months into convalescence, some changes persist related to tolerance, autoimmunity, and response to viral rechallenge.

Our comprehensive characterization of the host immune response in acute and severe SARS-CoV-2 infection provides an unparalleled multimodal dataset with a depth and scale that will allow exploring questions fundamental to SARS-CoV-2 and human immunology. This resource can be used to generate new hypotheses and to validate future findings from a wide range of investigations related to COVID-19, sepsis, and immune dysfunction.

## Materials and Methods

### Material Availability

This study did not generate new unique reagents.

### Data and code availability

Data will be made available upon publication. The scRNA-seq, OLINK, and antibody data and analysis results will be available for browsing on our own interactive website.

Analysis results will be archived on Zenodo. Count matrices (scRNA-Seq, TCR, BCR, and CITE-seq) and related data will be deposited to NCBI GEO. Source code for data analysis will be available on GitHub and Zenodo. The raw RNA sequencing data reported in this study cannot be deposited in a public repository because these data were collected at the beginning of the COVID-19 pandemic, and as such, a waiver of informed consent was approved by the Massachusetts General Hospital governing institutional review board, in compliance with the Code of Federal Regulation (45CFR 46, 2018 Common Rule). To protect the identity of individual subjects, public posting of raw sequencing data from the patients has not been approved; therefore, raw data is not provided.

### Patient selection

As previously described^13^, patients were enrolled in the Emergency Department (ED) of a large, urban, academic hospital in Boston from March 24, 2020 to April 30, 2020 during the peak of the initial COVID-19 surge. All study procedures involving human subjects were approved by the Mass General Brigham (formerly Partners) Human Research Committee, the governing institutional review board at Massachusetts General Hospital. A waiver of informed consent was approved in compliance with the Code of Federal Regulations (45CFR 46, 2018 Common Rule). Included were patients 18 years or older with a clinical concern upon ED arrival for COVID-19 and with acute respiratory distress, with at least one of the following: 1) tachypnea (22 breaths per minute), 2) oxygen saturation %92% on room air, 3) a requirement for supplemental oxygen, or 4) positive-pressure ventilation. The day 0 blood sample (N = 384) was obtained concurrent with the initial clinical blood draw in the ED, and day 3 (N = 217) and day 7 (N = 143) samples were obtained for COVID-19-positive patients, if still hospitalized at those times, yielding 744 samples. In addition, blood was collected from some patients at the time of substantial clinical deterioration (44 samples); these event-driven samples were excluded from linear models. Clinical course was followed to 28 days post-enrollment or until hospital discharge, if that occurred after 28 days. Patients were classified by acuity levels A1-A5 on days 0, 3, 7, and 28 (WHO Ordinal Outcomes Scale) where the acuity levels are described as follows: A1, death within 28 days (N = 42, 14%); A2, intubation, mechanical ventilation, and survival to ≥28 days (N = 67, 22%); A3, hospitalized and requiring supplemental oxygen (N = 133, 43%); A4, hospitalized without requiring supplemental oxygen (N = 41, 13%); and A5, discharged directly from the ED without subsequently returning and requiring admission within 28 days (N = 23, 8%). A1 and A2 were classified as severe (N = 109) and A3-A5 as non-severe (N = 197). Of all 384 enrolled, 78 (20%) tested negative for SARS-CoV-2; among these, for 50 (64%), suspicion for COVID-19 was very low based on careful retrospective chart review by M.R.F. and R.P.B., an emergency physician and infectious diseases physician, respectively. Among the remaining 28 patients, COVID-19 was a diagnostic possibility, yet most had multiple negative PCR tests during their hospital course. While these 78 subjects were categorized as controls, we only generated and analyzed scRNA-seq data from the 50 subjects with very low suspicion for COVID-19.

Demographic, past medical history and clinical data were collected and summarized for each outcome group, using medians with interquartile ranges and proportions with 95% confidence intervals, where appropriate. Detailed clinical data, including age, gender, ethnicity, and race, are summarized for all outcome cohorts in Table S1. Patient-level clinical data are available from previously the published study on Mendeley Data: https://data.mendeley.com/datasets/nf853r8xsj/1. To protect the identity of individual subjects, public posting of patient-level demographic information is limited as required by the Mass General Brigham Human Research Committee.

### Tocilizumab cohort study design

To understand how IL-6R inhibition modulates the acuity-specific immunity in SARS-CoV-2 infection, we obtained clinical specimens obtained as part of the BACC-BAY trial (NCT04356937), one of the first randomized, double-blind, placebo-controlled trials performed during the alpha-wave of the COVID-19 pandemic to evaluate the efficacy of tocilizumab at reducing multi-organ dysfunction (MOD). MOD was measured as the incidence of the composite endpoint of need for mechanical ventilation, renal replacement therapy, mechanical support, inotropes or vasopressors, and liver dysfunction and all-cause mortality in patients hospitalized with confirmed SARS-CoV-2 infection. In this trial, individuals with confirmed SARS-CoV-2 infection and hyperinflammatory clinical state as defined by fever (body temperature >38°C), pulmonary infiltrates, or the need for supplemental oxygen to maintain an oxygen saturation greater than 92% were randomly assigned in a 2:1 ratio to receive standard care plus a single dose of either tocilizumab or placebo. Of note, while the recruitment of research participants for this BACC-BAY trial and the MGH-specific acute cohort described in this paper were independent and unrelated, these studies did recruit patients contemporaneously from March through June 2020. After the results of this multi-center trial were published and approval was obtained from the trial’s data safety monitoring board to share unblinded data, we selected four research participants from this trial hospitalized at MGH from whom longitudinal PBMC collected at “baseline” (pre-tocilizumab, at point of randomization), and day 4, 7, and 21 or 28 post tocilizumab administration was available (Table S3). We performed 5’ V2 scRNA-seq/CITE-seq/TCR/BCR sequencing with the identical experimental approach as was undertaken for the central MGH acute cohort. The four patients were chosen based on specific parameters, including having the highest absolute IL-6 levels and C-reactive protein (CRP) at the time of randomization into the tocilizumab arm of the trial, and demonstrating the greatest decrease in IL-6 and CRP levels over a 28-day period following administration of tocilizumab (Figure 7, Table S22, Table S23). Detailed review of the medical record demonstrated that PBMC obtained from these four patients in the tocilizumab arm of the trial at the “baseline” time point correlated with 8-11 days after onset initial symptoms. Thus, temporally, “baseline” samples from this tocilizumab mini-cohort correlated most closely with day 0 of the MGH acute cohort. This four-person tocilizumab cohort was comprised of three males and one female aged 50-60 years old, all of whom were identified as patients hospitalized with COVID-19, requiring supplemental oxygen but not hospitalized in the intensive care unit setting and not requiring mechanical ventilation. These research participants did not receive remdesivir nor any additional concomitant immunosuppression other than tocilizumab, as it was not yet standard of care during the initial weeks of the alpha wave of the pandemic. All four patients survived their hospitalization and were discharged home following improvement in their COVID-19 infection.

### Convalescent cohort study design

Whole blood (collected in BD Vacutainer K2EDTA tubes) and serum (collected in BD Vacutainer serum tubes) samples from consented subjects recovered from COVID were collected in EDTA tubes at the Brigham and Women’s Hospital, Boston, MA, USA. Results from PCR testing when available, disease and demographic information (Table S2) were collected after blood draws through a RedCap-administered survey. All studies were performed under the IRB protocol number 2020P000849 “Biorepository for Samples from those at increased risk for or infected with SARS-CoV-2” approved at the Brigham and Women’s Hospital, Boston, MA, USA.

### Blood specimen processing

Blood samples were collected in EDTA tubes and processed no more than 3 hours post blood draw in a Biosafety Level 2+ laboratory on site. Whole blood was diluted with room temperature RPMI medium in a 1:2 ratio to facilitate cell separation for other analyses using the SepMate PBMC isolation tubes (STEMCELL) containing 16 mL Ficoll (GE Healthcare). PBMCs were collected using a Ficoll density gradient. Diluted whole blood was centrifuged at 1200 g for 20 minutes at 20C. After centrifugation, plasma (5 mL) was pipetted into 15 mL conical tubes and placed on ice during PBMC separation procedures, centrifuged at 1000g for 5 min at 4C, aliquoted into cryovials, and stored at 80C. For the PBMCs, the interphase was collected upon density gradient centrifugation and placed in a new 15 mL conical tube with p to 10ml of RPMI-10 (with human AB serum; Sigma Aldrich) to bring the volume to 15 mL and centrifuged at 330 g for 10 mins with high brake. PBMCs were the cryopreserved using CryoStor CS10 freezing media (BioLife Solutions) with up to 5–10 million cells/cryovial, placed in a chilled Mr. Frosty, stored at −80 °C for one day, then transferred to liquid nitrogen for long-term storage.

### PBMC CD45+ enrichment, cell hashing, CITE-seq staining

Cryopreserved PBMC samples were thawed at 37 °C, diluted with a 10x volume of RPMI with 10% heat-inactivated human AB serum (Sigma Aldrich), and centrifuged at 300 g for 7 mins. Cells were resuspended in CITE-seq buffer (RPMI with 2.5% [v/v] human AB serum and 2 mM EDTA) and added to 96-well plates. Dead cells were removed with an Annexin-V-conjugated bead kit (Stemcell Technologies, 17899) and red blood cells were removed with a glycophorin A-based antibody kit (Stemcell Technologies, 01738); modifications were made to the manufacturer’s protocols for each to accommodate a sample volume of 150 μL. Cells were quantified with an automated cell counter (Bio-Rad, TC20), after which, 2.5 x 105 cells were resuspended in CITE-seq buffer containing TruStain FcX blocker (BioLegend, 422302) and MojoSort CD45 Nanobeads (BioLegend, 480030). Hashtag antibodies (BioLegend) were added to samples (Table S27) followed by a 30 mins incubation on ice. Cells were then washed three times with CITE-seq buffer using a magnet to retain CD45+ cells. Live cells were counted with trypan blue, and samples from each cohort bearing different hashtag antibodies were pooled together at equal concentrations. All samples were pooled in groups of eight and loaded on one 10x Genomics channel; all 8 samples for each pool were balanced for age, gender, disease severity level, and co-morbidities.

Pooled samples were filtered with 40 μM strainers, centrifuged, and resuspended in CITE-seq buffer with TotalSeq-C antibody cocktail (BioLegend; Table S28). Cells were incubated on ice for 30 mins, followed by three washes with CITE-seq buffer and a final wash in the same buffer without EDTA (RPMI with 2.5% [v/v] human AB serum). Cells were resuspended in this buffer without EDTA, filtered a second time, and counted.

### Single-cell gene expression, feature barcode, and TCR/BCR library construction and sequencing

An input of 50,000 hashed PBMC samples was loaded on a single channel in the 10X Chromium instrument for blood immune cell analysis, aiming for a recovery goal of 25,000 single cells. Hashed PBMC single-cell libraries were generated with the Chromium Single Cell 5’ kit (V2 NextGEM 10X Genomics, PN-1000263) together with the 5’ Feature Barcode library kit (10X Genomics PN-1000256) according to manufacturer’s protocols. PCR-amplified cDNA was used for TCR and BCR enrichment with the Chromium Single Cell V(D)J Enrichment kit (10 Genomics PN-1000252 and PN-1000253). The library quality was assessed with an Agilent 2100 Bioanalyzer. All gene expression, feature barcode, TCR and BCR libraries were sequenced on an Illumina NovaSeq using the S4 300 cycles flow with the following sequencing parameters: (read 1 = 26; read 2 = 90; index 1 = 10; index 2 = 10).

Measurements from the three modalities were measured in the same cells and could be associated using the cell barcode.

### Autoantibody methodology

A 76-plex custom, bead-based antigen array was created to profile serum samples for IgG antibody reactivities. The array consisted of three broad categories of antigens: (i.) cytokines, chemokines and growth factors to measure autoantibodies against secreted proteins; (ii.) autoimmune disease-associated antigens; and (iii.) viral antigens. The “Cytokine” content included 47 cytokines, chemokines, growth factors, acute phase proteins, and cell surface proteins. The “Traditional Autoimmune-Associated” content included 18 commercial protein antigens associated with connective tissue diseases. The “Viral” content included 7 antigens derived from viruses such as SARS-CoV-2, Epstein Barr Virus (EBV) and cytomegalovirus (CMV). The names, vendors, and catalogue numbers of all antigens are listed in Table S29.

Each array was constructed and used to profile serum samples as previously described^105^. Antigens were covalently conjugated (Table S29) to uniquely barcoded, carboxylated magnetic beads (MagPlex-C, Luminex Corp.). The resulting bead array was distributed into a 384-well plate (Greiner BioOne) by transferring 5 µl of bead array per well. 45 µl of diluted serum or plasma sample (1:100) were transferred into the 384-well plate containing the bead array. Samples were subsequently incubated for 60 min on a shaker at room temperature and left overnight in 4°C. The following morning, beads were washed with 3 × 60 µl PBS-Tween on a plate washer (EL406, Biotek) and incubated with 50 µl of 1:1000 diluted R-phycoerythrin (R-PE) conjugated Fc-γ-specific goat anti-human IgG F(ab’)2 fragment (Jackson ImmunoResearch, 106-116-098) for 30 min. The beads were then washed with 3 × 60 µl PBS-Tween, re-suspended in 50 µl PBS-Tween, and analyzed with a FlexMap3DTM instrument (Luminex Corp.). Beads were validated using monoclonal antibodies, or prototyle plasma or serum samples with known reactivities.

For each run, positive controls were included, consisting of prototype human plasma or serum samples derived from participants with confirmed prior SARS-CoV-2 infection and/or autoimmune diseases with known reactive patterns (e.g., Scl-70, centromere, SSA, SSB, whole histones, RNP, anti-IFN, etc.). Healthy controls were obtained from the Stanford Biobank prior to the pandemic. Binding events were displayed as Median Fluorescence Intensity (MFI). For each sample, as normalization, MFI values for “bare bead” IDs were subtracted from the MFI values for each antigen-conjugated bead ID.

### Pre-processing of single cell and single nucleus RNA-seq data and quality control (QC) filtering steps

We used 10X Genomics Cell Ranger (version 4.0.0) to pre-process raw single-cell RNA-seq data, including demultiplexing FASTQ reads, aligning reads to the human reference genome that includes the SARS-CoV-2 transcriptome (md5sum bab4d11d604a7d28d63d8cdb7f55a74b GRCh38_and_SARSCoV2.tar.gz)^106^, and counting the unique molecular identifiers (UMIs) to produce a count matrix with one row for each gene and one column for each cell. We used the Terra platform (https://app.terra.bio/) to run Cell Ranger via the workflow script that is part of a collection called Cumulus^107^. We used R scripts to aggregate the data and filter out poor quality cells that had fewer than 500 genes detected (at least 1 read) or more than 20% of reads from 13 mitochondrial genes (MT-ND6, MT-CO2, MT-CYB, MT-ND2, MT-ND5, MT-CO1, MT-ND3, MT-ND4, MT-ND1, MT-ATP6, MT-CO3, MT-ND4L, MT-ATP8).

### Quality control filtering and demultiplexing single cell RNA-seq data

For the samples that were multiplexed, we used both genetic and hashing antibody-based demultiplexing to recover as many cells as possible from each sample. To pass quality control, we required each cell to have more than 500 genes detected and percent of reads from mitochondrial genes greater than 0.01% and less than 20%. Next, we also require that the cell is called a singlet by the Souporcell algorithm^108^.

To assign each cell to a hashtag barcode, we ran the demuxEM^109^ algorithm on the matrix of counts for each cell of 8 possible hashtag barcodes (8 by N cells). To assign each cell to a genotype, we ran the Souporcell algorithm on the scRNA-seq read alignment data, and we matched the cell genotypes to the known genotypes of each sample from bulk RNA-seq data^12^. Next, we used the following algorithm to match each of the 8 possible hashtag barcodes to each of the 8 possible genotypes in a sample. For each hashtag, we compute the percentage of cells with that hashtag and each of the 8 genotypes. For each genotype, we compute the percentage of cells with that genotype and each of the 8 hashtags. For each genotype, we choose the match with the greatest sum of percentages from the previous two steps. We discard matches that include less than 50% of the hashtag’s cells or less than 50% of the genotype’s cells.

### Normalizing count data

For a given single-cell counts matrix, we first divide each cell’s counts by the sum of its counts to get relative abundances. Next, we multiply the relative abundances by the median number of genes detected per cell to get CPM values. Finally, we add 1 to the values and use report log2(CPM +1) values for each gene in each cell, and we refer to this quantity as log2CPM in the figures.

### Selection of highly variable genes and principal component analysis

We selected highly variable genes, centered and scaled each gene, computed principal components, and ran the molecular cross validation (MCV) algorithm to select the optimal number of principal components for downstream analysis^110^.

To identify highly variable genes, we first used the raw counts matrix to compute a mean with Matrix::rowMeans() and standard deviation with proxyC::rowSds() for each gene, excluding genes that are detected in fewer than 50 cells. Next, we fit a local 2nd degree polynomial regression with stats::loess() with the formula log10(sd) ∼ log10(mean) to model the relationship between mean and variance for each gene’s counts. We computed the residual variance for each gene, selected the top 80% of genes with the greatest residual variance, and centered and scaled each gene to have mean zero and unit variance. We computed the singular value decomposition (SVD) of the scaled expression matrix and got principal component scores for each cell.

To select the optimal number of principal components, we used the MCV algorithm. With the raw count matrix, we randomly split the counts for each cell, resulting in two count matrices *A* and *B* whose sum is equal to the original matrix. With the normalized, centered, and scaled matrix *A’*, we computed principal components. Then, we reconstructed matrix *A’* from the top *K* PCA scores and loadings (*UVT*) and estimated a reconstruction loss with the mean squared error. We also reconstructed matrix *B’* using the PCA scores computed from *A’*, resulting in an estimate of the MCV loss. We recorded the losses for each choice of top *K* principal components between 2 and 80.

Then, we chose the *K* that minimized the MCV loss. These top *K* principal components were used in downstream analysis such as low dimensional embedding with UMAP and cell cluster prediction with linear discriminant analysis (LDA).

### Batch integration, cell clustering, and two-dimensional embedding

We used the Harmony algorithm to align PCA scores across batches of data^111^ and then we create a nearest neighbor network of cells by connecting each cell to the 50 cells with the least Euclidean distance in the space of the harmonized PCA scores. We used the Leiden algorithm^112^ from the leidenalg Python package on the nearest neighbor network to identify cell clusters (10 iterations). We used the UMAP algorithm^113^ to embed the network in two dimensions (spread = 1, min_dist = 0.25).

### Marker gene identification and cell cluster annotation

We used two complementary strategies to identify sets of marker genes uniquely expressed in each cell lineage and each cell subset. The first strategy uses presto::wilcoxauc() to compute the area under receiver operator curve (AUROC or AUC) for the log2CPM value of each gene as a predictor of the cluster membership for each cluster versus all other clusters^114^.

The second strategy uses a pseudobulk expression matrix^115^ for the cells in a cluster for each patient sample, such that the pseudobulk matrix has one row for each gene and one column for each cluster from each patient sample. We normalized the pseudobulk counts to log2CPM as described above. Then we used limma::lmFit() to test for differential gene expression between the samples in a cluster versus all other clusters, using the log2CPM pseudobulk matrix^116^.

We also created another pseudobulk expression matrix by summing over all cells in one of four cell lineages (CD8 T cells, CD4 T cells, MNPs, B cells). We found gene expression differences (AUC ≥ 0.7; pseudobulk FDR < 0.05) at the level of cell lineage as well as at the level of each cell cluster. We then annotated each cell cluster by using marker genes (i.e., area under the curve (AUC) ≥0.7, pseudobulk FDR < 0.05), which we also cross-referenced with published immune transcriptional signatures. We also used CITE-seq protein data (n = 197 proteins) to help annotate cellular subsets. For each protein, we plotted the expression on the UMAP embedding to visualize which of our cell clusters is marked by each protein.

### Cell type abundance association with disease acuity

We analyzed each cell lineage separately (B cells, CD4 T cells, CD8 T cells, MNPs). For each patient sample, we computed the percentage of cells that are assigned to each cell cluster. We used limma to fit linear models to the log2(percent) of cells in each cluster^116^. This is the main model we used to find cell clusters associated with COVID infection and severity (acuity_max):

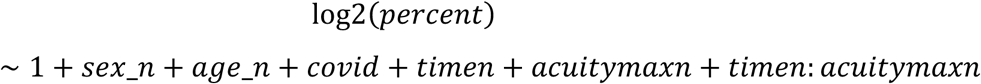

Next, we fit five additional models (one for each factor) to find the cell clusters associated with: viral load (vl), autoantibodies to interferon alpha (aa), neutralizing antibodies against SARS-CoV-2 (neut), and abundance of IL6 protein in blood (il6). Each of these additional models is the main model (shown above) with one additional coefficient.

Sex is encoded as a numeric variable *sex_n* [0, 1] where 1 indicates male and 0 indicates female. Age is encoded as a numeric variable *age_n* on the scale [0, 1] as follows:

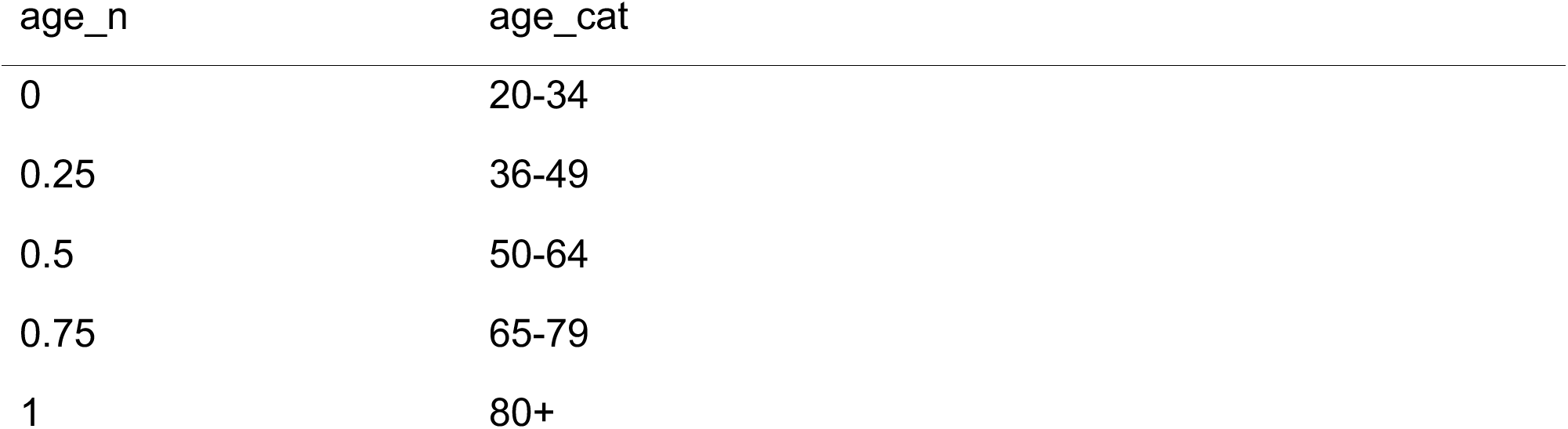

COVID status is encoded as a binary variable *covid* [0, 1] where 1 indicates COVID-positive and 0 indicates COVID-negative.

Time point is encoded as a numeric variable *timen* as follows:

| timen | time_point |
| --- | --- |
| -0.429 | Day 0 |
| 0 | Day 3 |
| 0.571 | Day 7 |

Maximum disease acuity is encoded as a numeric variable *acuity_maxn* as follows:

| acuity_maxn | acuity_max_cat |
| --- | --- |
| -0.5 | 5 Disch |
| -0.25 | 4 Hosp |
| 0 | 3 SupO2 |
| 0.25 | 2 Int/Vent |
| 0.5 | 1 Death |

### Association of gene expression with disease acuity and other factors

We used limma to fit linear models to the log2(CPM+1) of each gene in the pseudobulk expression data at the level of cell lineage, and also at the level of cell cluster. This is the main model we used to find genes that are associated with COVID acuity:

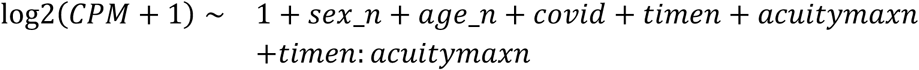

### Percent Neutralization

Percent neutralization was defined by LaSalle et al.^12^. Briefly, an *in vitro* assay was used to quantify the degree of SARS-CoV-2 viral neutralization when exposed to serum from each patient sample.

### Association of cell cluster abundance and gene expression with serum protein levels (OLINK)

Serum protein measurements of 1,463 proteins in serum from each patient sample were taken from Filbin et al.^13^.

For each scRNA-seq cell cluster and each OLINK protein, we fit a linear model with limma to find proteins associated with cell cluster abundance (at the level of patient samples):

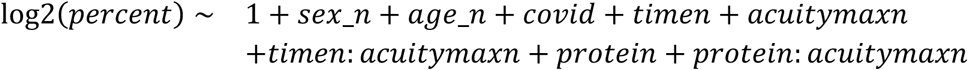

where *protein* represents the concentration (NPX) of a protein in the serum.

For each scRNA-seq gene and each OLINK protein, we fit a linear model with limma to find proteins associated with gene expression in each cell lineage:

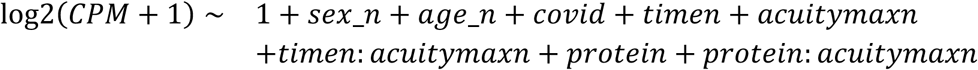

### Antibodies and autoantibodies

We measured 76 antibodies and autoantibodies in serum from each patient sample (see **Autoantibody methodology** above). For each antibody, we fit linear models to find antibodies associated with scRNA-seq cell cluster abundance:

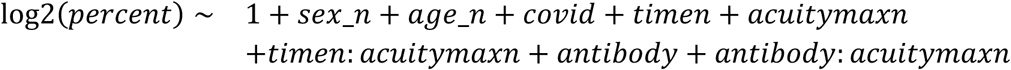

where *antibody* represents the log2 median fluorescence intensity (MFI) of an antibody in the serum.

We also fit linear models to find antibodies associated with scRNA-seq gene expression in each cell lineage:

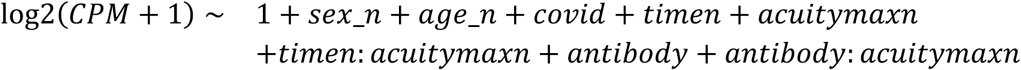

### TCR and BCR clonality analysis

We ran the Cell Ranger 4.0.0 VDJ pipeline on the provided reference files (md5sum 67e4fff7e80d7f7c87ec16475859c080 refdata-cellranger-vdj-GRCh38-alts-ensembl-4.0.0.tar.gz). We only considered cells that are part of our scRNA-seq analysis, and we considered a unique TCR sequence to be the concatenation of four components reported in the all_contig_annotations.csv file from Cell Ranger: TCRɑ V-gene, TCRɑ CDR3 amino acid sequence, TCRβ V-gene, and TCRβ CDR3 amino acid sequence. Clones missing any of these features were not included in the analyses. We considered a unique BCR clone to be the concatenation of two components reported in the all_contig_annotations.csv file from Cell Ranger: heavy-chain CDR3 amino acid sequence and light chain CDR3 amino acid sequence. We only considered clones that contain both a heavy chain CDR3 amino acid sequence and also one of a lambda or a kappa light chain CDR3 amino acid sequence.

For each cell lineage (CD8 T/NK, CD4 T, B cells), we estimated the clonality of TCR or BCR sequences. Then we tested for associations between maximum acuity and clonality, as defined below^117^.

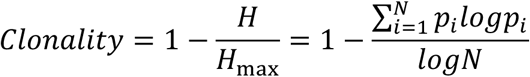

where *N* is the total number of unique TCR or BCR sequences. *H* is the observed Shannon entropy, and *Hmax* is the maximum possible entropy (*log(N)*). *pi* is the relative abundance of a specific TCR or BCR sequence.

We used linear models to test for association between clonality and maximum disease acuity. We fit the following model:

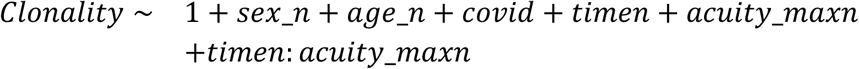

For CD8 T cells, we use *Clonality*. But, for CD4 T cells, we use *log*1*p*(*Clonality*), and for B cells we discard samples with *Clonality* > 1*e* − 5 use *log*10(*Clonality*) instead of *Clonality*, because most of the values are near zero for these cell lineages.

### Calling HLA genotypes from bulk RNA-seq data

We used arcasHLA^118^ with SmartSeq2 bulk RNA-seq reads from neutrophils from the same patient samples^12^ to call HLA genotypes on each patient sample. Then, we discarded the patients where all of their samples (e.g., D0, D3, D7) did not have matching genotypes.

### HLA association with COVID-19 acuity

We used hlabud to convert the HLA genotypes like HLA-A*01:01 to dosage matrices, where each row represents a patient and each column represents an amino acid at a position in the gene. Then, we fit the following model:

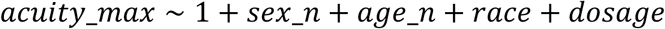

Where *dosage* is a variable with levels [0, 1, 2] representing the number of copies of a particular amino acid at a specific position in an HLA gene.

### Association of cell cluster abundance and gene expression with tocilizumab treatment

We used stats::lm() in R to fit linear models to the log2(percent) of each cell cluster, within each of the four cell lineages (MNP, CD8 T, CD4 T, B):

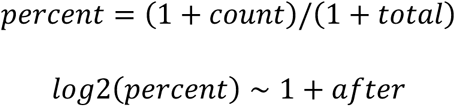

Where *count* is the number of cells in a cluster, *total* is the number of cells in one lineage, for each donor, and *gender* is a binary variable indicating whether the sample was taken before or after tocilizumab treatment.

We used limma to fit linear models to the log2(CPM+1) of each gene in the pseudobulk expression data at the level of cell lineage, and at the level of cell cluster:

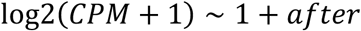

### Association of cell cluster abundance and gene expression with past COVID infection

We used stats::lm() in R to fit linear models to the log2(percent) of each cell cluster, within each of the four cell lineages (MNP, CD8 T, CD4 T, B):

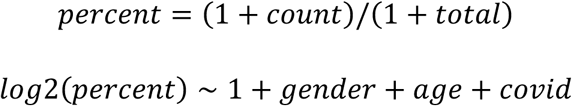

Where *count* is the number of cells in a cluster and *total* is the number of cells in one lineage, for each donor. *gender* indicates male or female gender, *age* indicates the numeric age of the donor, and *covid* is a binary indicator of SARS-CoV-2 infection three months ago.

We used limma to fit linear models to the log2(CPM+1) of each gene in the pseudobulk expression data at the level of cell lineage, and also at the level of cell cluster. This is the main model we used to find genes that are associated with past COVID infection:

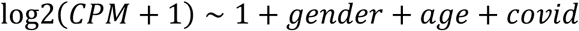

### Classification of cells from tocilizumab data into clusters from acute data

We concatenated the expression matrix of the MNPs from the tocilizumab data and the acute data. Then we used the methods described above to normalize the data, compute PCA scores, and integrate batches with Harmony. We trained a linear discriminant analysis (LDA) model (with 10,000 randomly selected cells from the acute data) to predict the acute cluster labels from the harmonized PCA scores. Then we input the PCA scores for each cell from the tocilizumab data to the LDA model to predict the acute cell cluster label for that cell.

## Lead Contact

Further information and requests for resources and reagents should be directed to and will be fulfilled by the Lead Contact, Alexandra-Chloé Villani.

## Data Availability

Data will be made available upon publication. The scRNA-seq, OLINK, and antibody data and analysis results will be available for browsing on our own interactive website. Analysis results will be archived on Zenodo. Count matrices (scRNA-Seq, TCR, BCR, and CITE-seq) and related data will be deposited to NCBI GEO. Source code for data analysis will be available on GitHub and Zenodo. The raw RNA sequencing data reported in this study cannot be deposited in a public repository because these data were collected at the beginning of the COVID-19 pandemic, and as such, a waiver of informed consent was approved by the Massachusetts General Hospital governing institutional review board, in compliance with the Code of Federal Regulation (45CFR 46, 2018 Common Rule). To protect the identity of individual subjects, public posting of raw sequencing data from the patients has not been approved; therefore, raw data is not provided.

## Acknowledgments

We owe deep gratitude to the study participants, Translational and Clinical Research Center (TCRC), and nursing staff, in particular Grace Holland, RN, Katherine Broderick, RN, and Siobhan Boyce, RN, and Kathryn Hall, NP, for sample collection. We thank the Massachusetts General Hospital for institutional support to enable enrollment when access to clinical spaces was limited. We thank the Departments of Emergency Medicine and Medicine for maintaining needed staffing levels during enrollment, when many research funding sources were suspended. We thank Caroline Beakes and Nicole Russell for assistance with data entry, and Pierre Ankomah for his feedback on data interpretation of myeloid cell subsets. We thank Arthur, Sandra, and Sarah Irving for a gift that enabled this study and funded the David P. Ryan, MD Endowed Chair in Cancer Research (to N.H.).

This manuscript is the result of funding in whole or in part by the National Institutes of Health (NIH). It is subject to the NIH Public Access Policy. Through acceptance of this federal funding, NIH has been given a right to make this manuscript publicly available in PubMed Central upon the Official Date of Publication, as defined by NIH.

We acknowledge the following funding sources: this work was supported by several training grants, including a NIAID grant T32AR007258 (to KS), three NHLBI grants 5T32HL116275-13 (to CC), 5T32HL129970-09 (to APN), and the K08HL157725 (to PS), as well as the American Heart Association Career Development Award (to PS). PS was also supported by the Brigham and Women’s Hospital Innovation Evergreen Fund.

EY was supported by funding from the Stanford Medical Scholars program. RJX acknowledges supports from NIH DK43351 and U19AI142784. RJX and AR were supported by the Manton Foundation and the Klarman Cell Observatory. PJU was supported by Third Rock Ventures; Henry Gustav Floren Trust; Stanford Department of Medicine Team Science Program; Stanford Medicine Office of the Dean; and National Institutes of Health R01 grants AI175771 and AI182319-02. RPB acknowledges funding support from the Massachusetts General Hospital Executive Committee on Research, the American Lung Association, and the Broad Institute’s Next Generation Scholar award. MBG, MRF, and NH were supported by an American Lung Association COVID-19 Action Initiative grant. MBG and MRF were supported by a grant from the Executive Committee on Research at MGH. NH acknowledges was supported by NIH/NIAID U19 AI082630, a Chair and gift from Sandra, Sarah, and Arthur Irving. ACV acknowledges funding support from the COVID-19 Clinical Trials Pilot grant from the Executive Committee on Research at MGH; a COVID-19 Chan Zuckerberg Initiative grant (2020-216954); the funds from the Manton Foundation and the Klarman Family Foundation; the Broad Institute’s Next Generation Scholar award; the MGH Howard M. Goodman Fellowship; the National Institutes of Health (DP2CA247831); work at the Broad Institute was supported by a gift from an anonymous donor. The funders had no role in study design, data collection and analysis, decision to publish, or preparation of the manuscript.

## Author Contributions

MBG, MRF, MSF, NH and ACV conceived and led the overall study design. PS, MKM, and ACV conceived and led the tocilizumab scRNA-seq study design. EMB, LC, AEW, OR, AR, DH, JD and RJX conceived the convalescence study cohort while ACV and PS conceived and led the convalescence scRNA-seq study design. PJU, NH, and ACV conceived the autoantibody study design. MRF led and oversaw the recruitment of the MGH acute COVID-19 cohort, with clinical additional clinical expertise provided by RPB and MBG. KLP, BL, BM, CL, HK, KK, NC recruited and consented patients from the MGH acute COVID-19 cohort. MSF and ACV oversaw the processing of the MGH PBMC and serum specimens for the acute cohort, with assistance from PCB, GMB, MBG, MRF, and NH. JS, TJL, KM, AG, IG, BR, MRL, NS, MF processed PBMC and serum samples from the acute cohort with assistance from TS, and DF; MB, LC, and AEW established the convalescent clinical cohort. LAZ, SMP, BKT, VP, CH, NP and DP processed PBMC and serum samples from the convalescent cohort, PS led the scRNA-seq experimental design and execution. PS, JT, TE, TJL, KM, AT, BYA, MR performed the scRNA-seq experiments, with assistance from DP and TD for the convalescent cohort. APN and EYAY generated the autoantibody data. KS designed and performed computational analysis with assistance from CC, NS, SR; KS, PS, CC, and ACV led the data interpretation with assistance from LB, PCB, RPB, PJU, MBG, MJM, MRF, MSF, and NH; ACV managed and supervised the entire study. KS, PS, CC, ACV wrote the manuscript with input from all authors.

## Supplementary Figures

**Figure S1.**
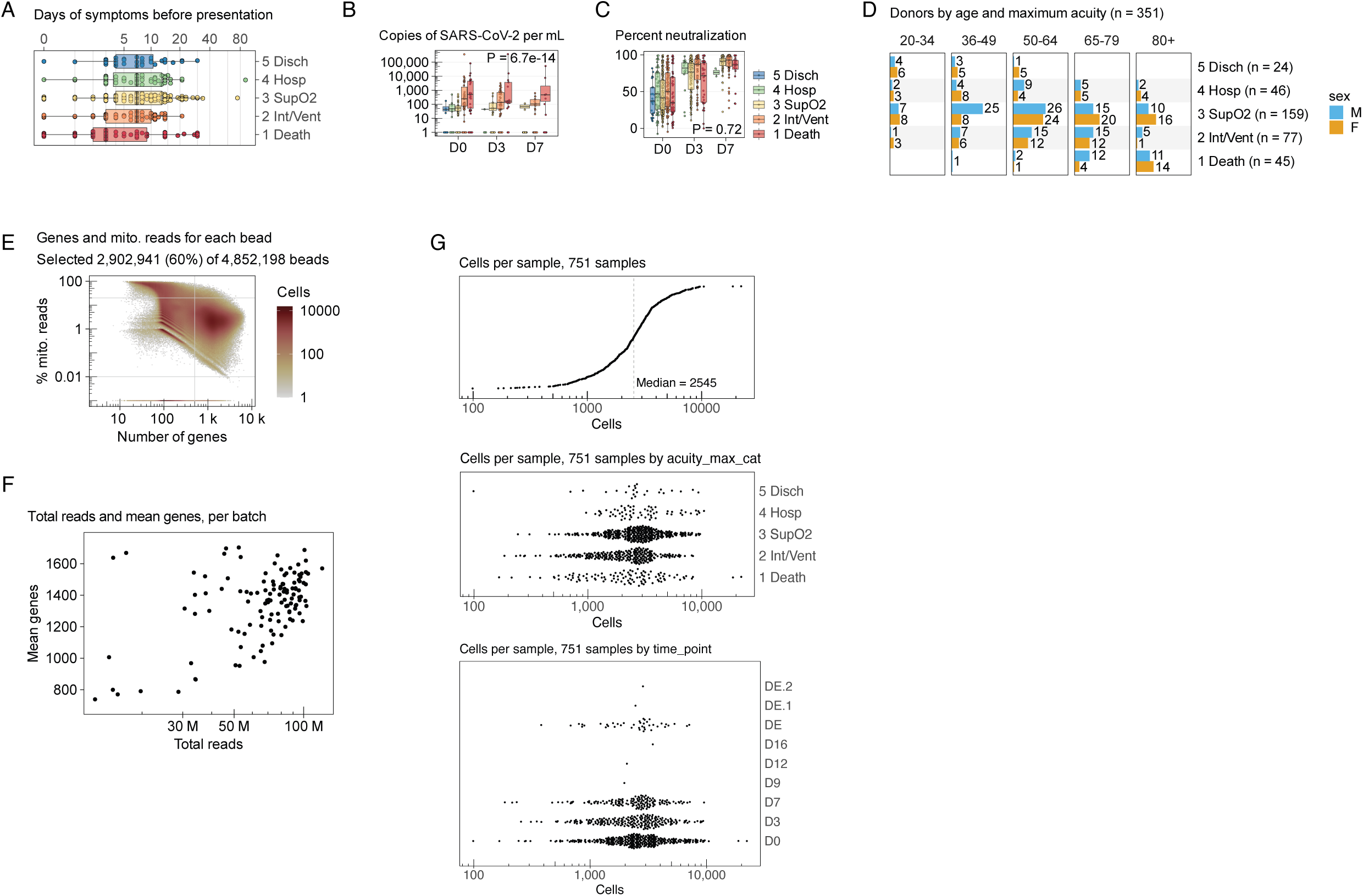
Quality control. **A.** Days of symptoms before presentation, displayed as a box plot (top) and histogram faceted by COVID severity (bottom). **B.** Copies of SARS-CoV-2 per mL of blood, by time (x-axis) and COVID severity (hue). P-value indicates association with COVID severity. **C.** Percent neutralization from an *in vitro* assay, by time (x-axis) and COVID severity (hue). P-value indicates association with COVID severity. **D.** Bar plot of donors by age and maximum COVID severity, faceted by age, hue indicates sex. **E.** Hexbin plot of the number of genes and percent of mitochondrial reads for each bead. X-axis indicates the number of genes detected in each bead. Y-axis indicates the percentage of reads assigned to 13 mitochondrial genes in each bead. Hue indicates the number of cells inside the hexagon. Horizontal and vertical lines show chosen cutoffs: 500 genes, 0.01% and 20% mitochondrial reads. **F.** Scatter plot with one dot for each batch. X-axis indicates the total number of sequencing reads in the batch. Y-axis indicates the mean genes per cell in the batch. **G.** Distribution of cells per sample. Top panel shows a median of 2,545 cells per sample. Middle panel shows cells per sample stratified by COVID-19 severity level. Bottom panel shows cells per sample stratified by time point.

**Figure S2.**
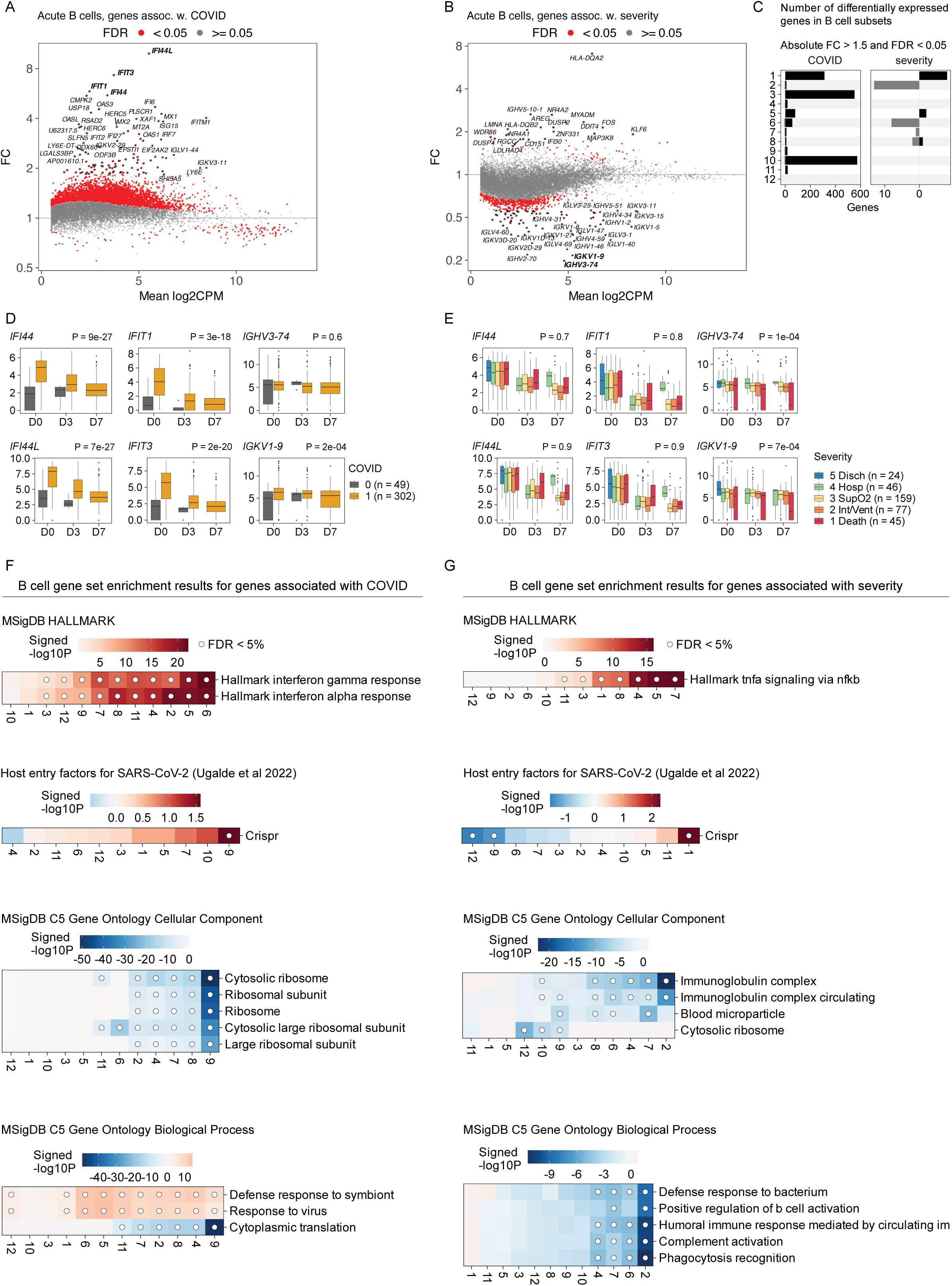
Differential expression for B cells from the acute data. **A.** MA plot for genes associated with COVID infection. **B.** MA plot for genes associated with COVID severity. **C.** Bar plot showing the number of differentially expressed genes for each cell subset, associated with COVID infection (left) or COVID severity (right). **D.** Box plots of two genes (*IFI44* and *IFIT1*) by time point (x-axis) and COVID infection (hue). **E.** Box plots of two genes (*IFI44* and *IFIT1*) by time point (x-axis) and COVID severity (hue). **F, G.** COVID infection (**F**) and severity (**G**) associated gene sets for each cell subset (columns). Hue indicates the signed log10 enrichment P-value from fgsea(), dot indicates FDR < 5%.

**Figure S3.**
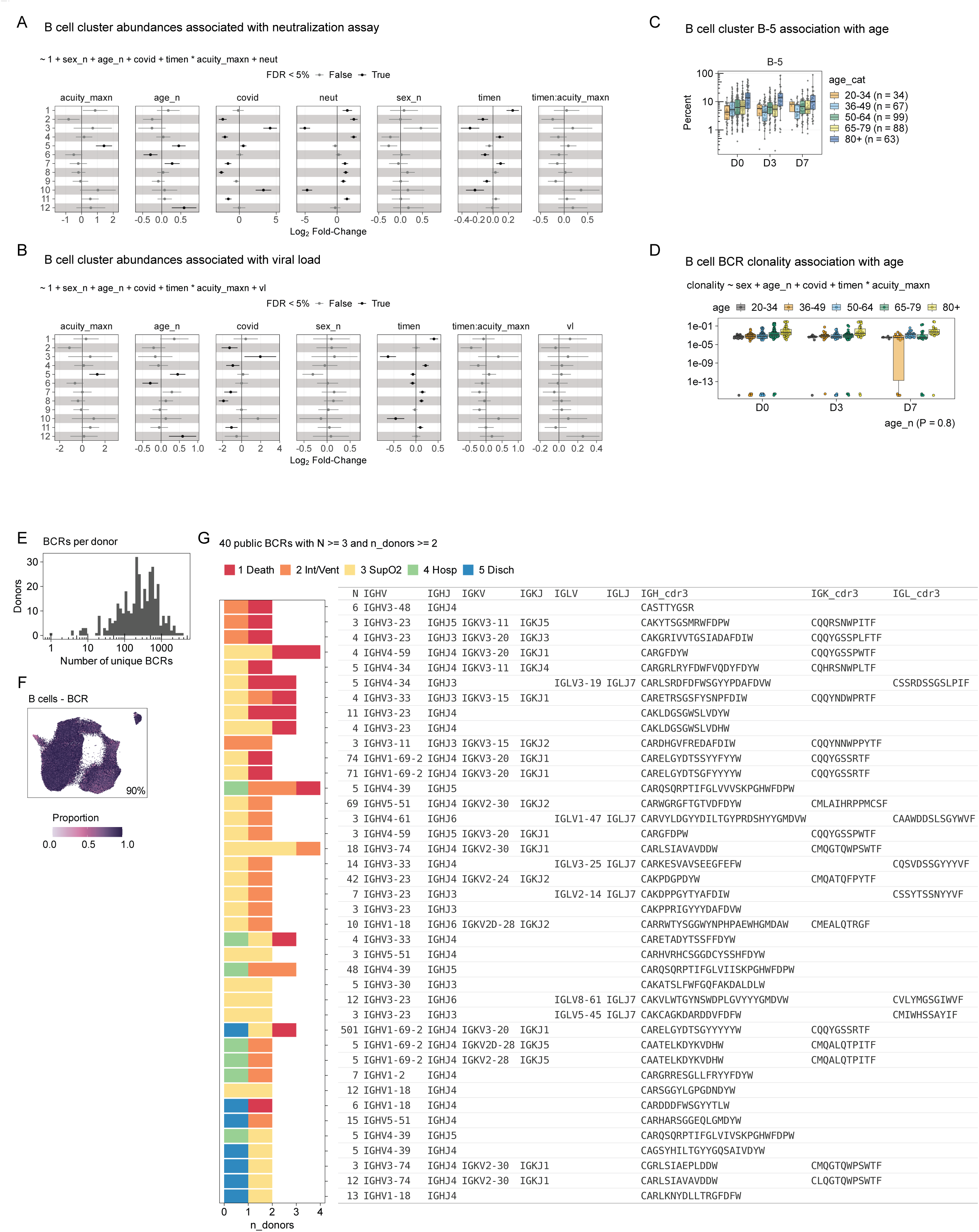
BCR analysis for B cells from acute data. **A.** Error bars representing the estimates and 95% CI of coefficients from the linear model log2(pct) ∼ 1 + sex + age_n + covid + timen + acuity_maxn + timen:acuity_max + neut Columns indicate coefficients (age, time, sex, covid, interaction between time and severity, severity, neutralization). Rows indicate the B cell subsets. **B.** Error bars representing the estimates and 95% CI of coefficients from the linear model log2(pct) ∼ 1 + sex + age_n + covid + timen + acuity_maxn + timen:acuity_max + vl (bottom panel). Columns indicate coefficients (age, time, sex, covid, interaction between time and severity, severity, viral load). Rows indicate the B cell subsets. **C.** Box plot of B cell subset B-5 abundance, by age. Dots represent patients, box plots indicate median and interquartile range. X-axis and hue indicate age of patients, y-axis indicates abundance of B-5 in each sample. **D.** Box plot of B cell BCR clonality, by age. Dots represent patients, box plots indicate median and interquartile range. X-axis and hue indicate age of patients, y-axis indicates clonality of TCRs in each sample. P-value indicates association with age. **E.** Histogram of the number of unique BCRs per donor in B cells from the acute data. **F.** UMAP embedding, where the color of each hexagon indicates the proportion of cells in that hexagon that have BCR data. The percent of cells with BCR data is shown. **G.** Table of 40 public BCRs that are observed at least 3 times in the dataset and observed in at least 2 donors. Bars indicate the number of donors that have each BCR, and hue indicates the maximum COVID severity for each donor.

**Figure S4.**
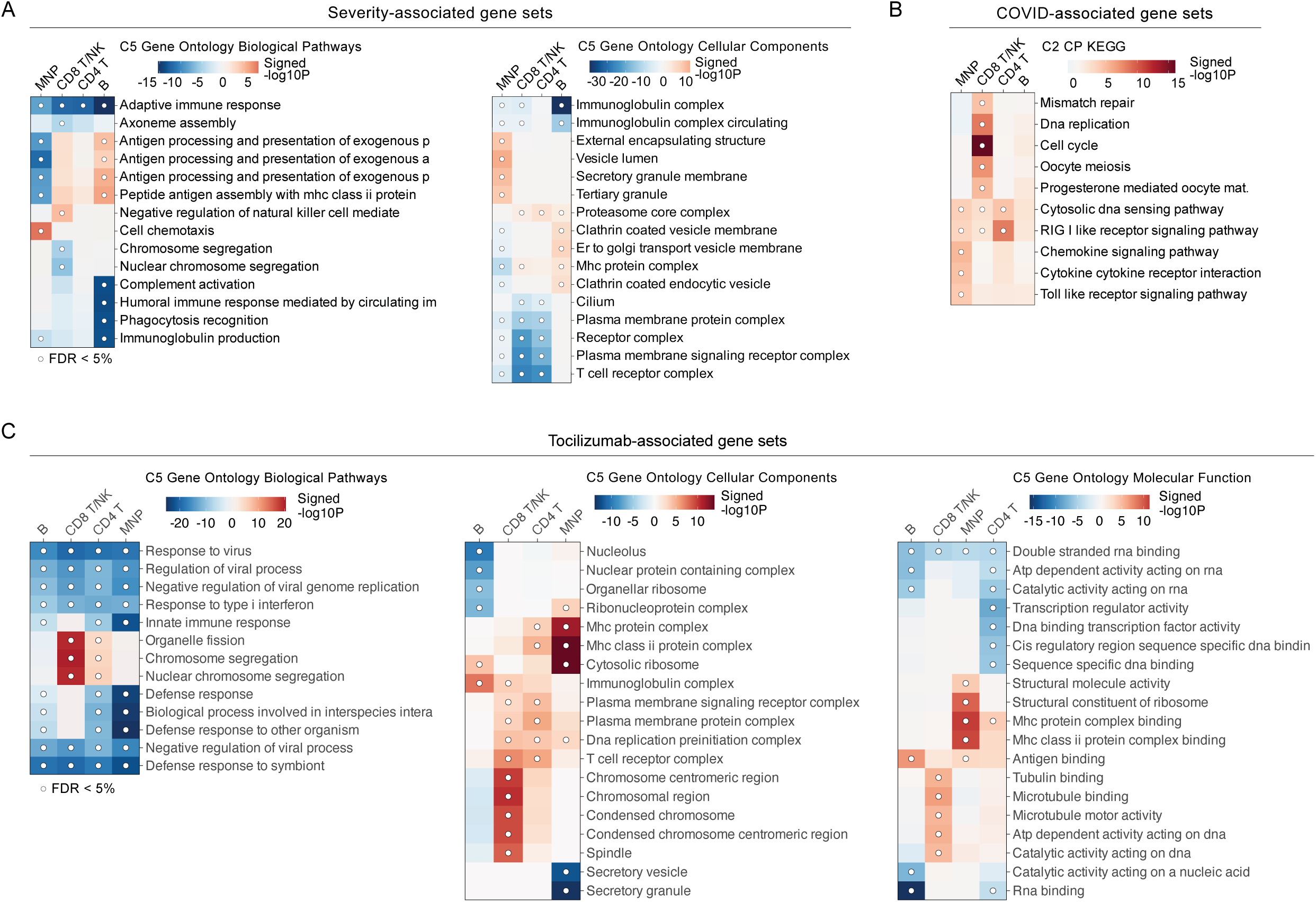
Gene set enrichment for acute and tocilizumab data. **A.** COVID severity associated gene sets from the C5 Gene Ontology Biological Process collection (left), C5 Gene Ontology Cellular Components collection (right) for each cell lineage. Hue indicates the signed log10 enrichment P-value from fgsea(), dot indicates FDR < 5%. **B.** COVID infection-associated gene sets from the C2 CP KEGG collection (**B**) for each cell lineage (columns). Hue indicates the signed log10 enrichment P-value from fgsea(), dot indicates FDR < 5%. **C.** Associations with tocilizumab treatment for Gene Ontology Biological Process (left), Cellular Component (middle), and Molecular Function (right). Hue indicates the signed log10 enrichment P-value from fgsea(), dot indicates FDR < 5%.

**Figure S5.**
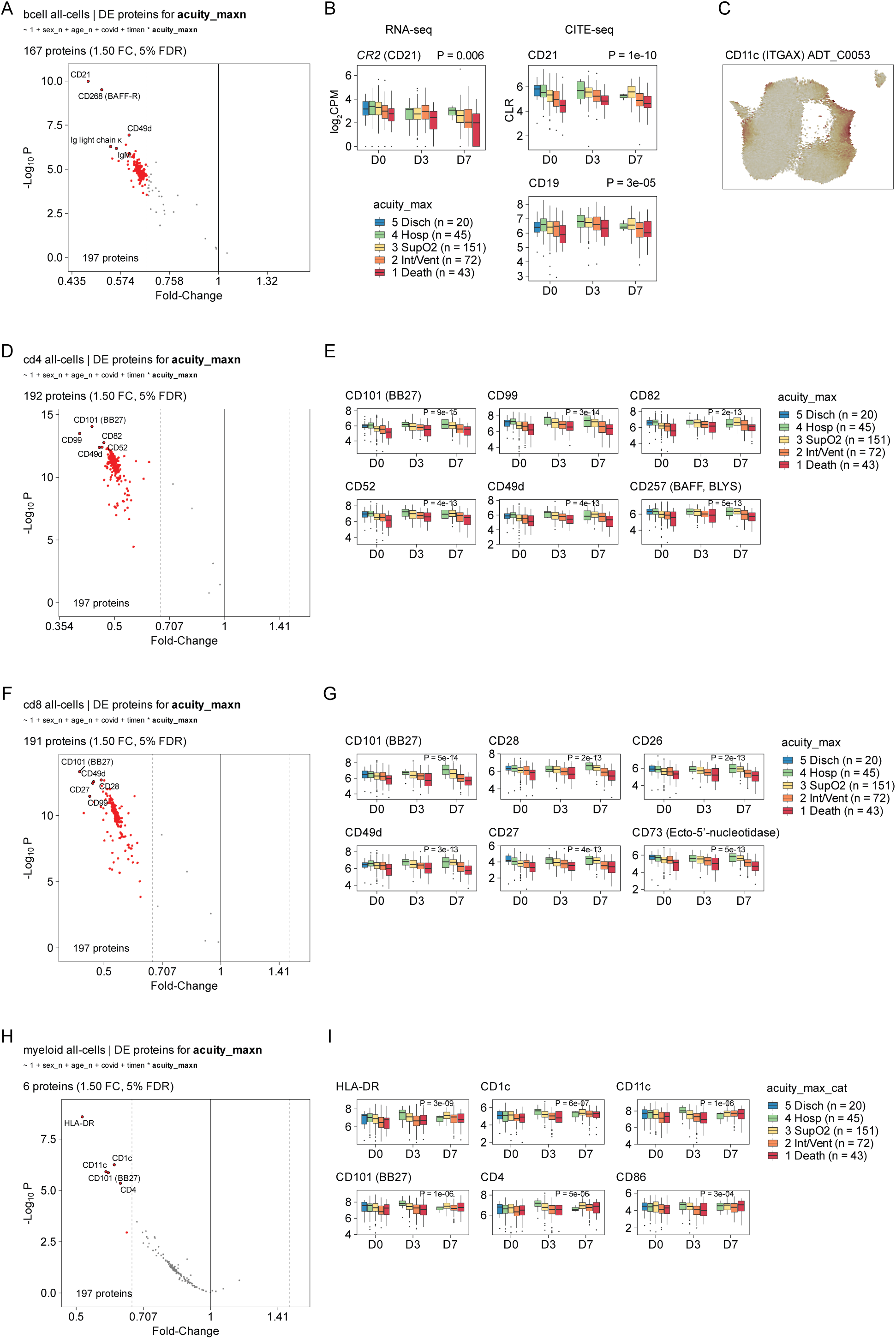
CITE-seq protein associations with COVID-19 severity. **A, D, F, H.** Volcano plot showing the log2 fold-change (x-axis) and negative log10 p-value (y-axis) for the association of 197 CITE-seq proteins with COVID-19 severity. **B, E, G, I.** Box plots of **CR2** RNA-seq gene expression and CD21 CITE-seq protein level. Dots represent patients, box plots indicate median and interquartile range, x-axis represents three time points (D0, D3, D7). Hue indicates COVID severity. **C.** UMAP embedding of B cells from the acute data. Hue indicates CITE-seq protein level of CD11c (ITGAX).

**Figure S6.**
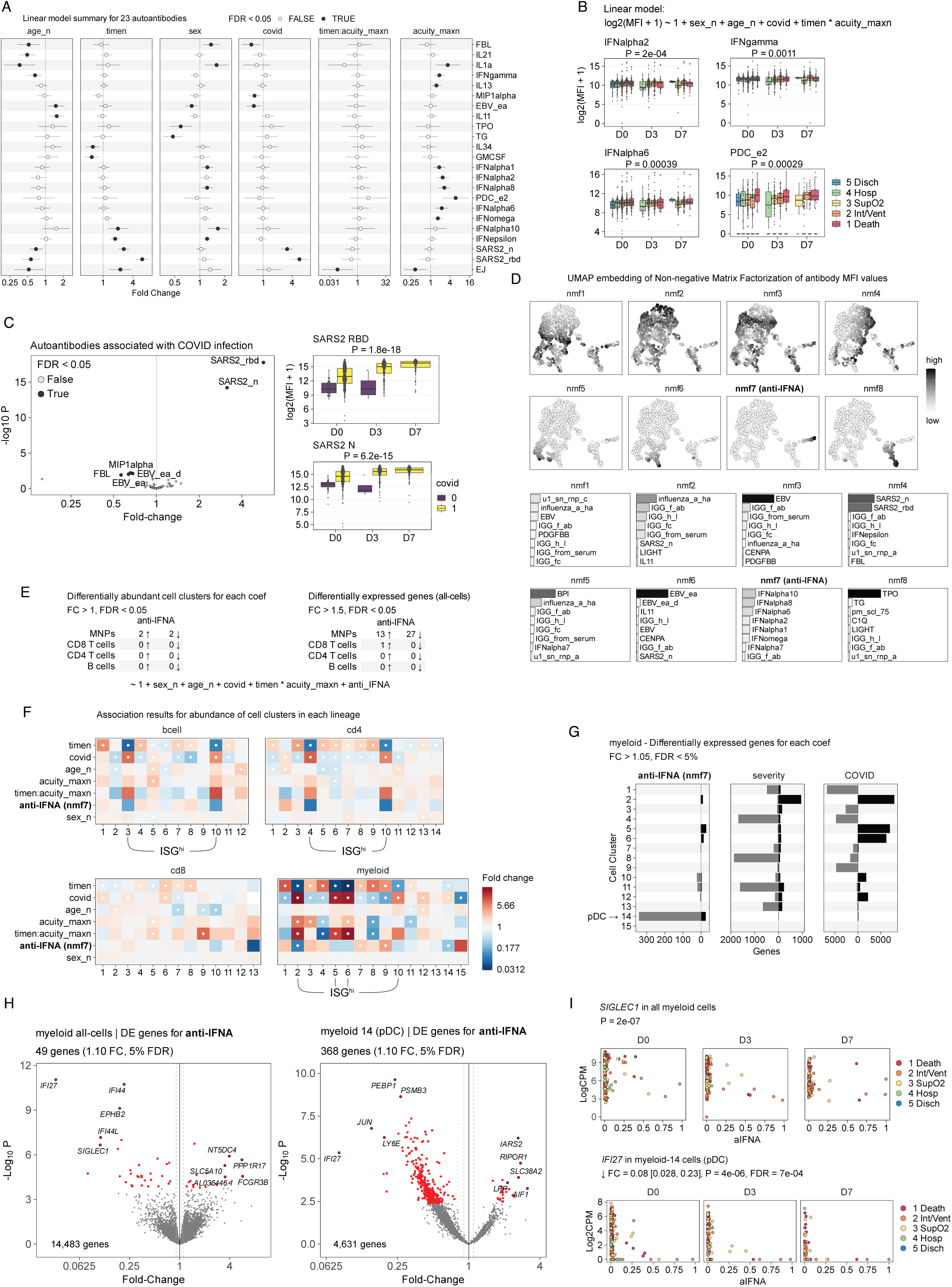
Autoantibodies associated with COVID infection and severity. **A.** Error bars representing the estimates and 95% CI of coefficients from the linear model log2(MFI+1) ∼ 1 + sex + age_n + covid + timen + acuity_maxn + timen:acuity_max. Columns indicate coefficients (age, time, sex, covid, interaction between time and severity, and severity). Rows indicate the antibody targets. **B.** Box plots of antibodies associated with COVID severity. x-axis represents time points and y-axis represents the log2(MFI+1) measurement of the antibody. Hue indicates COVID infection status. **C.** 74 autoantibodies associated with SARS-CoV-2 infection, x-axis represents fold-change and y-axis represents negative log10 p-value. Two box plots show the measurements of antibodies against SARS2_RBD and SARS2_N, x-axis represents time points and y-axis represents the log2(MFI+1) measurement of the antibody (median fluorescence intensity, or MFI, see **Methods** for details). Hue indicates COVID infection status. **D.** UMAP embedding of NMF factorization of the full matrix of 74 autoantibodies across all samples in the acute dataset. Hue indicates the normalized value of each NMF component. Bottom panels represent loading values for top 8 antibodies for each of the 8 NMF components, x-axis and hue both indicate the loading value. **E.** Within each cell lineage, the number of differentially abundant cell subsets (left) and number of differentially expressed genes (right) that are associated with NMF component 7 (nmf7, labeled “aa”). For example, 2 cell subsets in the MNP lineage are more abundant in proportion to the aa variable, and 2 cell subsets are less abundant in proportion to the aa variable. **F.** Heatmaps showing the fold-changes for associations of each variable (row) with each cell subset (column), in each of the 4 cell lineages (bcell, cd4, cd8, myeloid). Dots indicate FDR < 5%. Hue indicates fold-change. The rows indicate the coefficients from the same linear model shown in panel **B**. **G.** Number of differentially expressed genes in the MNP (myeloid) lineage for each cell subset (row) and each coefficient (aa (nmf7), acuity_maxn, covid). **H.** Volcano plot of differentially expressed genes associated with aa (nmf7) in the myeloid cell lineage, x-axis shows fold-change and y-axis shows negative log10 p-value. Left panel shows results for pseudobulk expression in all myeloid cells, right panel shows results for pseudobulk expression in myeloid cell subset 14 (pDC). **I.** Scatter plots showing the relationship between gene expression (y-axis) of *SIGLEC1* (top panels) and *IFI44L* (bottom panels) versus the value of aa (x-axis). Each dot represents a sample, hue indicates COVID severity. P-value indicates association of gene expression with aa in the linear model from panel **B**.

**Figure S7.**
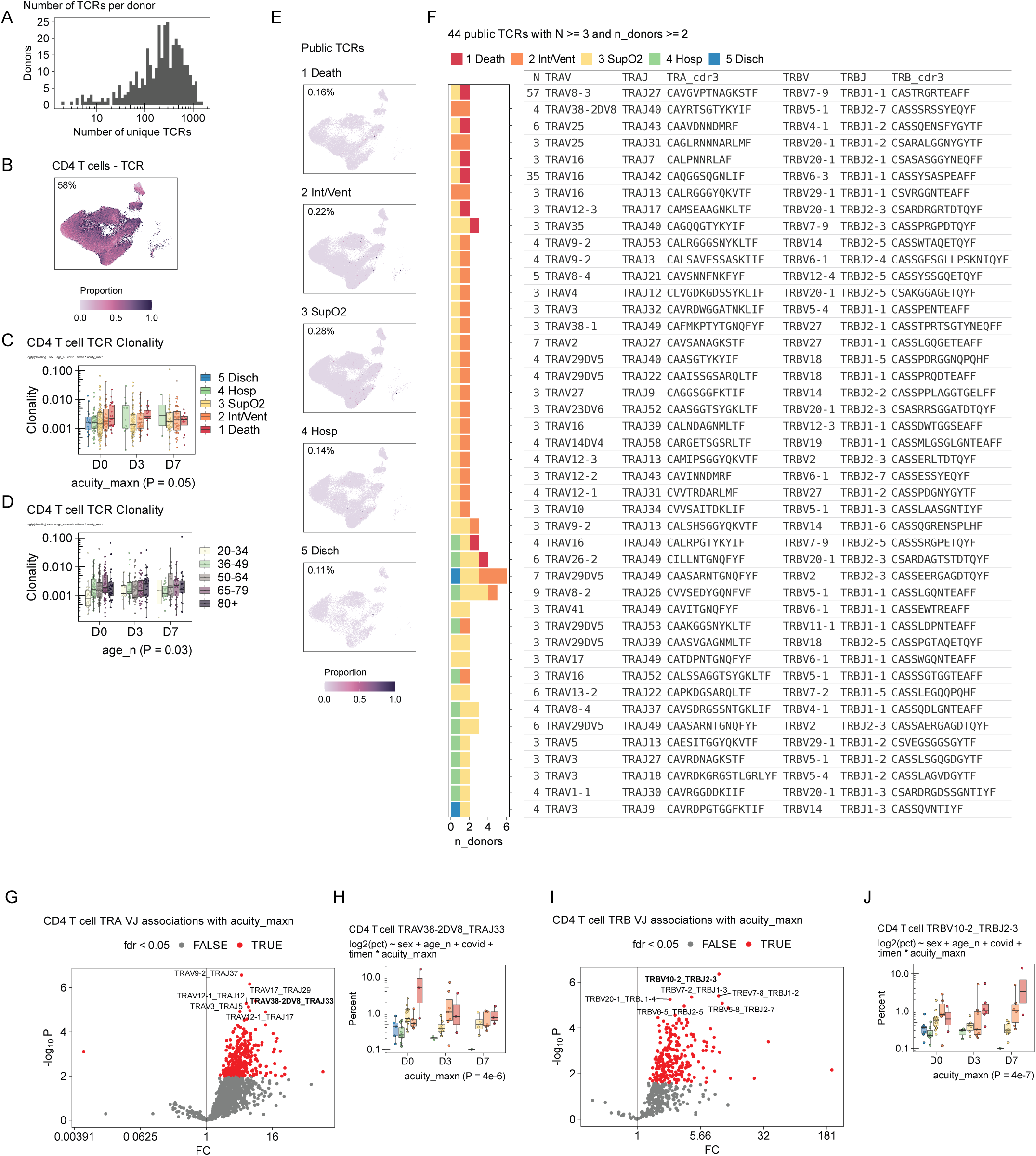
TCR analysis for CD4 T cells from acute data. **A.** Histogram of the number of unique TCRs per donor in CD4 T cells from the acute data. **B.** UMAP embedding, where the color of each hexagon indicates the proportion of cells in that hexagon that have TCR data. The percent of cells with TCR data is shown. **C.** Box plot of CD4 T cell TCR clonality, by COVID severity. Dots represent patients, box plots indicate median and interquartile range. X-axis and hue indicate time point, y-axis indicates clonality of TCRs in each sample. P-value indicates association with severity. **D.** Box plot of CD4 T cell TCR clonality, by age. Dots represent patients, box plots indicate median and interquartile range. X-axis and hue indicate time point, y-axis indicates clonality of TCRs in each sample. P-value indicates association with age. **E.** UMAP embedding, where the color of each hexagon indicates the proportion of cells in that hexagon that have a public TCR. Facets are by COVID severity. Percent of cells with public TCRs are shown. **F.** Table of public TCRs that are observed at least 3 times in the dataset, and observed in at least 4 donors. Bars indicate the number of donors that have each TCR, and hue indicates the maximum COVID severity for each donor. **G, I.** Volcano plot showing analysis results for TRAV and TRAJ association with COVID severity or TRBV and TRBJ association, where each dot represents one combination of V and J genes for the TCR alpha or beta chain. x-axis indicates fold-change and y-axis indicates negative log10 p-value. **H, J.** Box plots showing the abundance of cells with a particular VJ combination associated with severity. Dots represent patients, box plots indicate median and interquartile range. Hue indicates COVID severity; y-axis indicates percent of CD4 T cells with the specified combination of V and J genes. P-value for the severity coefficient from the multivariate linear model are shown.

**Figure S8.**
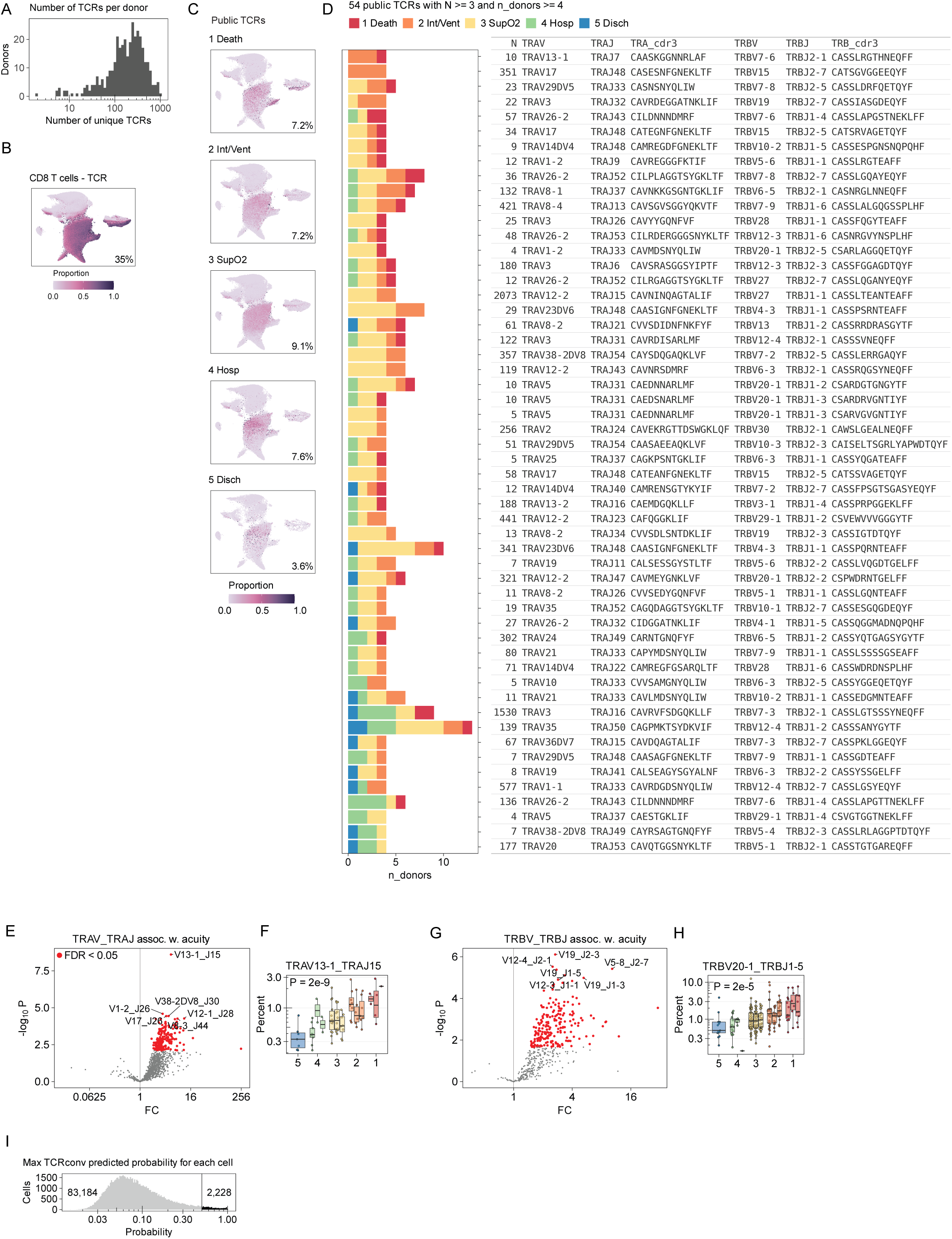
TCR analysis for CD8 T cells from acute data. **A.** Histogram of the number of unique TCRs per donor in CD8 T cells from the acute data. **B.** UMAP embedding, where the color of each hexagon indicates the proportion of cells in that hexagon that have TCR data. The percent of cells with TCR data is shown. **C.** UMAP embedding, where the color of each hexagon indicates the proportion of cells in that hexagon that have a public TCR. Facets are by COVID severity. Percent of cells with public TCRs are shown. **D.** Table of public TCRs that are observed at least 3 times in the dataset, and observed in at least 4 donors. Bars indicate the number of donors that have each TCR, and hue indicates the maximum COVID severity for each donor. **E, G.** Volcano plot showing analysis results for TRAV and TRAJ association (**E**) with COVID severity or TRBV and TRBJ association (**G**), where each dot represents one combination of V and J genes for the TCR alpha or beta chain. x-axis indicates fold-change and y-axis indicates negative log10 p-value. **F, H.** Box plots showing the abundance of cells with a particular VJ combination associated with severity. Dots represent patients, box plots indicate median and interquartile range. Hue indicates COVID severity; y-axis indicates percent of CD8 T cells with the specified combination of V and J genes. The x-axis indicates COVID severity, and each box represents one time point (D0, D3, D7). P-value for the severity coefficient from the multivariate linear model are shown. **I.** Histogram of TCRconv predicted probabilities (or scores) for each of the CD8 T cell TCRs in our data, generated solely from the TCR beta chain. Vertical line at 0.5 indicates the cutoff for high confidence predictions. Number of total cells is shown, as well as the number of cells that pass the cutoff.

**Figure S9.**
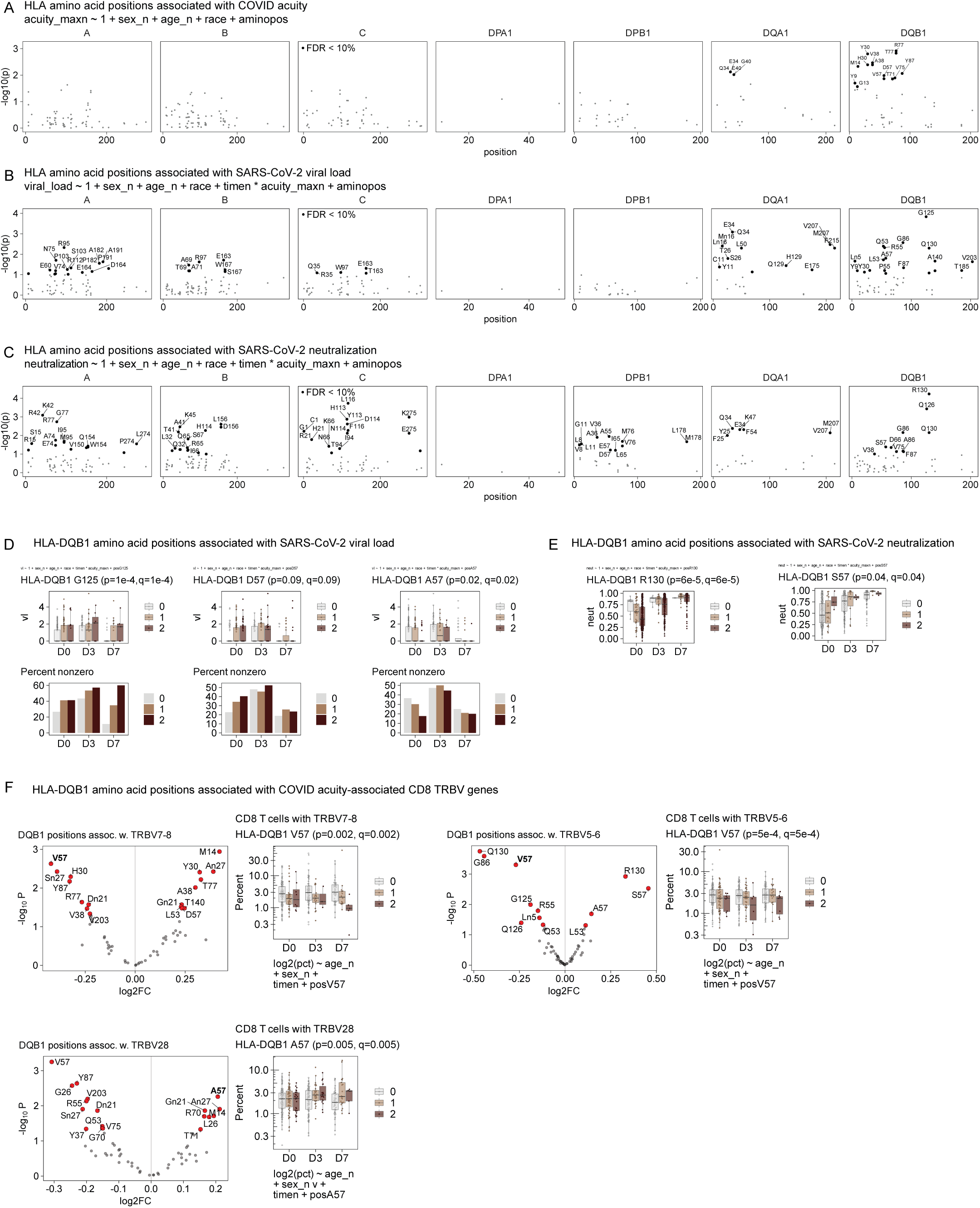
HLA amino acid positions associated with severity, viral load, neutralization, and abundance of CD8 T cells with TRBV genes. **A, B, C.** Associations of HLA amino acid positions with COVID severity (**A**), SARS-CoV-2 viral load (**B**), and *in vitro* SARS-CoV-2 neutralization assay (**C**). X-axis represents position along each protein and y-axis represents negative log10 p-value. One panel per gene. Black dots indicate positions with FDR < 10%, labeled by the amino acid at that position. **D.** Association of HLA-DQB1 G125, D57, and A57 with viral load, x-axis indicates time, and color indicates number of copies of the indicated allele. In top panels, dots represent samples, box plots indicate median and interquartile range. In bottom panels, each bar represents the percent of samples with nonzero values for viral load. **E.** Association of HLA-DQB1 R130 and S57 with SARS-CoV-2 neutralization. X-axis indicates time, and color indicates number of copies of the indicated allele. Dots represent samples, box plots indicate median and interquartile range. **F.** HLA-DQB1 amino acid positions associated with the abundance of CD8 T cells with different TRBV genes. Volcano plots show amino acid positions, x-axis indicates log2 fold-change of CD8 T cell abundance and y-axis indicates negative log10 p-value. Box plots show abundance of CD8 T cells with the indicated TRBV gene on the y-axis, with median and interquartile range, color indicates number of copies of the amino acid position (0, 1, 2), x-axis indicates time point.

**Figure S10.**
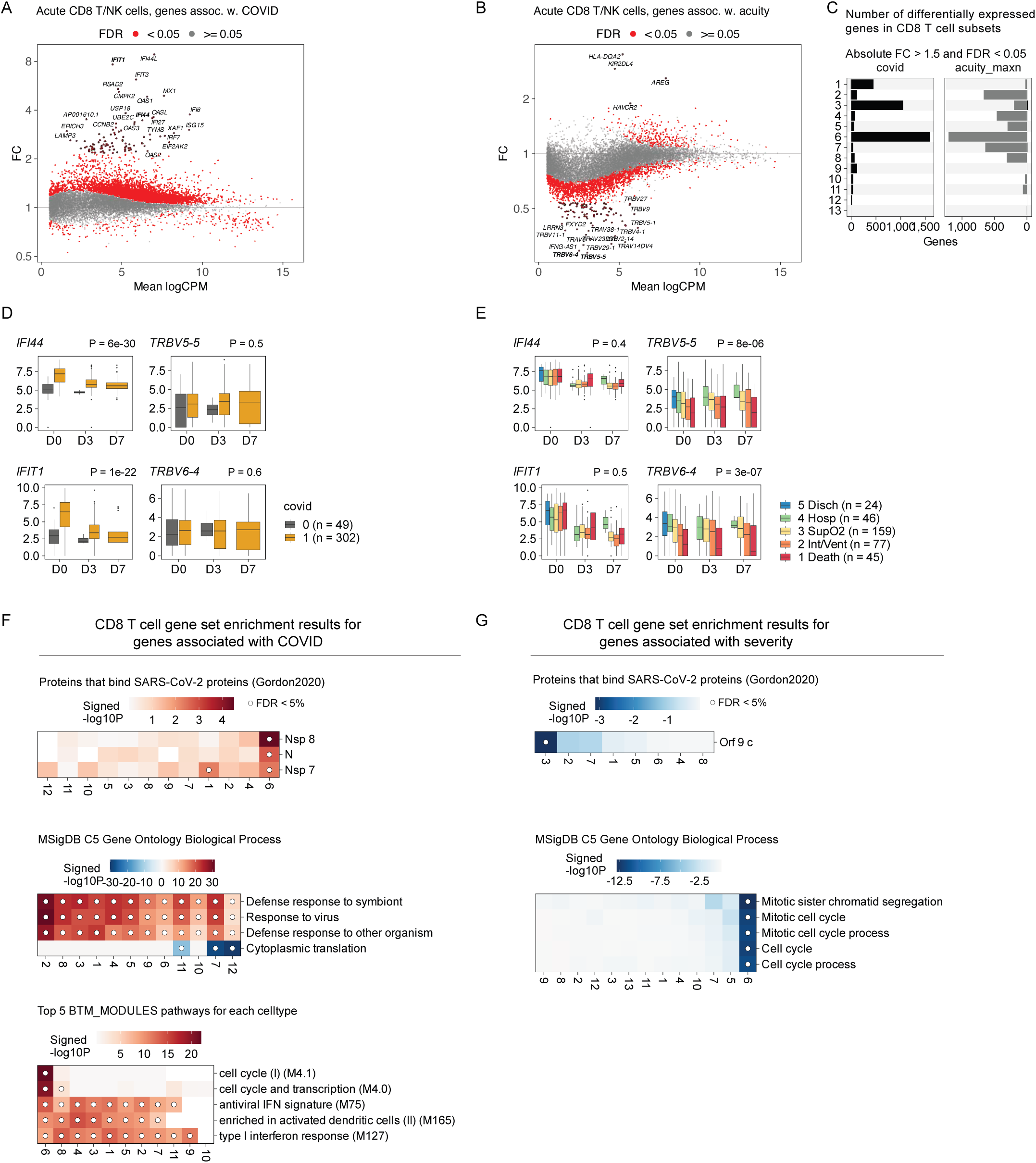
Differential gene expression for CD8 T/NK cells in the acute data. **A.** MA plot for genes associated with COVID infection. **B.** MA plot for genes associated with COVID severity. **C.** Bar plot showing the number of differentially expressed genes for each cell subset, associated with COVID infection (left) or COVID severity (right). **D.** Box plots of two genes (*IFI44* and *IFIT1*) by time point (x-axis) and COVID infection (hue). **E.** Box plots of two genes (*IFI44* and *IFIT1*) by time point (x-axis) and COVID severity (hue). **F, G.** COVID infection (**F**) and severity (**G**) associated gene sets for each cell subset (columns). Hue indicates the signed log10 enrichment P-value from fgsea(), dot indicates FDR < 5%.

**Figure S11.**
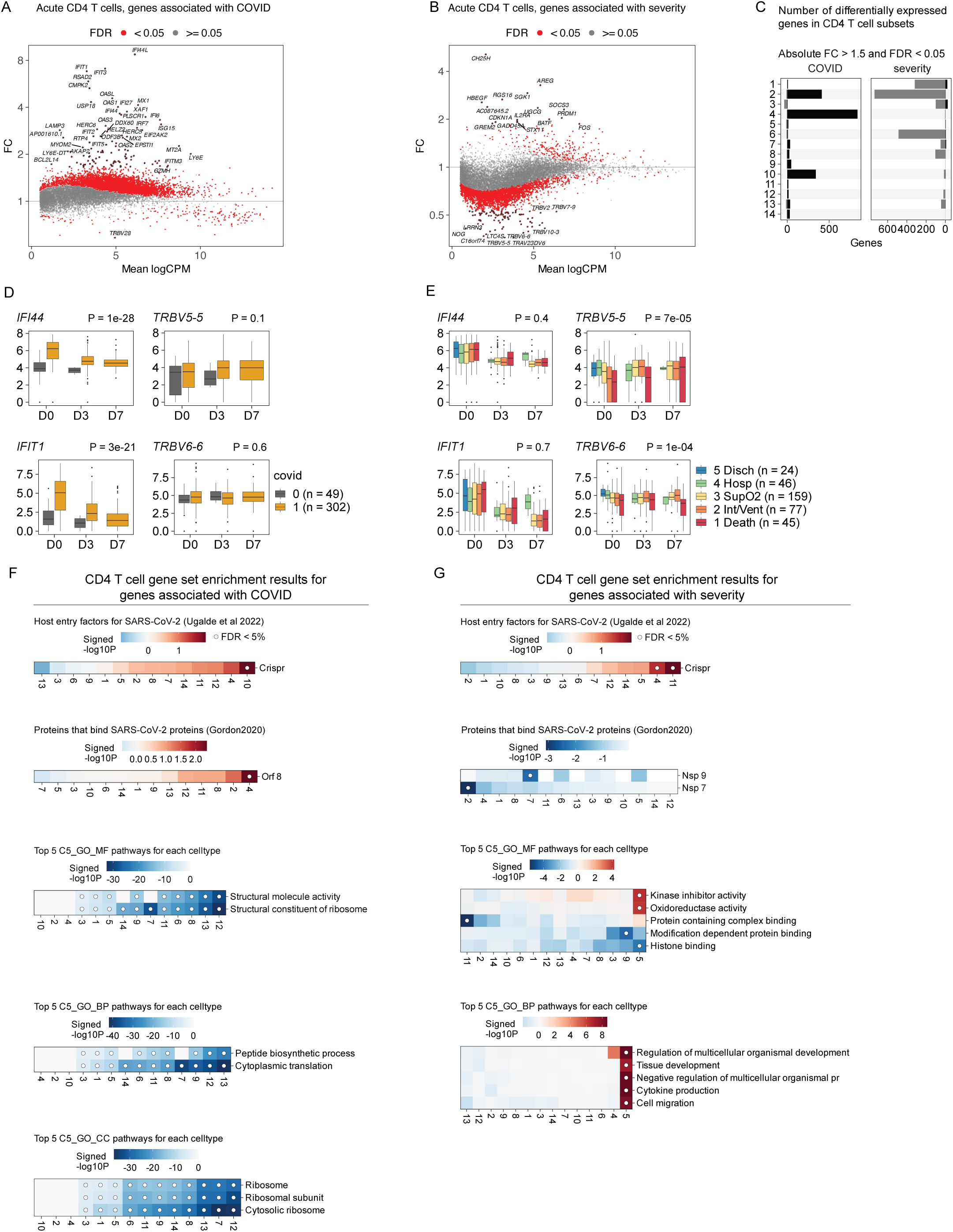
Differential expression for CD4 T cells from acute data. **A.** MA plot for genes associated with COVID infection. **B.** MA plot for genes associated with COVID severity. **C.** Bar plot showing the number of differentially expressed genes for each cell subset, associated with COVID infection (left) or COVID severity (right). **D.** Box plots of two genes (*IFI44* and *IFIT1*) by time point (x-axis) and COVID infection (hue). **E.** Box plots of two genes (*IFI44* and *IFIT1*) by time point (x-axis) and COVID severity (hue). **F, G.** COVID infection (**F**) and severity (**G**) associated gene sets for each cell subset (columns). Hue indicates the signed log10 enrichment P-value from fgsea(), dot indicates FDR < 5%.

**Figure S12.**
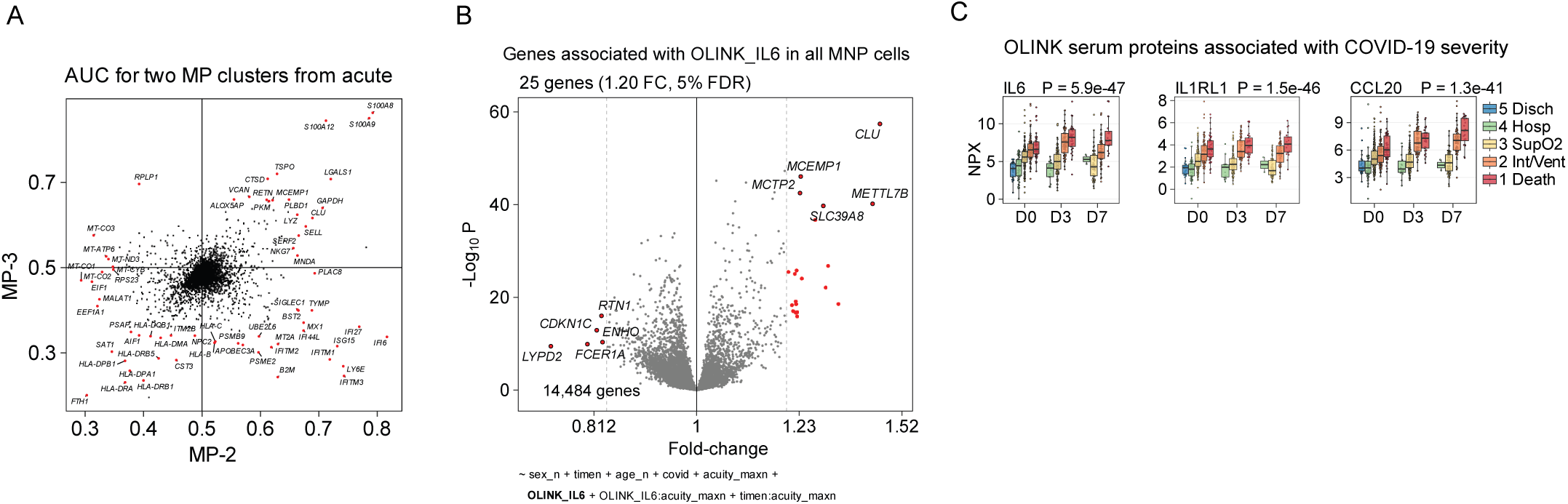
Comparison of myeloid cell subsets from the acute data. **A.** Scatter plot of genes. X-axis represents the area under receiver operator curve (AUC) for gene markers of cell subset MP-2, and y-axis represents AUC for gene markers of cell subset MP-3. **B.** Volcano plot showing the fold-change (x-axis) and negative log10 p-value (y-axis) for the association of gene expression in all myeloid cells versus the serum protein level of IL6. **C.** Box plots showing examples of COVID severity-associated serum proteins. X-axis indicates time point (D3, D0, D7). Y-axis indicates NPX protein level. Each dot represents one patient sample. Box plots show median and interquartile range. Hue indicates disease severity. Multivariate linear regression, two-sided p-value for the coefficient for severity.

**Figure S13.**
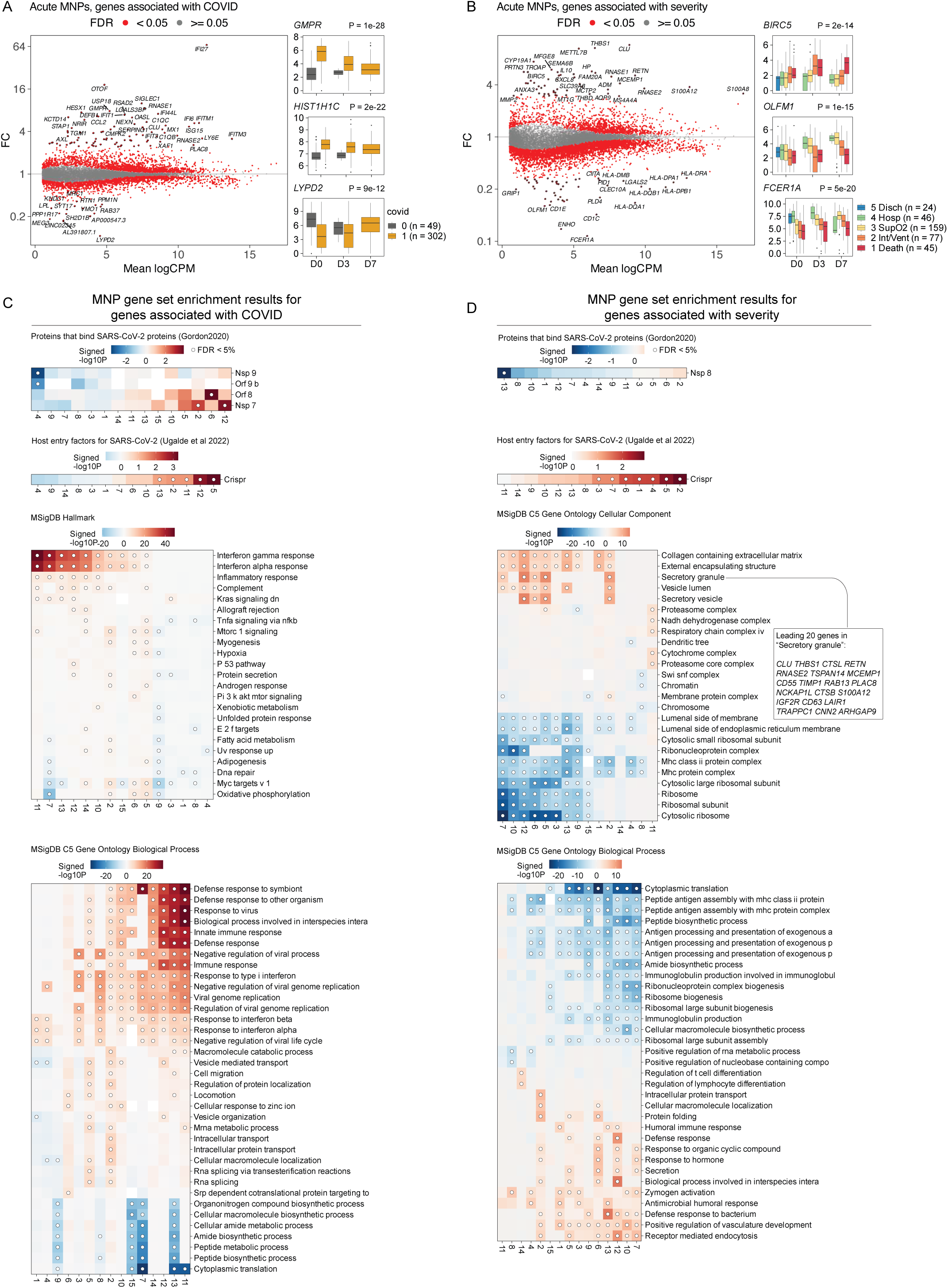
Differential gene expression for MNP cells in the acute data. **A.** MA plot for COVID infection, where each point represents a gene, x-axis represents mean expression (log2CPM) and y-axis represents fold-change for COVID infection. Three box plots represent gene expression of *GMPR*, *HIST1H1C*, *LYPD2* across time points (x-axis) and COVID infection status (hue). **B.** MA plot for genes associated with COVID severity. **C, D.** COVID infection (**C**) and severity (**D**) associated gene sets for each cell subset (columns). Hue indicates the signed log10 enrichment P-value from fgsea(), dot indicates FDR < 5%.

**Figure S14.**
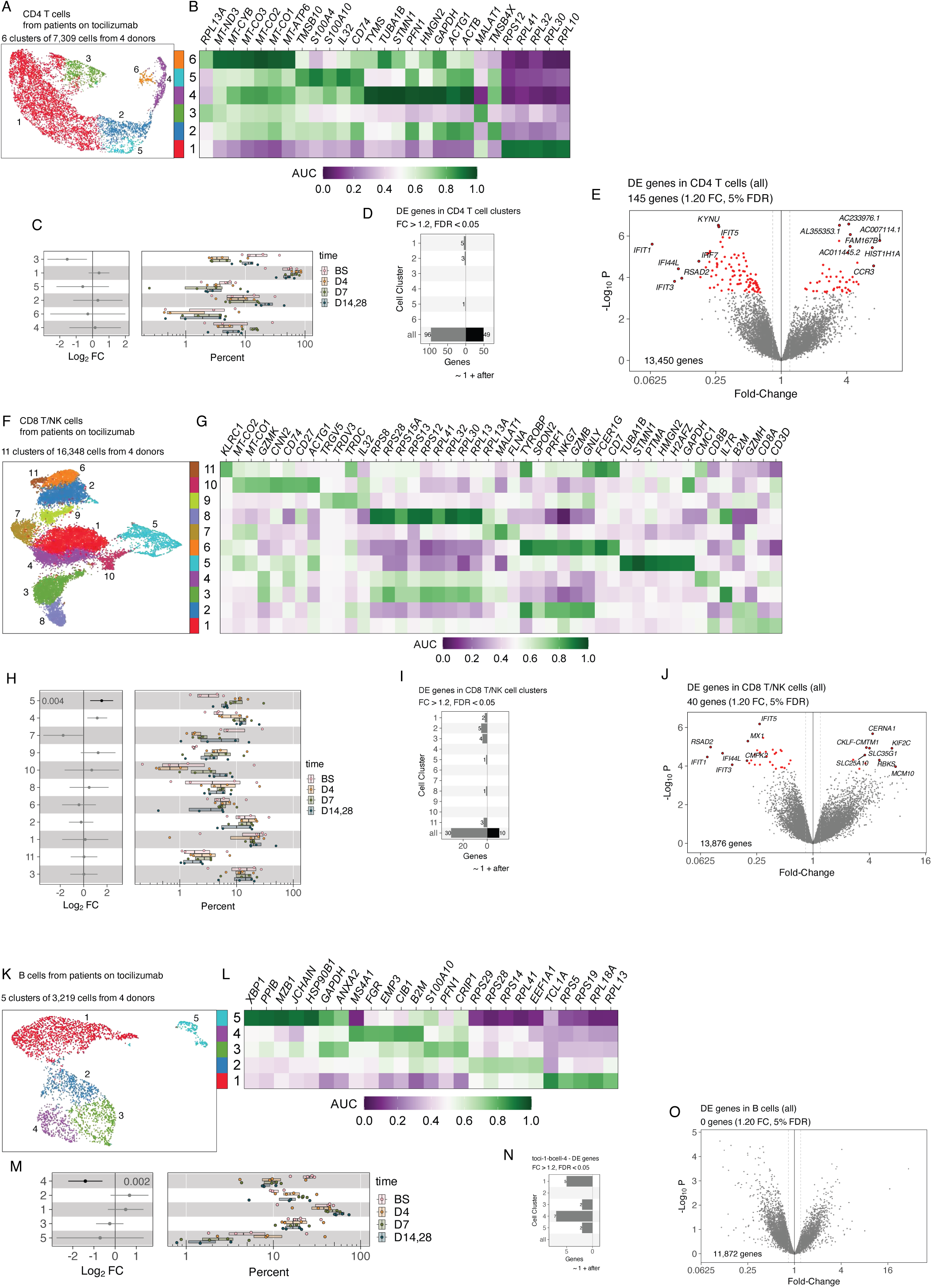
CD4 T cells, CD8 T/NK cells, and B cells from the tocilizumab data. **A, F, K.** UMAP embedding of CD4 T cells (**A**), CD8 T/NK cells (**F**), B cells (**K**), hue indicates membership to cell subsets. **B, G, L.** Heatmap of marker genes selected for each cell subset, hue indicates AUC. **C, H, M.** Box plots of cell subset abundances (right panel). Dots represent patients, box plots indicate median and interquartile range. Hue indicates time point after tocilizumab; x-axis indicates percent of cells assigned to each cell subset. Error bars (left panel) indicate the 95% CI of the estimated log2 fold-change for each cell subset over time. **D, I, N.** Grey and black bars indicate the number of differentially expressed genes in each cell subset with absolute fold-change > 1.2 and FDR < 5%. **E, J, O.** Volcano plot where each dot is a gene, x-axis indicates fold-change and y-axis indicates negative log10 p-value. Red indicates FDR < 5%.

**Figure S15.**
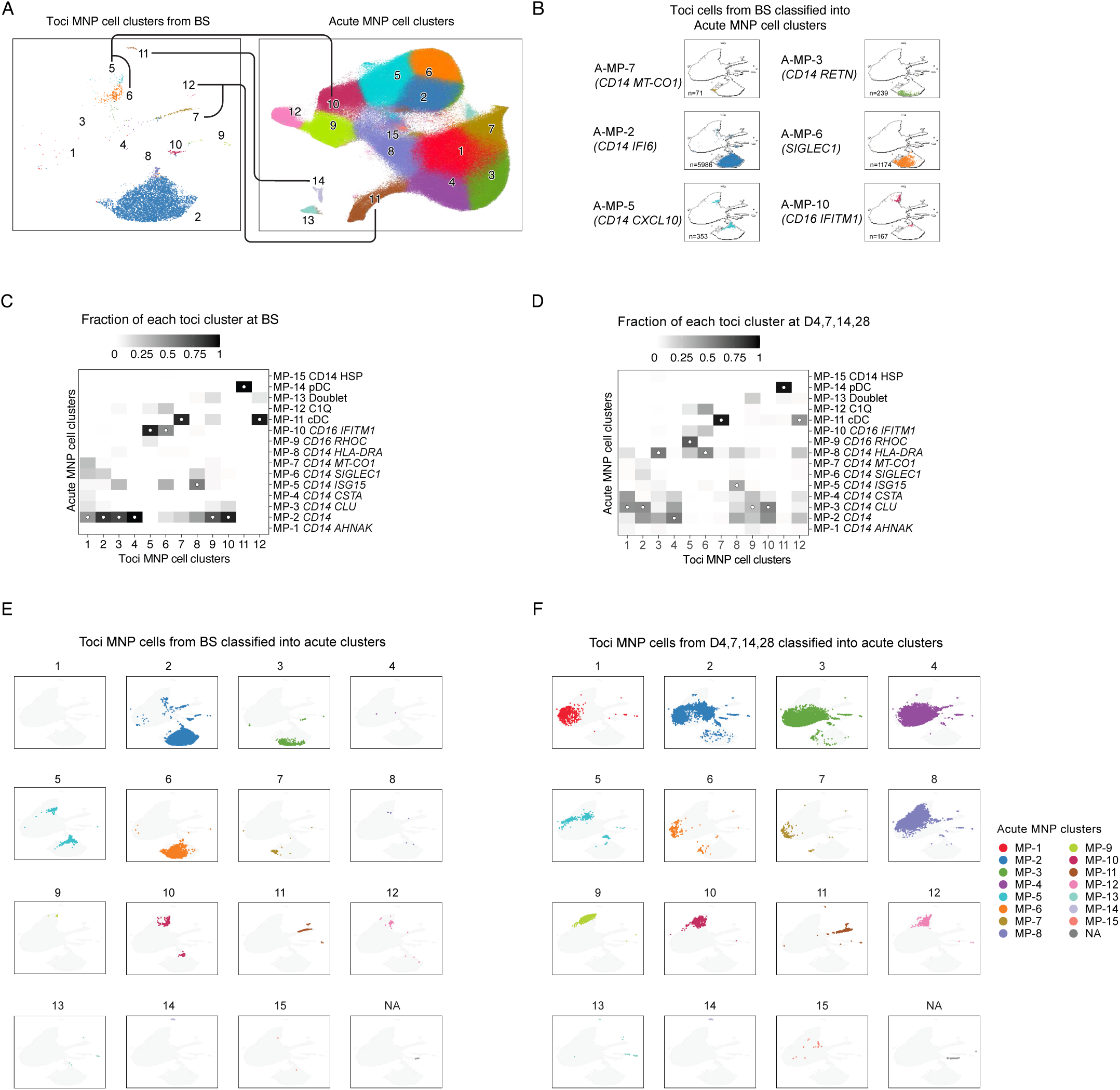
Classification of MNPs from the tocilizumab data into subsets from the acute data, continued. **A.** UMAP embeddings for Tocilizumab MNP cells at time point BS (left) and Acute MNP cells (right). Lines indicate the mapping between cell subsets (e.g. Toci 5,6 maps to Acute 10). **B.** Tocilizumab MNP cells from time point BS classified into the cell subsets from the acute dataset. Hue indicates the Acute cell subset, so the tocilizumab cells are colored by the acute labels. Text labels are the names of the acute data cell subsets. Number of Tocilizumab MNP cells from time point BS are shown in the lower-left corner. **C, D.** Heatmaps indicating the fraction of cells from each toci cell subset (columns) at baseline (**C**) or later time points (D4, 7, 14, 28) (**D**) that were classified into each acute data cell subset (rows). White dot in each column indicates the row with maximum value. **E, F.** UMAP embeddings for Tocilizumab MNP cells at baseline (**E**) or later time points (**F**). Each toci cell is classified into one of the acute subsets. Facets and colors indicate acute cell subsets.

**Figure S16.**
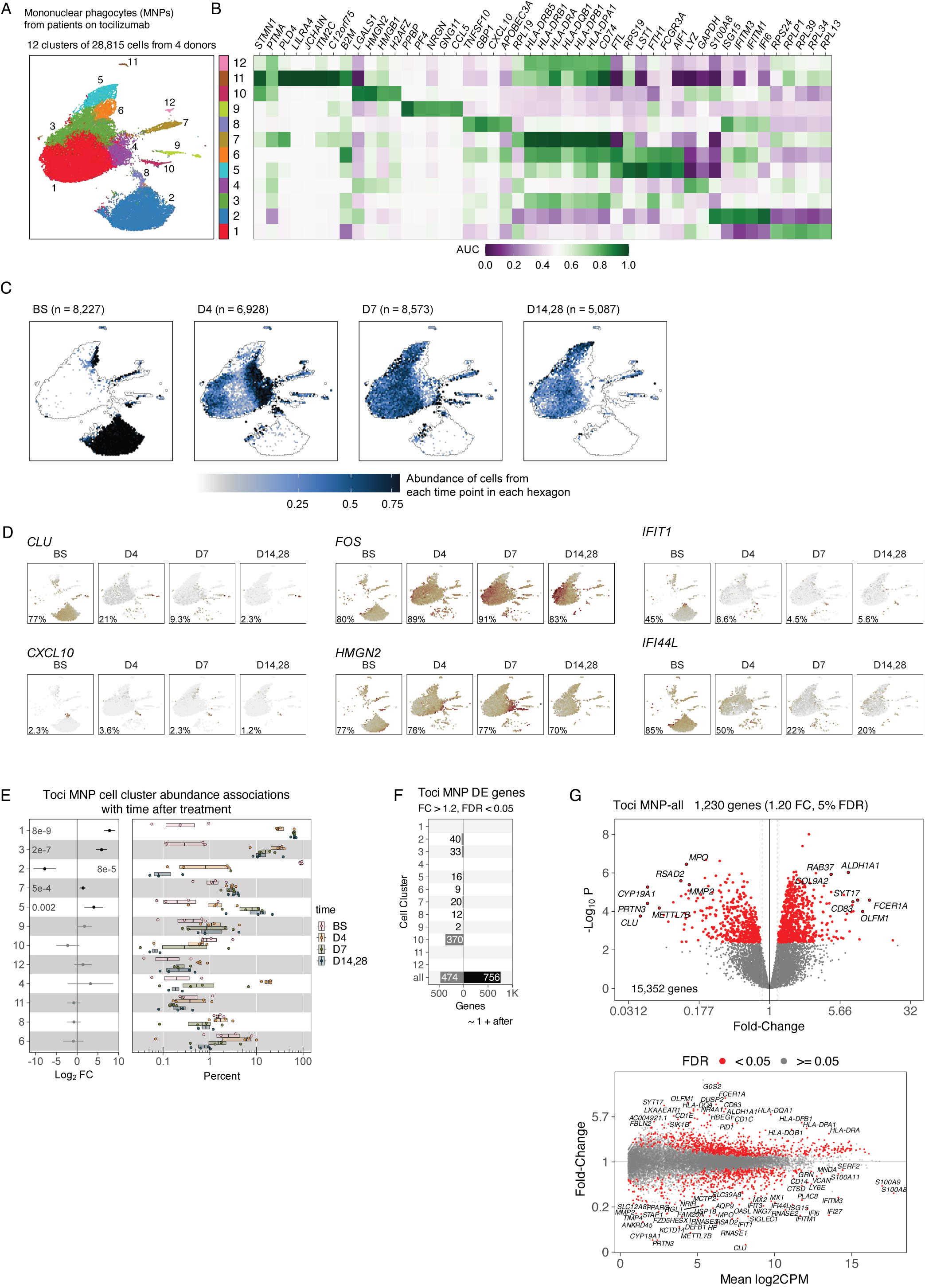
MNPs from the tocilizumab data. **A.** UMAP embedding of 28,815 MNPs, hue indicates membership to 12 subsets. **B.** Heatmap of marker genes selected for each cell subset, hue indicates AUC. **C.** UMAP embedding faceted by time point (BS; D4; D7; D14,28). Data are binned into hexagons, and the color of each hexagons indicates the proportion of data from all time points that is located in that part of the UMAP embedding. **D.** UMAP embeddings faceted by time point, showing the binned gene expression level of four genes (*CLU*, *FOS*, *CXCL10*, *HMGN2*). Hue indicates the mean log2 CPM level of the cells in each hexagon. **E.** Box plots of cell subset abundances (right panel). Dots represent patients, box plots indicate median and interquartile range. Hue indicates time point after tocilizumab; x-axis indicates percent of cells assigned to each cell subset. Error bars (left panel) indicate the 95% CI of the estimated log2 fold-change for each cell subset over time. **F.** Grey and black bars indicate the number of differentially expressed genes in each cell subset with absolute fold-change > 1.2 and FDR < 5%. **G.** Volcano plot (top) where each dot is a gene, x-axis indicates fold-change and y-axis indicates negative log10 p-value. MA plot (bottom) where each dot is gene, x-axis indicates mean log2 CPM gene expression and y-axis indicates fold-change. Red indicates FDR < 5%.

**Figure S17.**
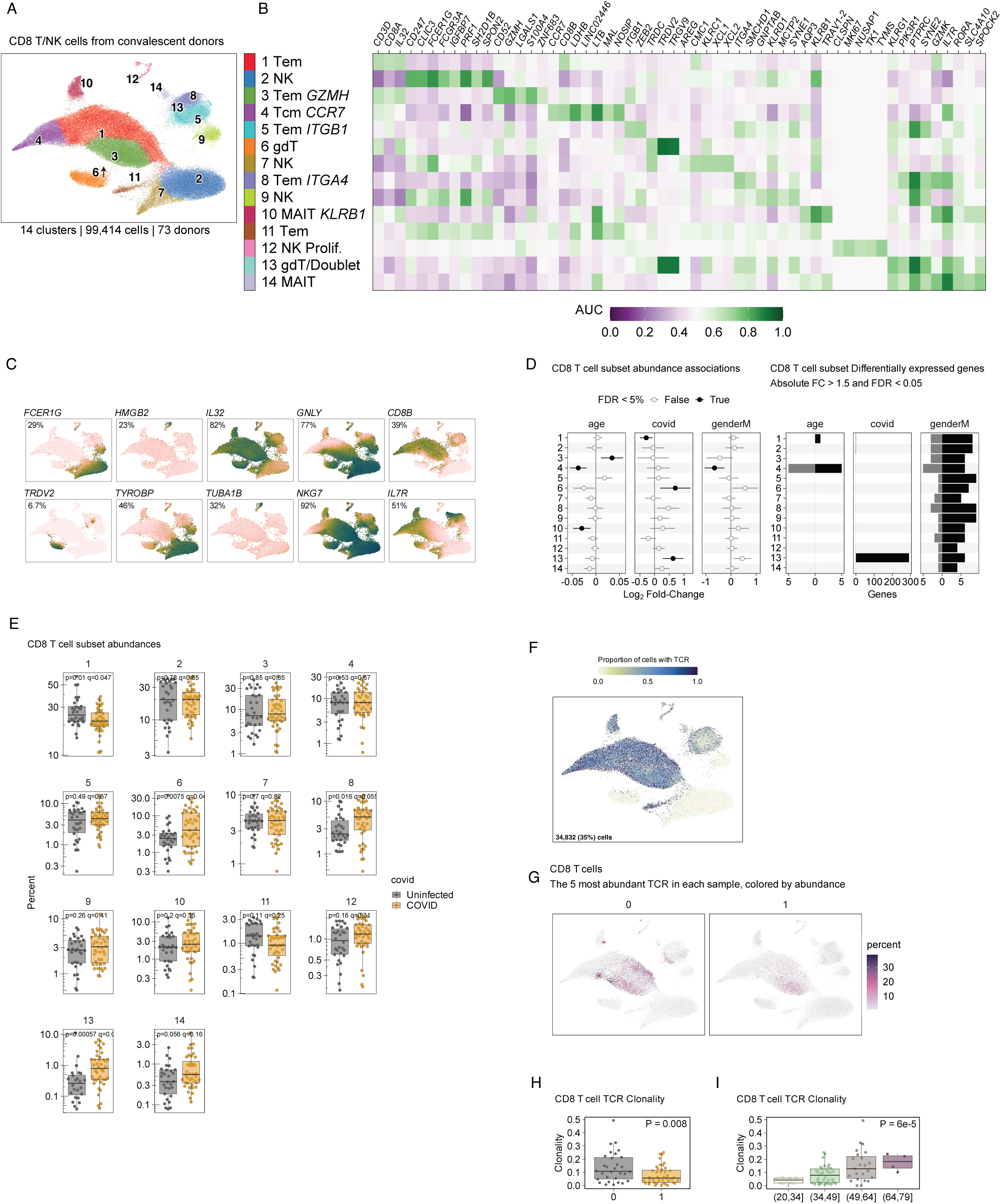
CD8 T cells from the convalescent data. **A.** UMAP embedding of 99,414 CD8 T/NK cells, hue indicates membership to 14 subsets. **B.** Heatmap of marker genes selected for each cell subset, hue indicates AUC. **C.** UMAP panels with gene expression levels for genes across all CD8 T/NK cells: *FCER1G*, *HMGB2*, *IL32*, *GNLY*, *CD8B*, *TRDV2*, *TYROBP*, *TUBA1B*, *NKG7*, *IL7R*. The percent of cells with non-zero expression is shown in each panel. **D.** Cell subset abundance (left) and gene expression (right) associations. Error bars indicate 95% CI for coefficients age, covid, and genderM from multivariate linear regression analysis. Black circles indicate FDR < 5%. Grey and black bars indicate the number of differentially expressed genes in each cell subset with absolute fold-change > 1.5 and FDR < 5%. **E.** Box plots of cell subset abundances. Dots represent patients, box plots indicate median and interquartile range. Hue indicates COVID infected or Uninfected, y-axis indicates percent of CD8 T cells assigned to each cell subset. **F.** UMAP of all CD8 T/NK cells, with hexagons representing cells in each region of the UMAP embedding. Hue indicates the percent of cells in a hexagon that have TCR data. The number and percent of cells with TCR data is shown. **G.** UMAP of all CD8 T/NK cells, faceted by COVID infection (0 Uninfected and 1 Infected). Hue indicates percent of cells with TCRs that are present in the list of top 5 most abundant TCRs from each sample. **H.** Box plot of CD8 T cell TCR clonality, by COVID infection. Dots represent patients, box plots indicate median and interquartile range. X-axis and hue indicates COVID infected or Uninfected, y-axis indicates clonality of TCRs in each sample. P-value indicates association with COVID infection. **I.** Box plot of CD8 T cell TCR clonality, by age. Dots represent patients, box plots indicate median and interquartile range. X-axis and hue indicates age of patients, y-axis indicates clonality of TCRs in each sample. P-value indicates association with age.

**Figure S18.**
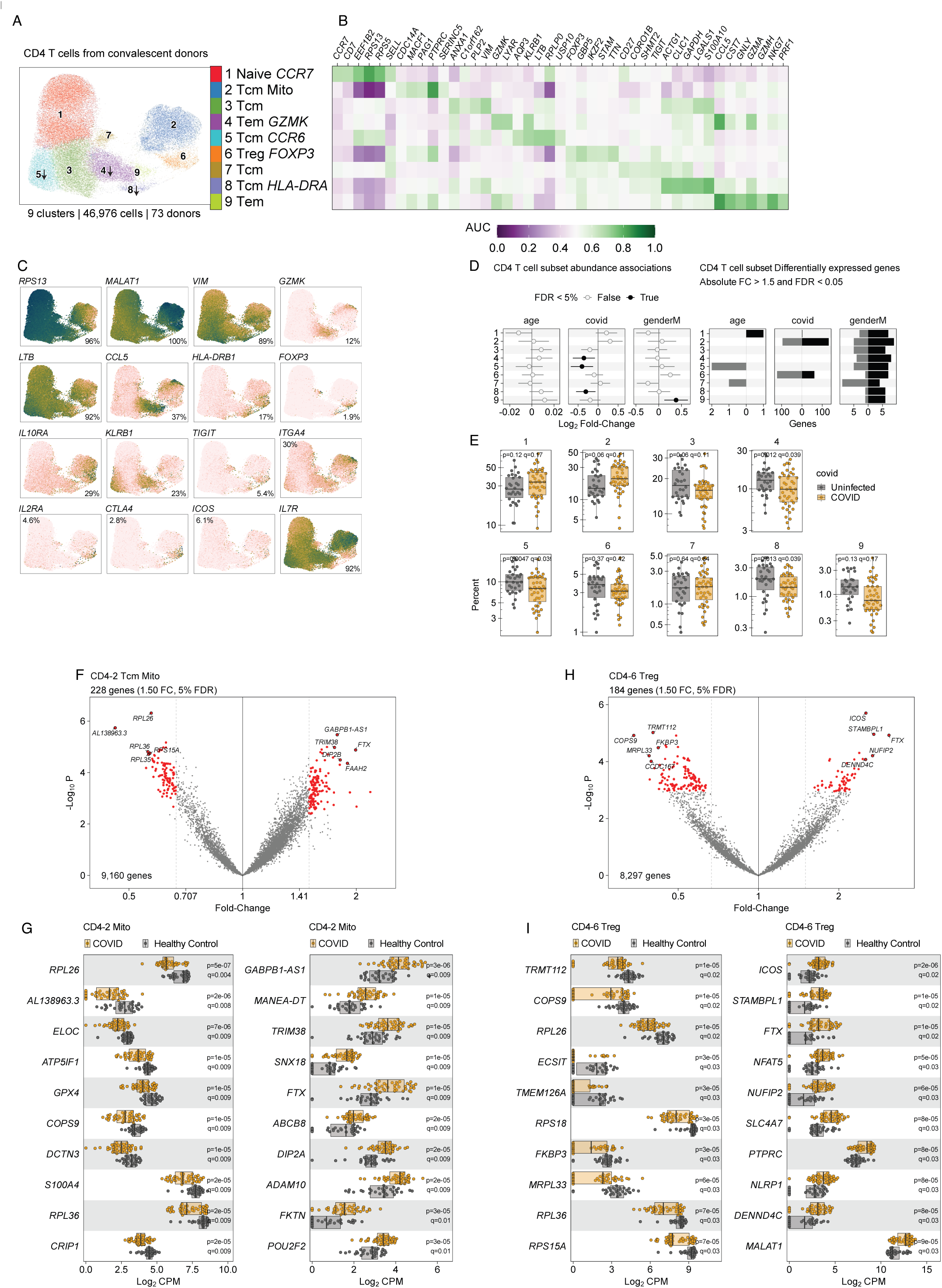
CD4 T cells from the convalescent data. **A.** UMAP embedding of 46,976 CD4 T cells, hue indicates membership to 9 subsets. **B.** Heatmap of marker genes selected for each cell subset, hue indicates AUC. **C.** UMAP panels with gene expression levels for genes across all CD4 T cells: *RPS13*, *MALAT1*, *VIM*, *GZMK*, *LTB*, *CCL5*, *HLA-DRB1*, *FOXP3*, *IL10RA*, *KLRB1*, *TIGIT*, *ITGA4*, *IL2RA*, *CTLA4*, *ICOS*, *IL7R*. The percent of cells with non-zero expression is shown in each panel. **D.** Cell subset abundance (left) and gene expression (right) associations. Error bars indicate 95% CI for coefficients age, covid, and genderM from multivariate linear regression analysis. Black circles indicate FDR < 5%. Grey and black bars indicate the number of differentially expressed genes in each cell subset with absolute fold-change > 1.5 and FDR < 5%. **E.** Box plots of cell subset abundances. Dots represent patients, box plots indicate median and interquartile range. Hue indicates COVID infected or Uninfected, y-axis indicates percent of CD4 T cells assigned to each cell subset. **F.** Volcano plot for genes in cell subset CD4-2 (Tcm Mito) associated with COVID infection. X-axis represents fold-change for each gene, and y-axis represents negative log10 p-value. **G.** Box plots for gene expression in cell subset CD4-6 (Treg) associated with COVID infection. P-values and q-values (FDR) are shown for each gene. Left panel shows repressed genes and right panel shows induced genes. **H.** Volcano plot for genes in cell subset CD4-6 (Treg) associated with COVID infection. X-axis represents fold-change for each gene, and y-axis represents negative log10 p-value. **I.** Box plots for gene expression in cell subset CD4-6 (Treg) associated with COVID infection. Dots represent patients, box plots indicate median and interquartile range. P-values and q-values (FDR) are shown for each gene. Left panel shows repressed genes and right panel shows induced genes.

**Figure S19.**
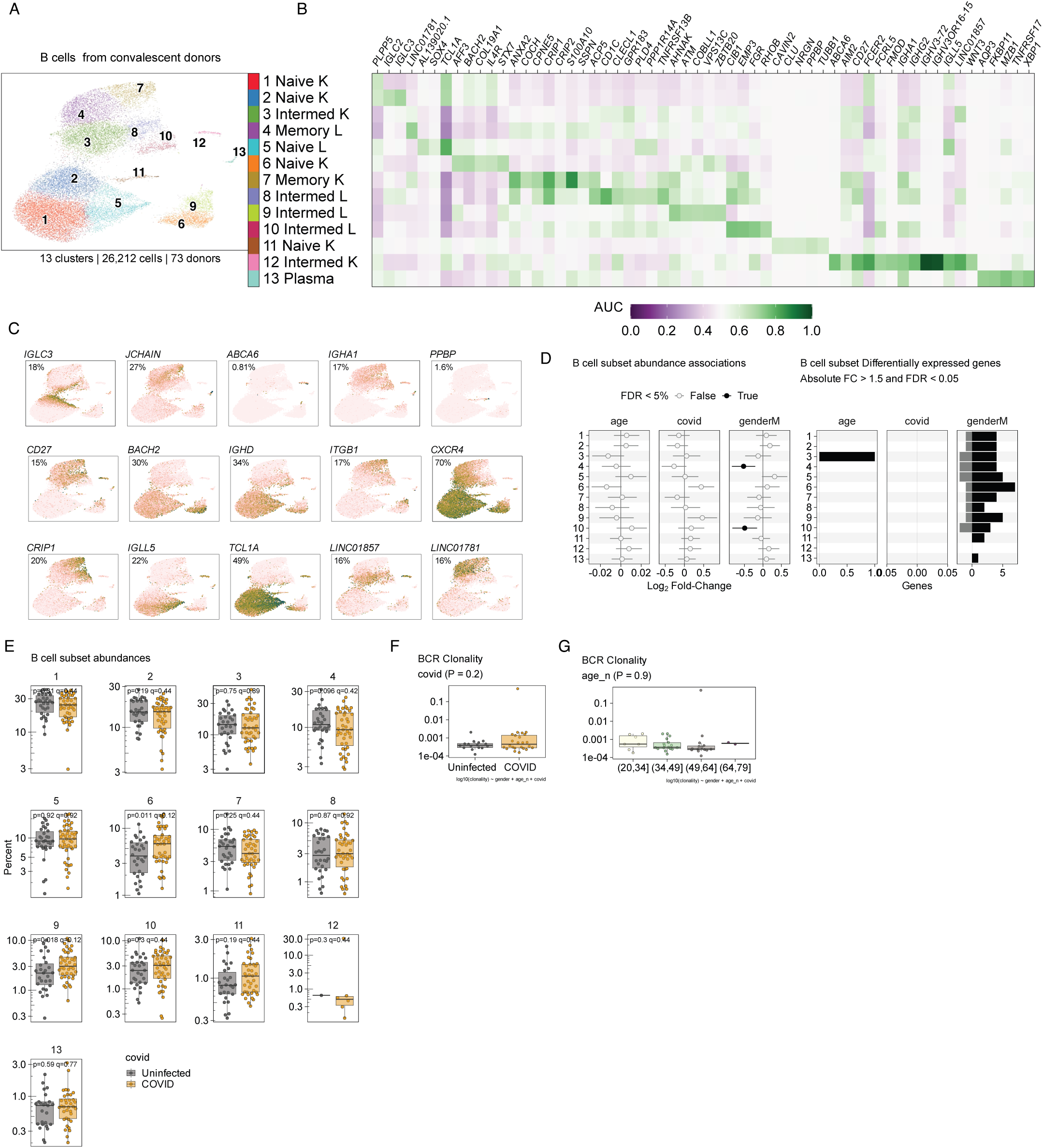
B cells from the convalescent data. **A.** UMAP embedding of 26,212 B cells, hue indicates membership to 13 subsets. **B.** Heatmap of marker genes selected for each cell subset, hue indicates AUC. **C.** UMAP panels with gene expression levels for genes across all MNPs: *IGLC3*, *JCHAIN*, *ABCA6*, *IGHA1*, *PPBP*, *CD27*, *BACH2*, *IGHD*, *ITGB1*, *CXCR4*, *CRIP1*, *IGLL5*, *TCL1A*, *LINC01857*, *LINC01781*. The percent of cells with non-zero expression is shown in each panel. **D.** Cell subset abundance (left) and gene expression (right) associations. Error bars indicate 95% CI for coefficients age, covid, and genderM from multivariate linear regression analysis. Black circles indicate FDR < 5%. Grey and black bars indicate the number of differentially expressed genes in each cell subset with absolute fold-change > 1.5 and FDR < 5%. **E.** Box plots of cell subset abundances. Dots represent patients, box plots indicate median and interquartile range. Hue indicates COVID infected or Uninfected, y-axis indicates percent of CD8 T cells assigned to each cell subset. **F.** Box plot of B cell BCR clonality, by COVID infection. Dots represent patients, box plots indicate median and interquartile range. X-axis and hue indicates COVID infected or Uninfected, y-axis indicates clonality of TCRs in each sample. P-value indicates association with COVID infection. **G.** Box plot of B cell BCR clonality, by age. Dots represent patients, box plots indicate median and interquartile range. X-axis and hue indicates age of patients, y-axis indicates clonality of TCRs in each sample. P-value indicates association with age.

**Figure S20.**
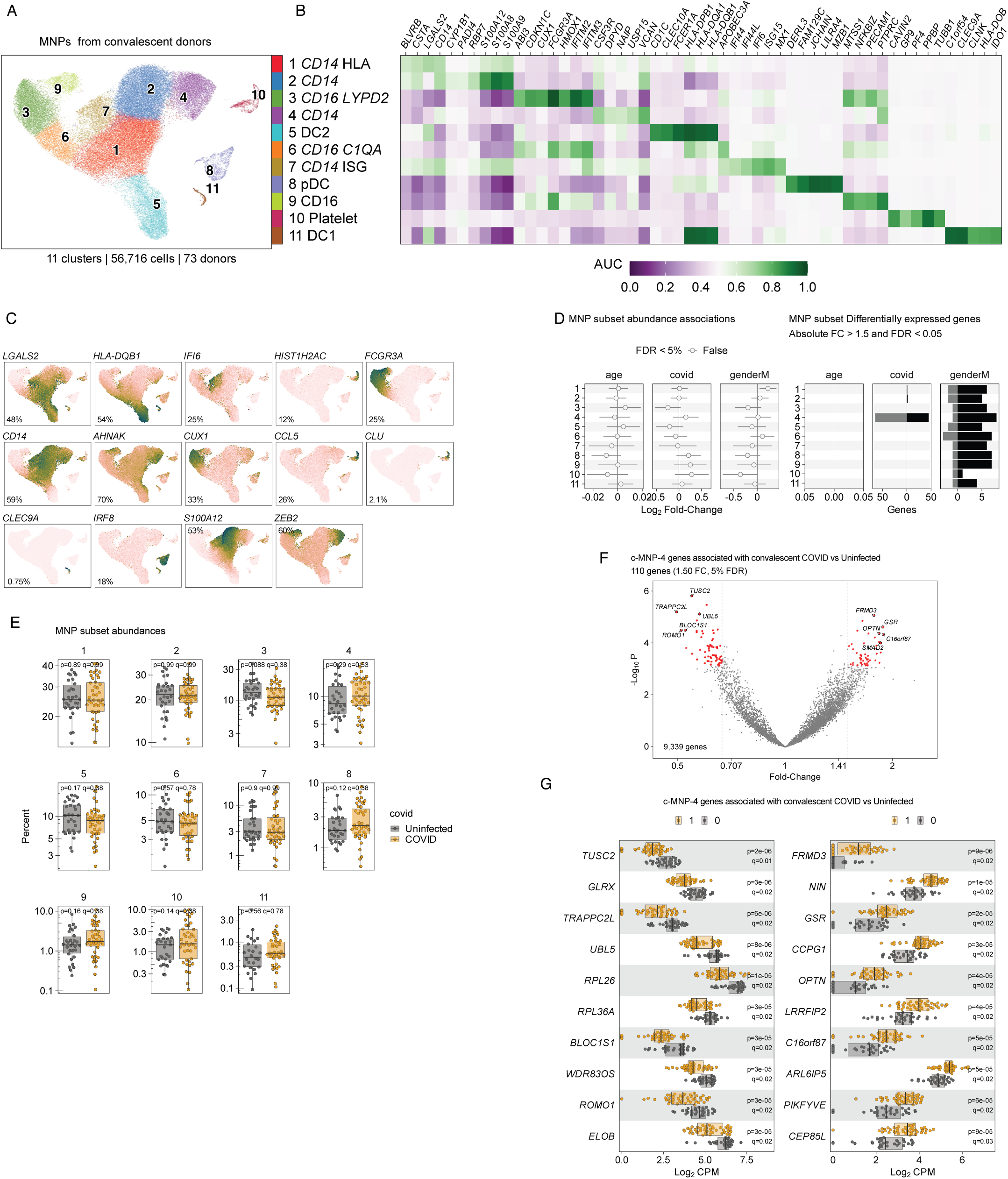
Mononuclear phagocytes from the convalescent data. **A.** UMAP embedding of 56,617 mononuclear phagocytes (MNPs), hue indicates membership to 11 subsets. **B.** Heatmap of marker genes selected for each cell subset, hue indicates AUC. **C.** UMAP panels with gene expression levels for genes across all MNPs: *LGALS2*, *HLA-DQB1*, *IFI6*, *HIST1H2AC*, *FCGR3A*, *CD14*, *AHNAK*, *CUX1*, *CCL5*, *CLU*, *CLEC9A*, *IRF8*, *S100A12*, *ZEB2*. The percent of cells with non-zero expression is shown in each panel. **D.** Cell subset abundance (left) and gene expression (right) associations. Error bars indicate 95% CI for coefficients age, covid, and genderM from multivariate linear regression analysis. Black circles indicate FDR < 5%. Grey and black bars indicate the number of differentially expressed genes in each cell subset with absolute fold-change > 1.5 and FDR < 5%. **E.** Box plots of cell subset abundances. Dots represent patients, box plots indicate median and interquartile range. Hue indicates COVID infected or Uninfected, y-axis indicates percent of CD8 T cells assigned to each cell subset. **F.** Volcano plot for genes in cell subset MNP-4 (CD14) associated with convalescent COVID infection. X-axis represents fold-change for each gene, and y-axis represents negative log10 p-value. **G.** Box plots for gene expression in cell subset MNP-4 (CD14) associated with convalescent COVID infection. Dots represent patients, box plots indicate median and interquartile range. P-values and q-values (FDR) are shown for each gene. Left panel shows repressed genes and right panel shows induced genes.

**Figure S21.**
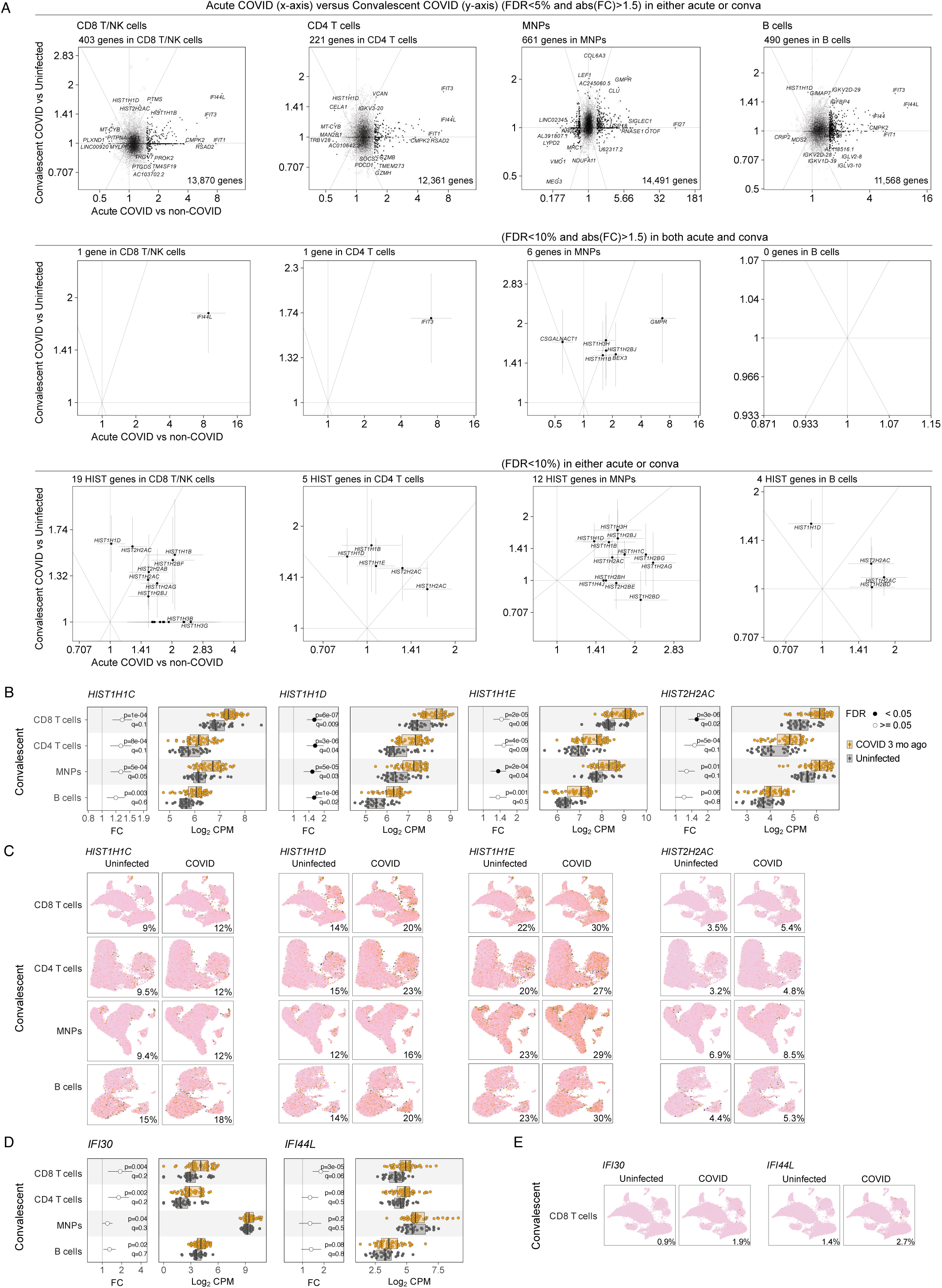
Comparison of differential gene expression results from the convalescent data and the acute data. **A.** Scatter plot where each point represents a gene, the x-axis represents the fold-change associated with COVID infection in the acute data, and the y-axis represents the fold-change associated with COVID infection in the convalescent data. From left to right, the columns represent cell types: CD8 T/NK cells, CD4 T cells, MNPs, and B cells. The top row of plots includes genes that meet FDR < 5% and abs(FC) > 1.5 in either the acute dataset or in the convalescent dataset. The middle row includes genes that meet FDR < 10% and abs(FC) > 1.5 in both datasets. The bottom row includes genes whose name starts with HIST and that meet FDR < 10% in either the acute dataset or in the convalescent dataset. Error bars indicate 95% confident intervals. **B.** Error bars and box plots for 4 genes (*HIST1H1C*, *HIST1H1D*, *HIST1H1E*, *HIST2H2AC*) in 4 cell lineages (CD8 T cells, CD4 T cells, MNPs, B cells). Dots and error bars indicate fold-change for COVID infected versus uninfected individuals in the convalescent dataset. Box plots indicate the log2CPM gene expression values, hue indicates COVID infection status. **C.** UMAP figures of gene expression, faceted by COVID infection status. Columns correspond to genes (*HIST1H1C*, *HIST1H1D*, *HIST1H1E*, *HIST2H2AC*) and rows to cell lineages (CD8 T cells, CD4 T cells, MNPs, B cells). Numbers in the lower-right corner indicate the percent of cells with non-zero expression of that gene, i.e., 9% of CD8 T cells in Uninfected individuals have expression of *HIST1H1C* and 12% of CD8 T cells in COVID infected individuals have expression. **D.** Same as B, error bars and box plots for 2 genes (*IFI30* and *IFI44L*). **E.** UMAP figures of gene expression, faceted by COVID infection status. Columns correspond to genes (*IFI30* and *IFI44L*).

**Figure S22.**
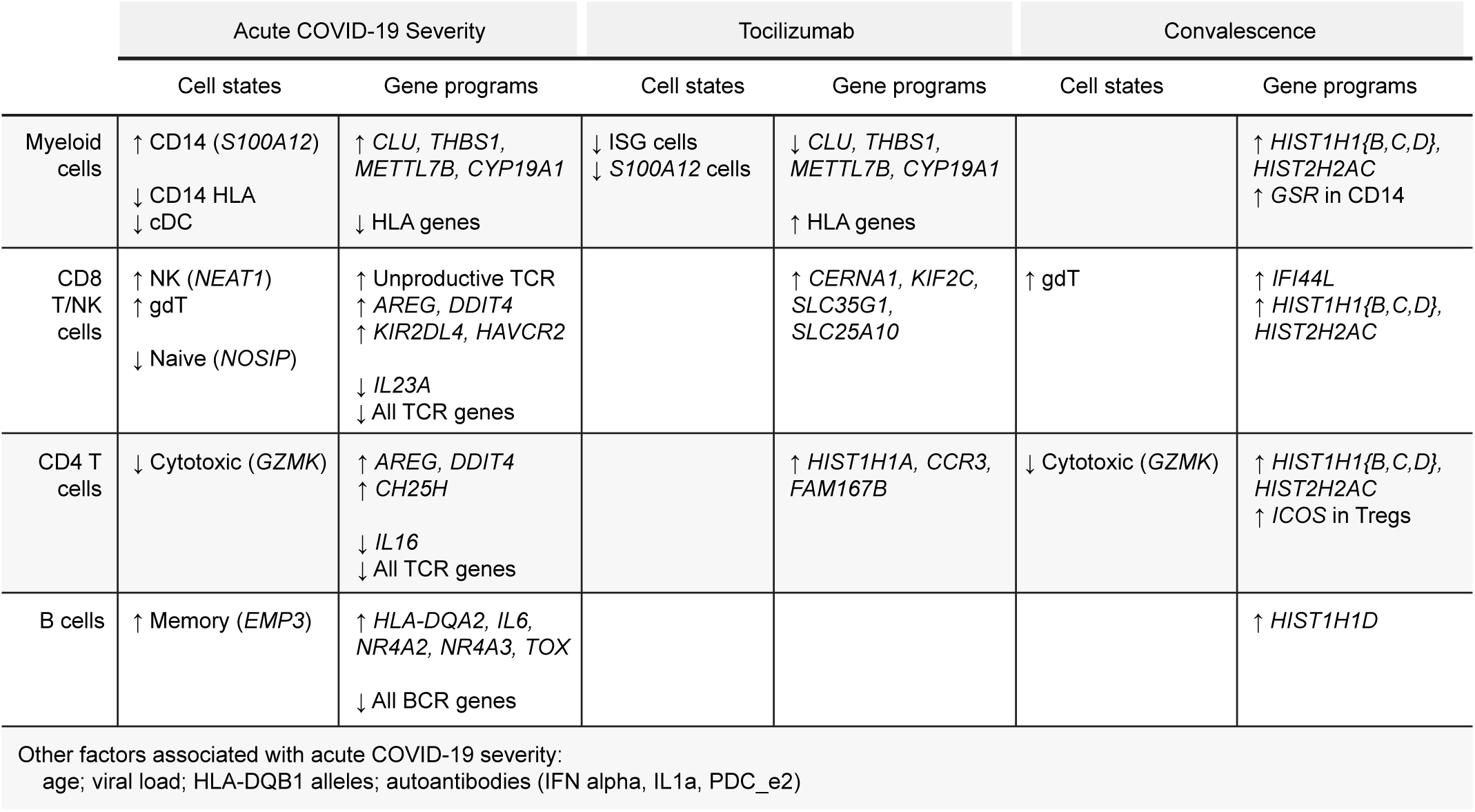
Summary table of main results.

## Supplementary Tables

Table with an overview of main results from this study.

**Table S1.**
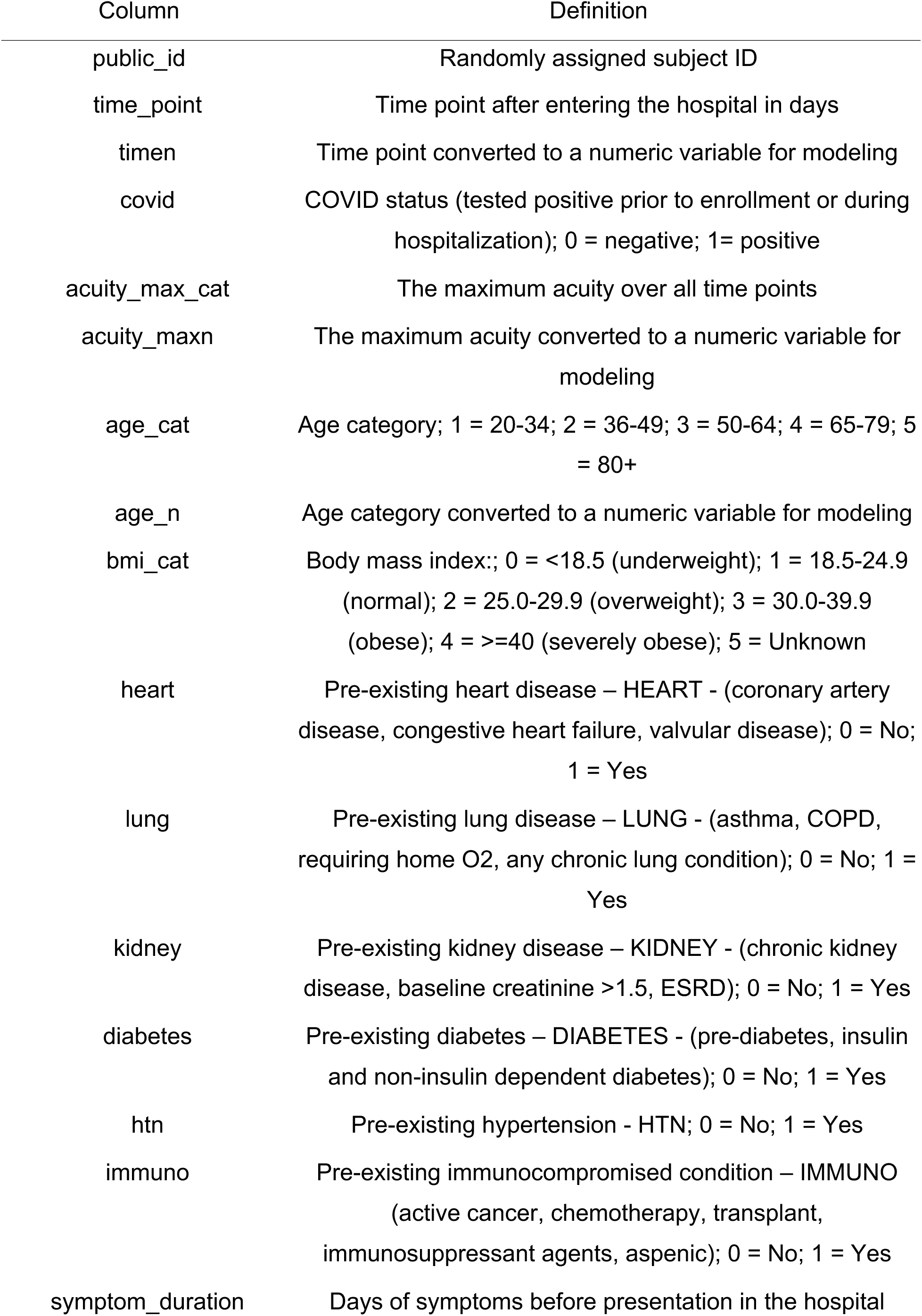
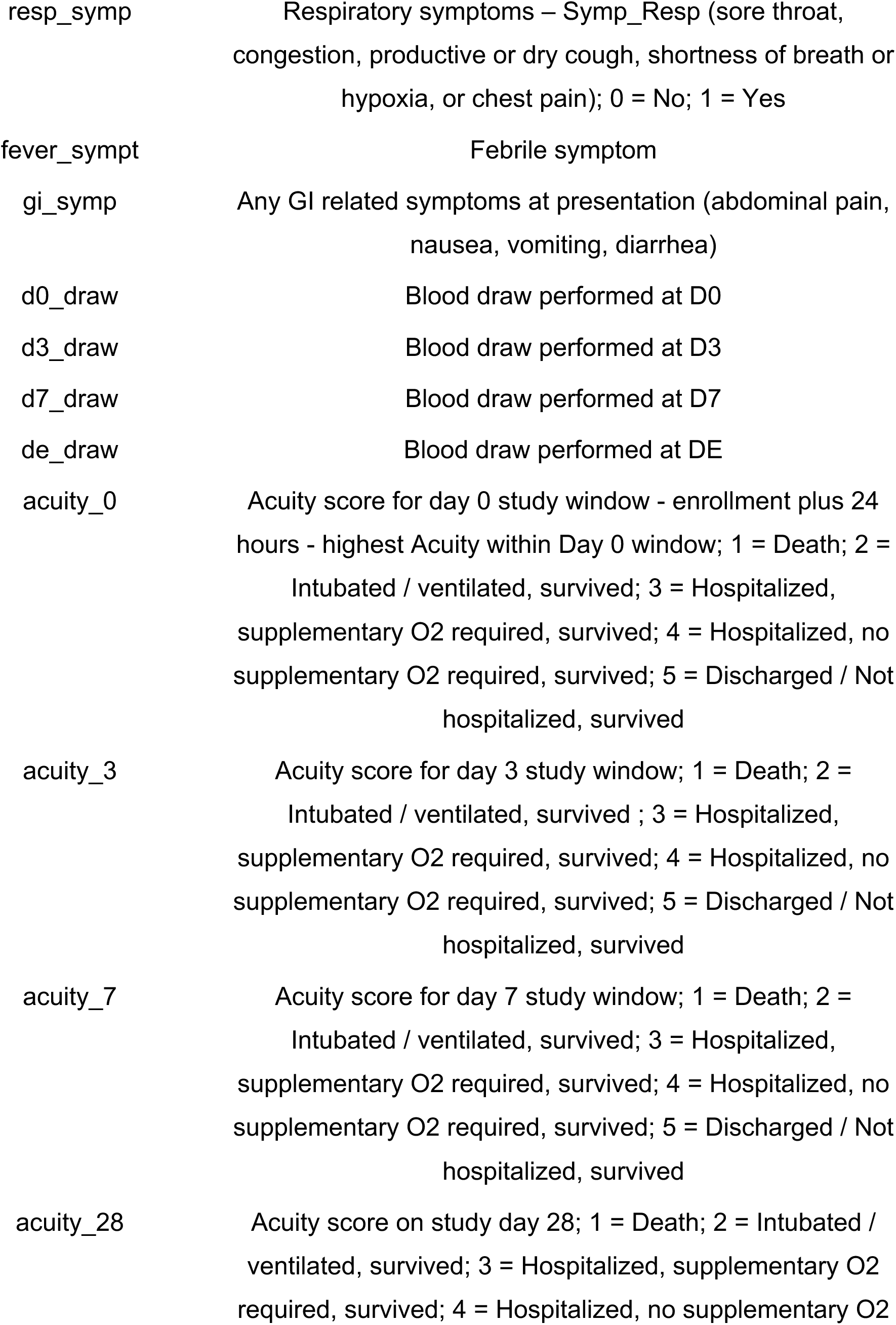
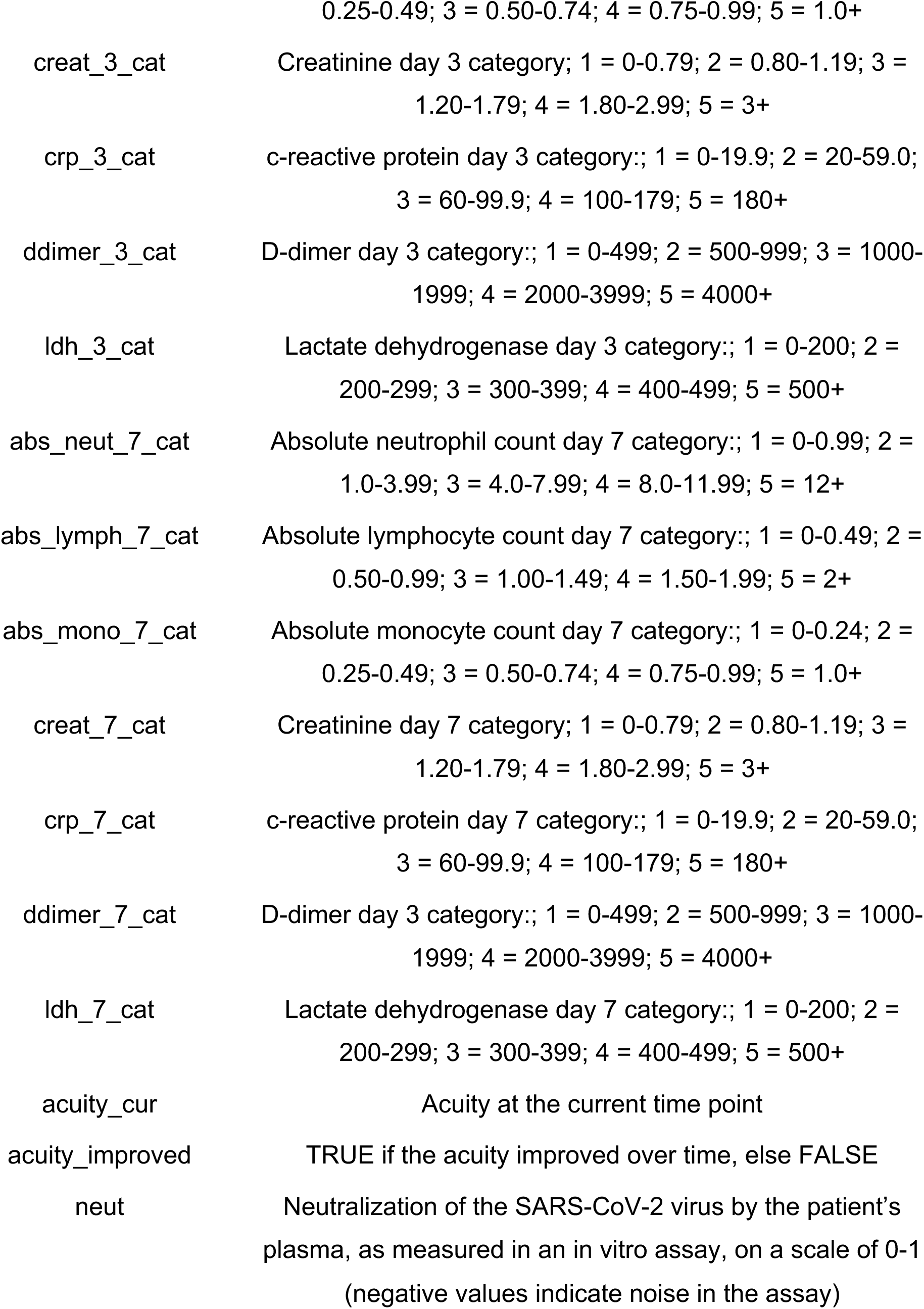
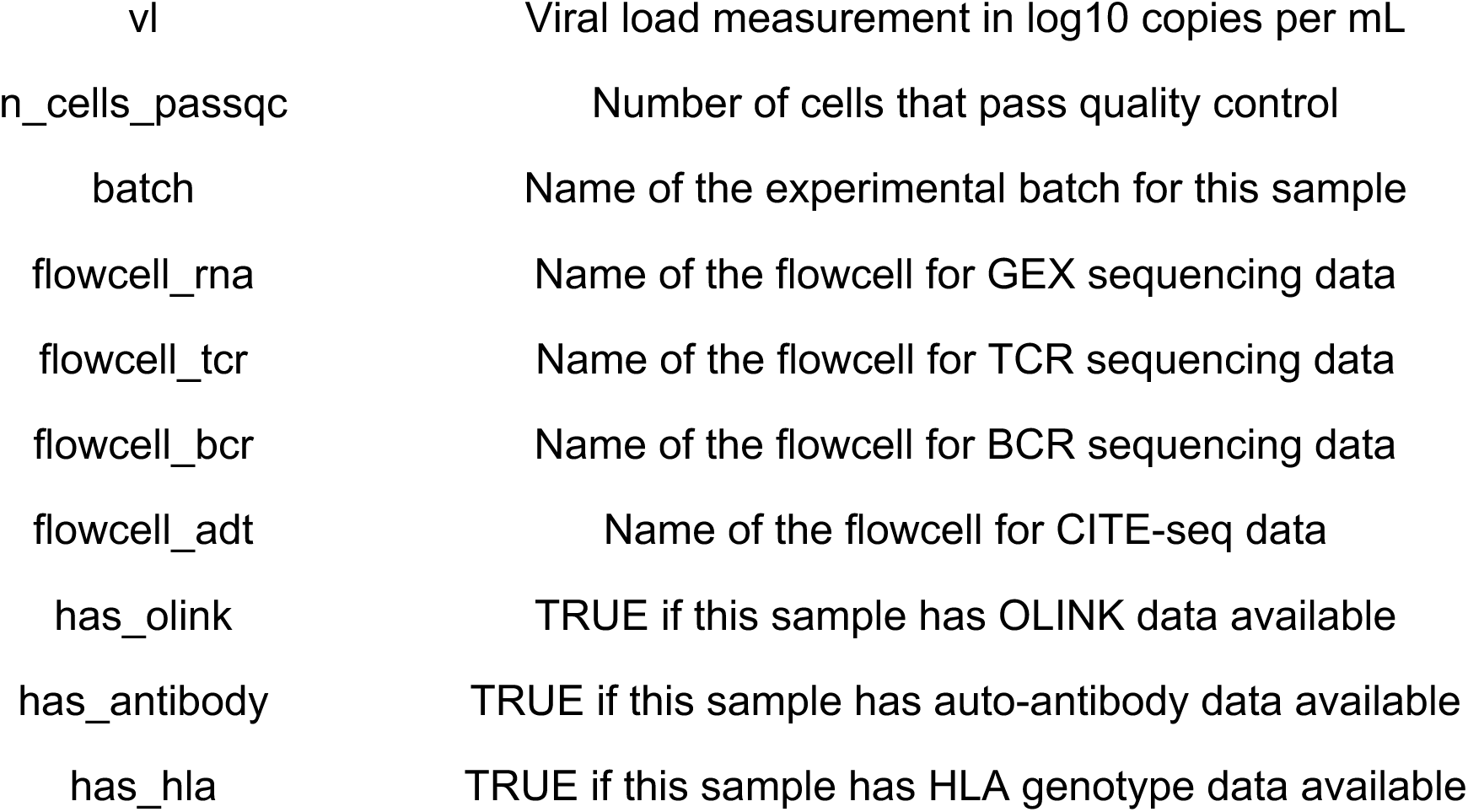
Patient metadata for the acute data. Information about each patient sample from the acute data. The table includes metadata for 908 samples representing 355 patients at a total of 9 time points and 5 acuity levels.

**Table S2.**
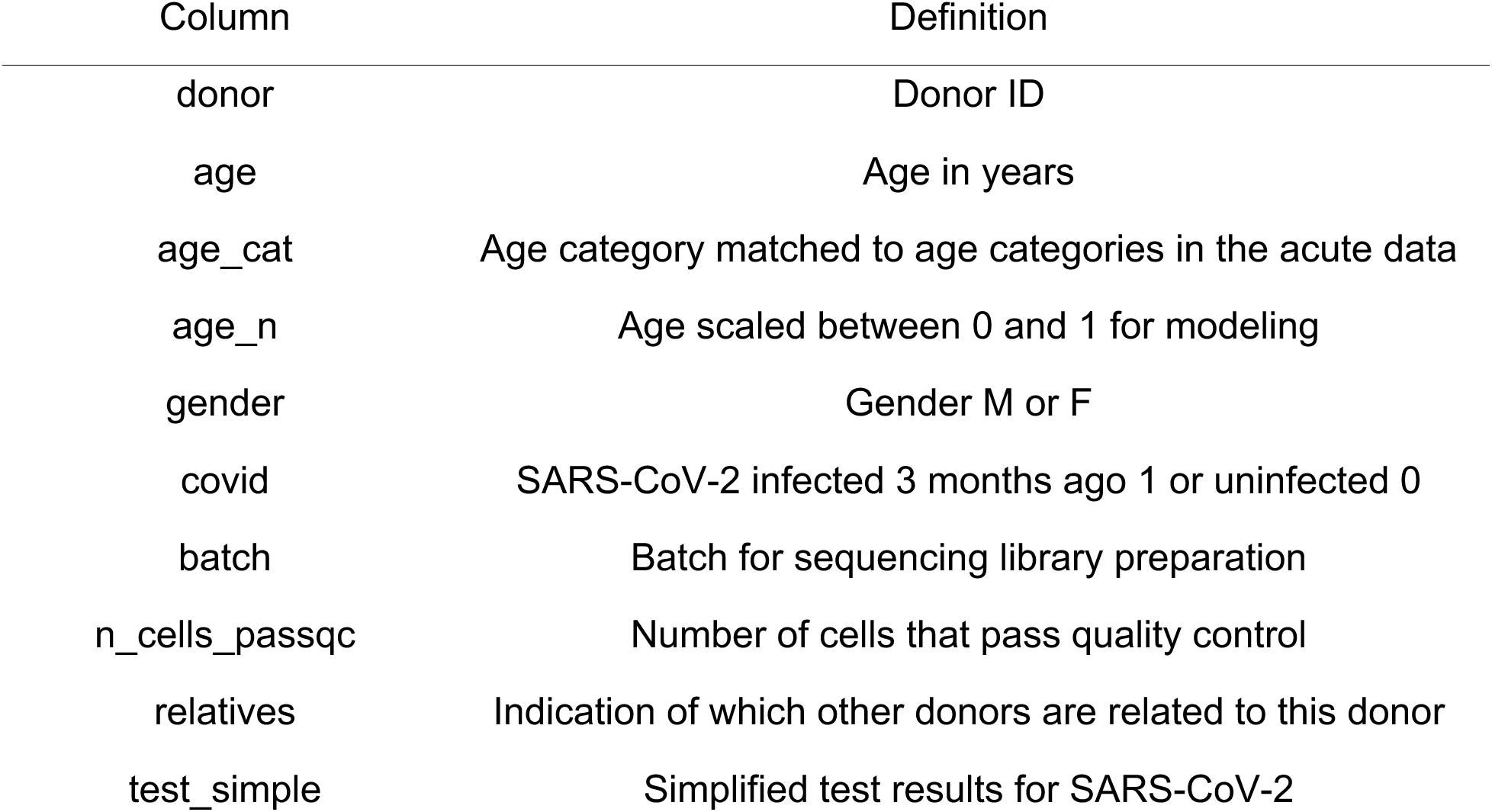
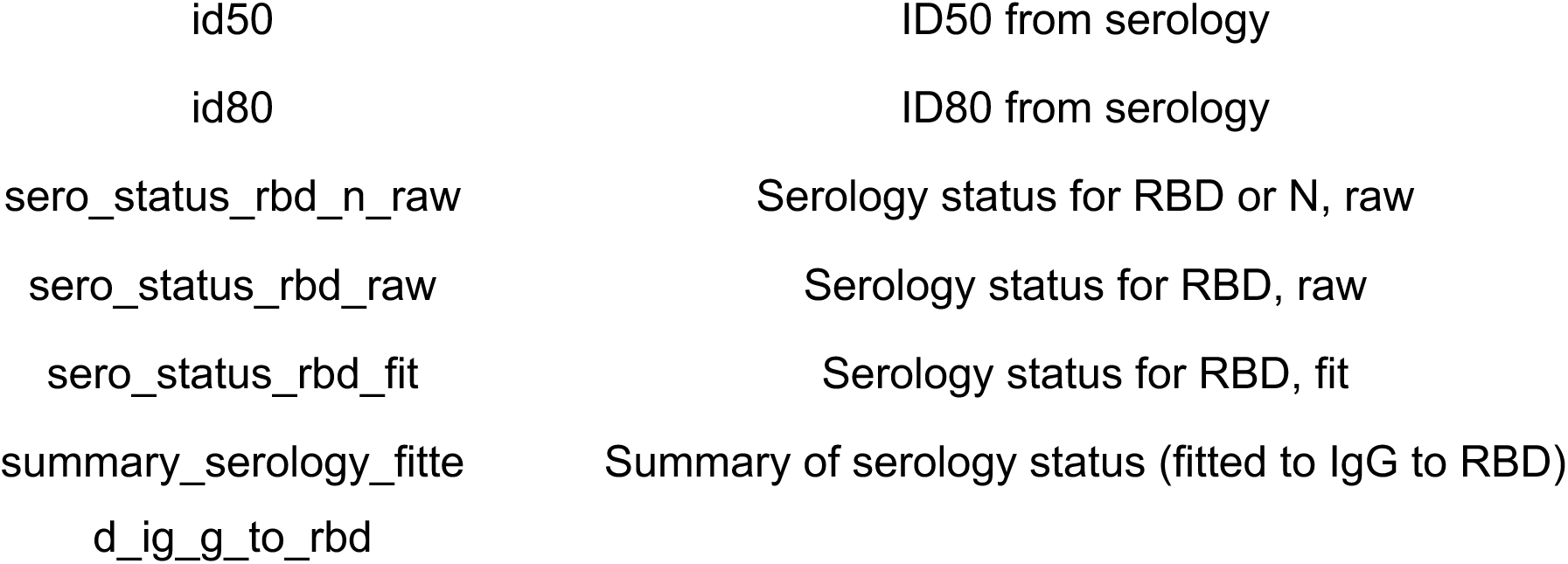
Patient metadata for the convalescent data. Information about each patient sample from the convalescent data. The table includes metadata for 152 patients infected with SARS-CoV-2 (n=91) or uninfected (n=61).

**Table S3.** Patient metadata for the tocilizumab data. Information about each patient sample from the tocilizumab data. The table includes metadata for 16 samples representing 4 patients treated with tocilizumab at a total of 5 time points.

| Column | Definition |
| --- | --- |
| donor | Donor ID |
| age | Donor age |
| gender | Donor gender |
| time | Days after tocilizumab BS=0 |
| n_cells_passqc | Number of cells in scRNA-seq that pass quality control |

**Table S4.**
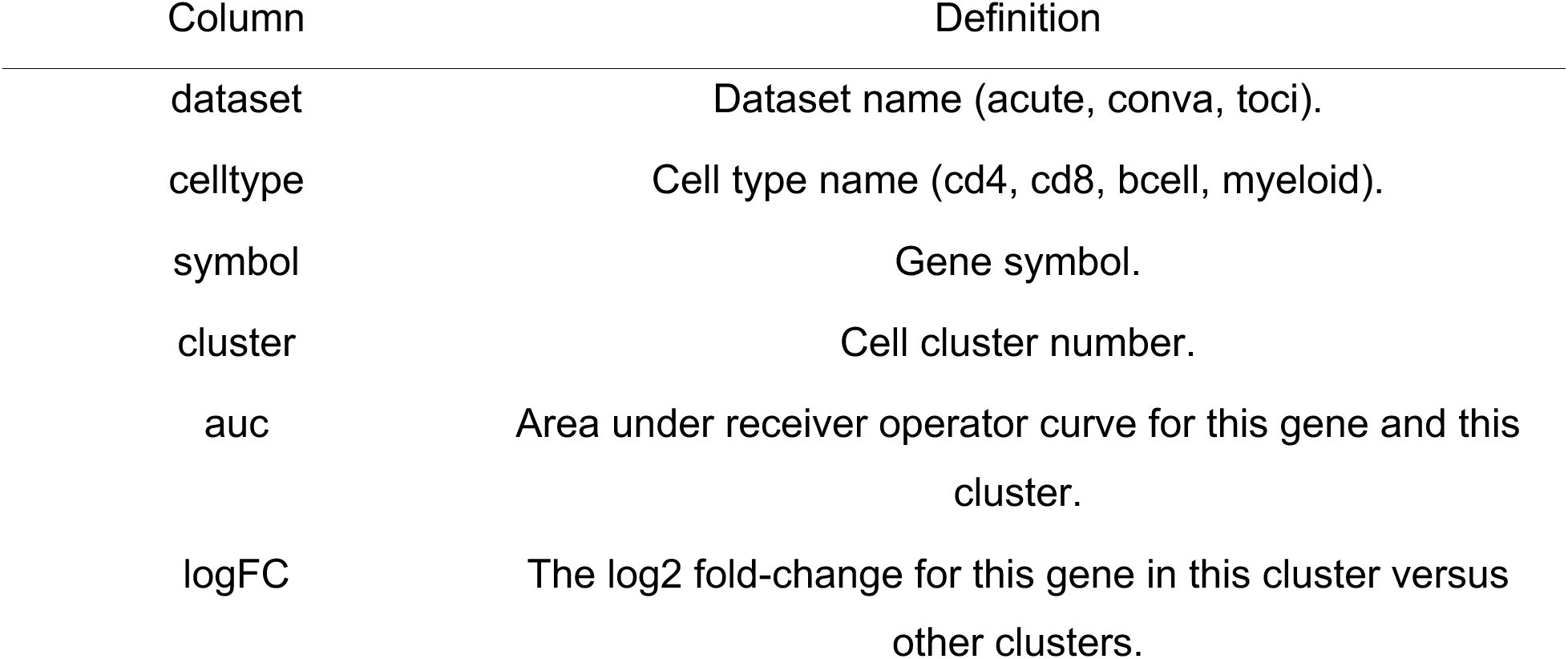
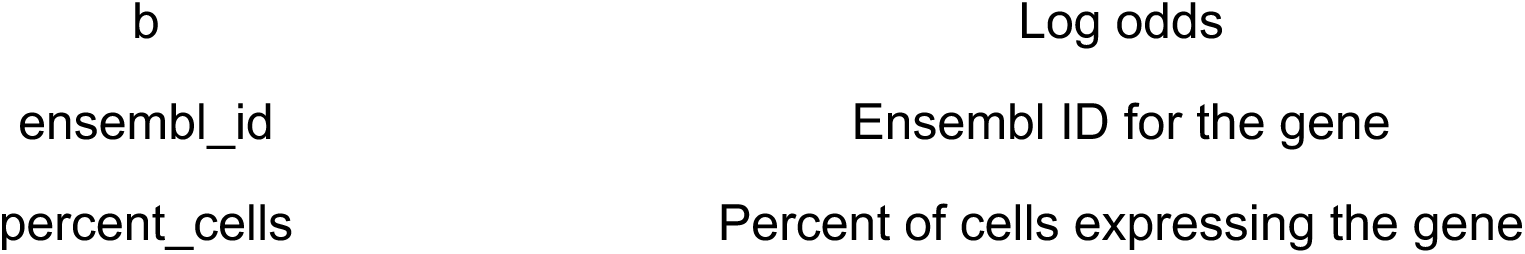
Cell cluster marker one-versus-all statistics. For each cell cluster in the acute data, tocilizumab data, and convalescent data, one row per gene that lists the statistics for that gene. The statistics describe how well each gene can discriminate cells that belong to some cluster versus cells that do not belong to that cluster. The table has 3,200,798 rows, one row for each gene (n=29,601) in each cell cluster (n=135) from all three of the datasets (acute, convalescent, and tocilizumab).

**Table S5.** Cell cluster abundance association. Cell cluster abundance association statistics for 135 cell clusters from all three of the datasets (acute, convalescent, and tocilizumab). See Methods for a description of the linear models that were fit for each dataset.

| Column | Definition |
| --- | --- |
| dataset | Dataset name (acute, conva, toci). |
| celltype | Cell type name (cd4, cd8, bcell, myeloid). |
| cluster | Cell cluster number. |
| coef | Name of the coefficient from the linear model. |
| logFC | The estimate of the coefficient for this cell cluster, interpretable as a log2 fold-change in the abundance of this cluster that is proportional to an increment of 1 in this variable. |
| ci_low | The lower bound of the 95% confidence interval for the estimate. |
| ci_high | The upper bound of the 95% confidence interval for the estimate. |
| t | The t-statistic for this cluster and this variable. |
| p | The p-value for this cluster and this variable. |
| fdr | The adjusted p-value (Benjamini-Hochberg) this cluster and this variable. |

**Table S6.** Differential gene expression statistics for the acute data (acuity, covid, neut, aa, vl). A table with gene expression statistics for four linear models fit to each gene in the acute data. We provide results for cell lineages (cd4, cd8, bcell, myeloid) as well as cell clusters within each lineage.

| Column | Definition |
| --- | --- |
| celltype | Name of the acute dataset cell lineage (cd4, cd8, bcell, myeloid) |
| cluster | Number of the cell cluster in the cell lineage |
| coef | Name of the parameter from the linear model |
| gene | Name of the gene |
| ave_expr | Average log2CPM expression of the gene |
| quantile_expr | Quantile of the average expression |
| p_value | P-value for the coefficient |
| adj_p_val | False Discovery Rate (adjusted p-value) |
| log_fc | Coefficient for the parameter (log2 Fold-Change) |
| ci_l | Lower bound of the 95% confidence interval for the log2 FC |
| ci_r | Upper bound of the 95% confidence interval for the log2 FC |
| t | t-statistic for the coefficient |
| b | Log odds |
| percent_cells | Percent of cells expressing the gene |
| rank | Rank of the gene, based on p-value and log2FC |
| model | Name of the model fit to the gene |

**Table S7.** Public BCR sequences observed in the acute data. BCR sequences that are observed in 2 or more acute donors.

| Column | Definition |
| --- | --- |
| public_id | Donor ID |
| time_point | Time point after entering the hospital in days |
| acuity_max_cat | The maximum acuity over all time points |
| age_cat | Age category |
| covid | COVID status (tested positive prior to enrollment or during hospitalization) |
| n_donors | Number of donors that have this BCR |
| n | Number of cells that have this BCR, per sample (donor_time) |
| rank | Rank of the abundance of this BCR, per sample (donor_time) |
| percent | This BCR accounts for what percent of BCRs from this sample |
| bcr_id | Concatenation of alpha and beta V, J, and CDR3 |
| IGHV | IGH V gene |
| IGHJ | IGH J gene |
| IGKV | IGK V gene |
| IGKJ | IGK J gene |
| IGLV | IGL V gene |
| IGLJ | IGL J gene |
| IGH_cdr3 | IGH CDR3 amino acid sequence |
| IGK_cdr3 | IGK CDR3 amino acid sequence |
| IGL_cdr3 | IGL CDR3 amino acid sequence |

**Table S8.** BCR association results for the acute data. Association results for each BCR with acuity in B cells.

| Column | Definition |
| --- | --- |
| celltype | Cell lineage (B cell) |
| test | Which gene (or genes) are tested in this row |
| IGHV | IGH V gene, if tested in this row or else NA |
| IGHJ | IGH J gene, if tested in this row or else NA |
| IGKV | IGK V gene, if tested in this row or else NA |
| IGKJ | IGK J gene, if tested in this row or else NA |
| IGLV | IGL V gene, if tested in this row or else NA |
| IGLJ | IGL J gene, if tested in this row or else NA |
| IGHV_IGHJ | IGH V and J genes, if tested in this row or else NA |
| IGKV_IGKJ | IGK V and J genes, if tested in this row or else NA |
| IGLV_IGLJ | IGL V and J genes, if tested in this row or else NA |
| parameter | Name of the parameter from the linear model |
| coefficient | Coefficient for the parameter |
| se | Standard Error for the coefficient |
| ci | Confidence interval for the coefficient |
| ci_low | Lower bound of the confidence interval |
| ci_high | Upper bound of the confidence interval |
| t | t-statistic for the coefficient |
| df_error | Degrees of freedom for the linear model |
| p | P-value for the coefficient |
| fdr | False Discovery Rate (adjusted p-value) |

**Table S9.**
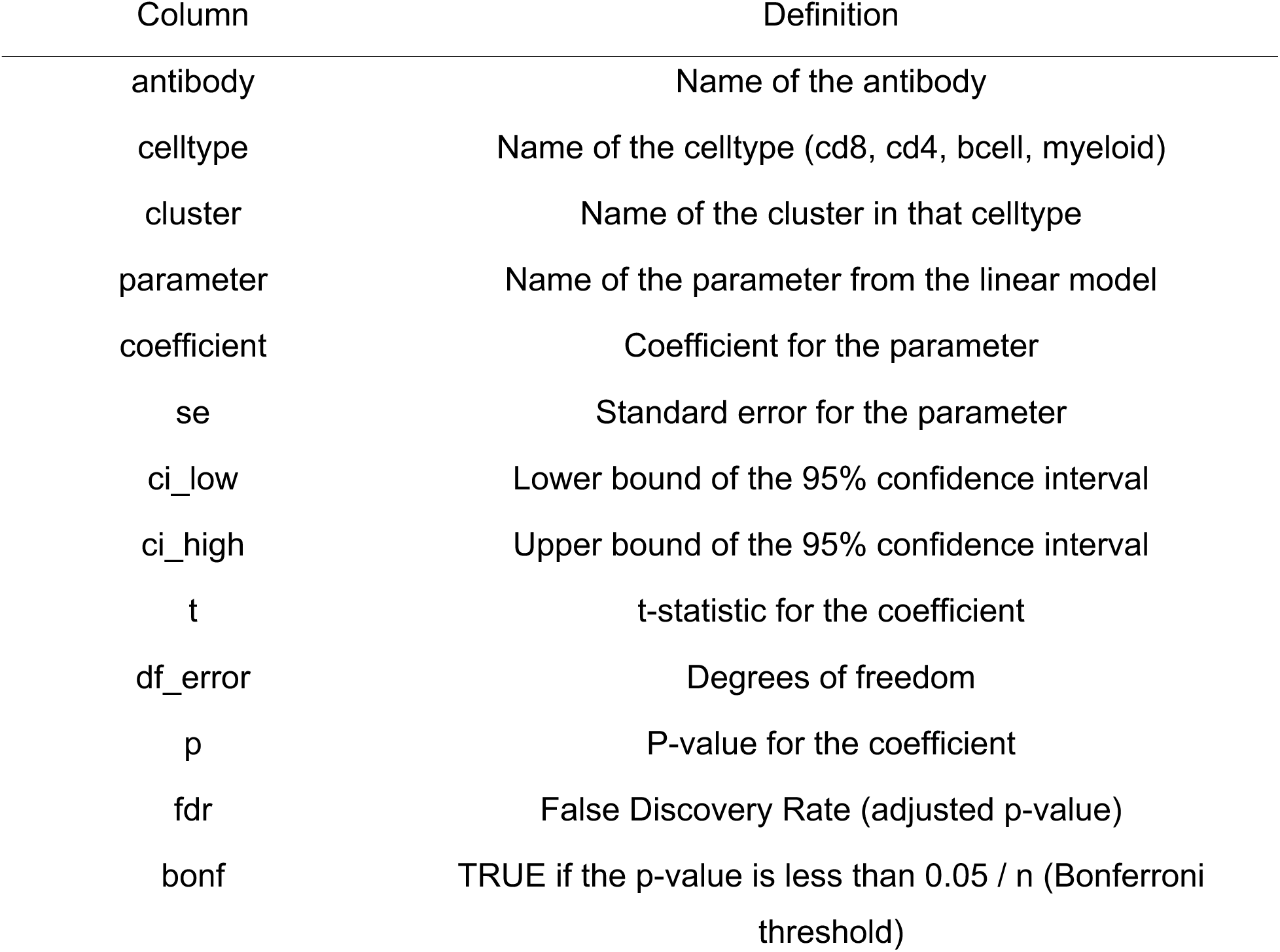
Antibody associations with cell clusters in the acute data. Association results for 75 antibodies with the abundance of all cell clusters from the acute scRNA-seq data.

| Column | Definition |
| --- | --- |
| antibody | Name of the antibody |
| celltype | Name of the celltype (cd8, cd4, bcell, myeloid) |
| cluster | Name of the cluster in that celltype |
| parameter | Name of the parameter from the linear model |
| coefficient | Coefficient for the parameter |
| se | Standard error for the parameter |
| ci_low | Lower bound of the 95% confidence interval |
| ci_high | Upper bound of the 95% confidence interval |
| t | t-statistic for the coefficient |
| df_error | Degrees of freedom |
| p | P-value for the coefficient |
| fdr | False Discovery Rate (adjusted p-value) |
| bonf | TRUE if the p-value is less than 0.05 / n (Bonferroni threshold) |

**Table S10.** Gene set enrichment results for the acute data. Association results for 6605 gene sets with severity, age, infection, sex, and time.

| Column | Definition |
| --- | --- |
| celltype | acute cell type (myeloid, cd4, cd8, bcell) |
| cluster | cluster within the cell type, or all |
| coef | name of the linear model coefficient that is tested with fgsea |
| geneset | name of the gene set collection (e.g., MSIGDB_HALLMARK) |
| pathway | name of the pathway in the gene set collection |
| pval | p-value from fgsea |
| padj | adjusted p-value (FDR) |
| log2err | log2 error from fgsea |
| ES | enrichment score from fgsea |
| NES | normalized enrichment score from gsea |
| size | number of genes in this pathway |
| leadingEdge | the top genes with the strongest log2FC in this pathway |

**Table S11.** Antibody association results for the acute data. Association results for 75 antibodies with severity, age, infection, sex, and time.

| Column | Definition |
| --- | --- |
| name | Name of the protein |
| parameter | Name of the parameter from the linear model |
| coefficient | Coefficient for the parameter |
| se | Standard error for the parameter |
| ci_low | Lower bound of the 95% confidence interval |
| ci_high | Upper bound of the 95% confidence interval |
| t | t-statistic for the coefficient |
| df_error | Degrees of freedom |
| p | P-value for the coefficient |
| fdr | False Discovery Rate (adjusted p-value) |
| bonf | TRUE if the p-value is less than 0.05 / n (Bonferroni threshold) |

**Table S12.**
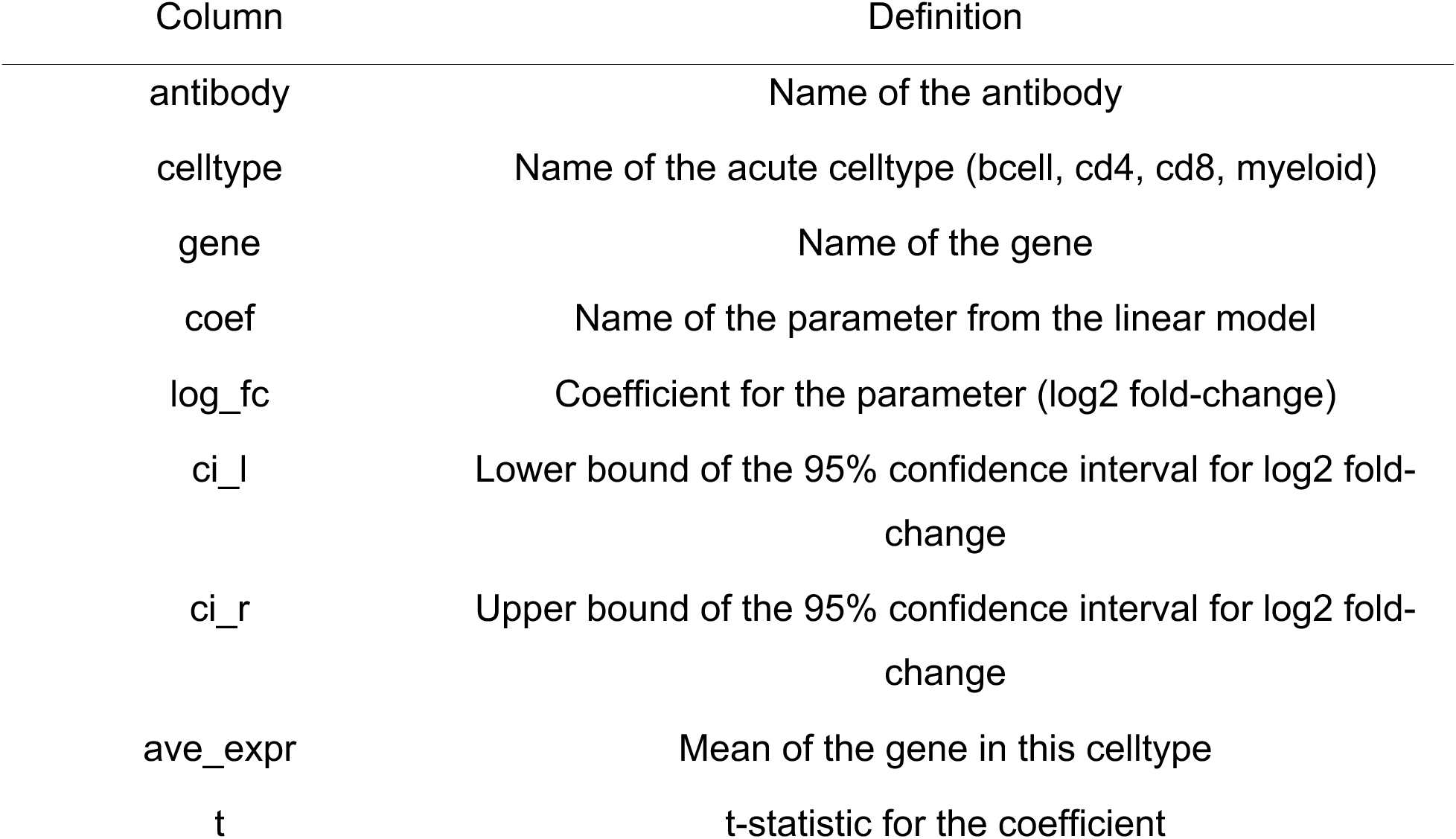
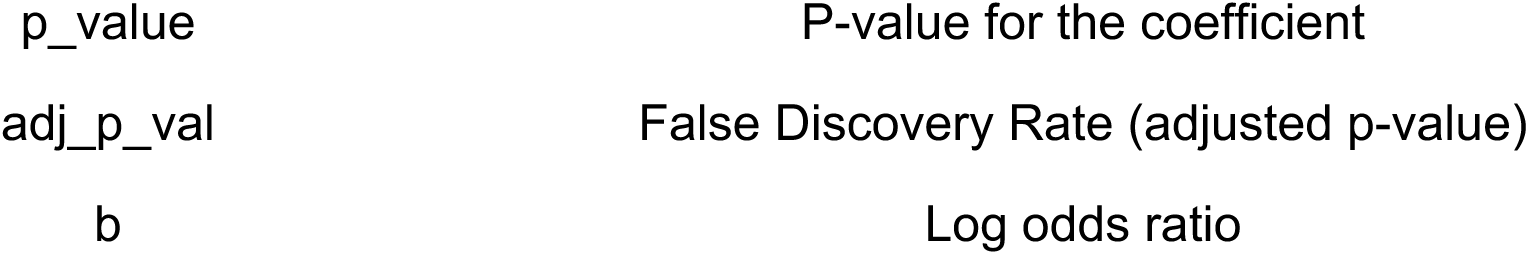
Antibody associations with gene expression in the acute data. A table (35,742,420 rows, 12 columns) with gene expression statistics for a linear model fit to each gene and each of the 75 antibodies in the acute data. We provide results for gene expression at the level of cell lineages (cd4, cd8, bcell, myeloid).

**Table S13.** Public TCR sequences observed in the acute data. TCR sequences that are observed in 2 or more acute donors. Organized into two spreadsheets (cd8, cd4).

| Column | Definition |
| --- | --- |
| public_id | Donor ID |
| time_point | Time point after entering the hospital in days |
| acuity_max_cat | The maximum acuity over all time points |
| age_cat | Age category |
| covid | COVID status (tested positive prior to enrollment or during hospitalization) |
| n_donors | Number of donors that have this TCR |
| n | Number of cells that have this TCR, per sample (donor_time) |
| rank | Rank of the abundance of this TCR, per sample (donor_time) |
| percent | This TCR accounts for what percent of TCRs from this sample |
| tcr_id | Concatenation of alpha and beta V, J, and CDR3 |
| TRBV | Beta V gene |
| TRAV | Alpha V gene |
| TRBJ | Beta J gene |
| TRAJ | Alpha J gene |
| TRB_cdr3 | Beta CDR3 amino acid sequence |
| TRA_cdr3 | Alpha CDR3 amino acid sequence |

**Table S14.** TCR association results for the acute data. Association results for each TCR with acuity in CD8 T cells and CD4 T cells.

| Column | Definition |
| --- | --- |
| celltype | Cell lineage (either CD4 or CD8 T cell) |
| test | Which gene (or genes) are tested in this row |
| TRAV | Alpha V gene, if tested in this row or else NA |
| TRAJ | Alpha J gene, if tested in this row or else NA |
| TRBV | Beta V gene, if tested in this row or else NA |
| TRBJ | Beta J gene, if tested in this row or else NA |
| TRAV_TRAJ | Alpha V and J genes, if tested in this row or else NA |
| TRBV_TRBJ | Beta V and J genes, if tested in this row or else NA |
| parameter | Name of the parameter from the linear model |
| coefficient | Coefficient for the parameter |
| se | Standard Error for the coefficient |
| ci | Confidence interval for the coefficient |
| ci_low | Lower bound of the confidence interval |
| ci_high | Upper bound of the confidence interval |
| t | t-statistic for the coefficient |
| df_error | Degrees of freedom for the linear model |
| p | P-value for the coefficient |
| fdr | False Discovery Rate (adjusted p-value) |

**Table S15.** HLA amino acid positions associated with COVID acuity. Association results for 348 amino acid positions across 7 HLA genes (A, B, C, DPA1, DPB1, DQA1, DQB1) with COVID-19 acuity.

| Column | Definition |
| --- | --- |
| gene | Name of the HLA gene |
| position | Name of the amino acid position in the HLA gene |
| param | Name of the parameter from the linear model |
| coefficient | Coefficient for the parameter |
| se | Standard error for the parameter |
| ci_low | Lower bound of the 95% confidence interval |
| ci_high | Upper bound of the 95% confidence interval |
| t | t-statistic for the coefficient |
| df_error | Degrees of freedom |
| p | P-value for the coefficient |
| fdr | False Discovery Rate (adjusted p-value) |

**Table S16.**
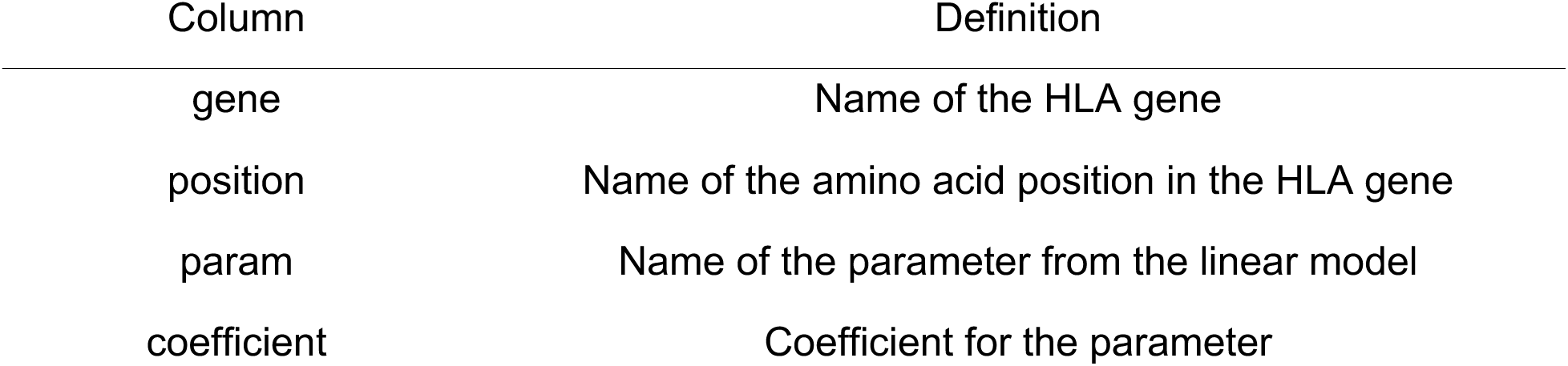
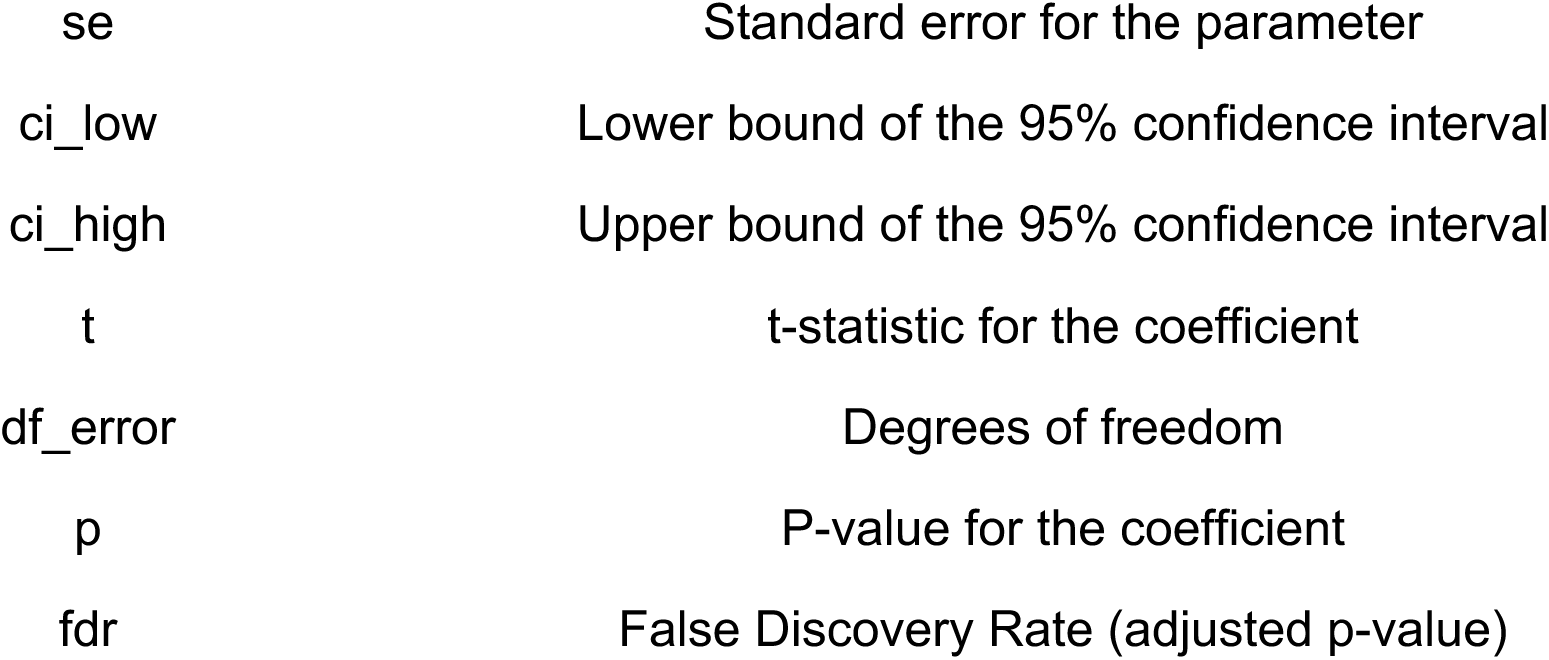
HLA amino acid positions associated with SARS-CoV-2 viral load. Association results for 381 amino acid positions across 7 HLA genes (A, B, C, DPA1, DPB1, DQA1, DQB1) with the SARS-CoV-2 viral load in each sample.

| Column | Definition |
| --- | --- |
| gene | Name of the HLA gene |
| position | Name of the amino acid position in the HLA gene |
| param | Name of the parameter from the linear model |
| coefficient | Coefficient for the parameter |
| se | Standard error for the parameter |
| ci_low | Lower bound of the 95% confidence interval |
| ci_high | Upper bound of the 95% confidence interval |
| t | t-statistic for the coefficient |
| df_error | Degrees of freedom |
| p | P-value for the coefficient |
| fdr | False Discovery Rate (adjusted p-value) |

**Table S17.**
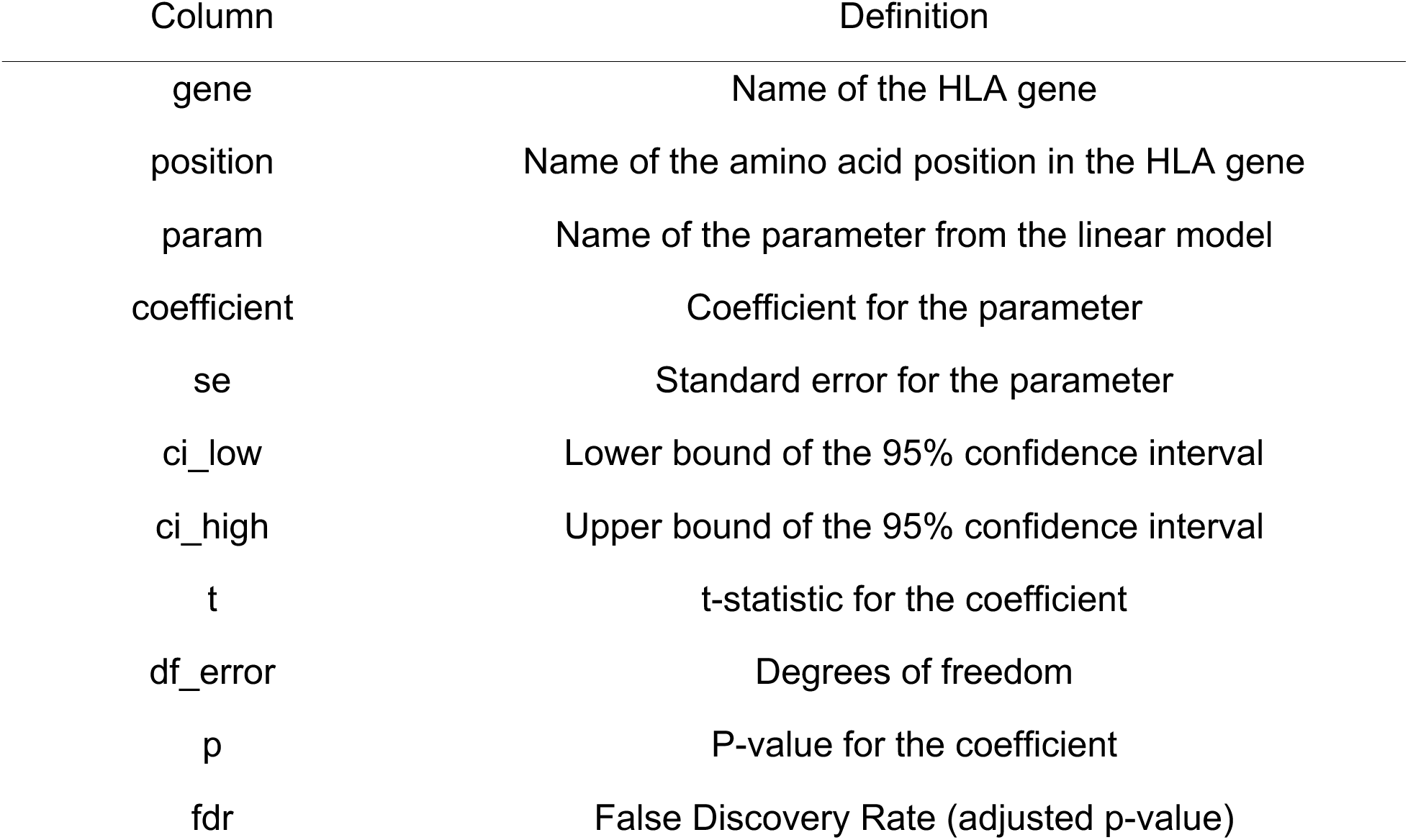
HLA amino acid positions associated with SARS-CoV-2 neutralization. Association results for 381 amino acid positions across 7 HLA genes (A, B, C, DPA1, DPB1, DQA1, DQB1) with an *in vitro* SARS-CoV-2 neutralization assay (see Methods).

**Table S18.** HLA amino acid positions associated with CD8 T cell TRBV genes. Association results for 530 amino acid positions across 13 HLA genes (A, B, C, DMA, DMB, DOB, DPA1, DPB1, DQA1, DQB1, DRA, E, F) and the abundance of CD8 T cells with different TRBV genes.

| Column | Definition |
| --- | --- |
| celltype | Name of the acute celltype (cd8) for which we run the analysis |
| TRBV | Name of the TRBV gene |
| gene | Name of the HLA gene |
| position | Name of the amino acid position in the HLA gene |
| param | Name of the parameter from the linear model |
| coefficient | Coefficient for the parameter |
| se | Standard error for the parameter |
| ci_low | Lower bound of the 95% confidence interval |
| ci_high | Upper bound of the 95% confidence interval |
| t | t-statistic for the coefficient |
| df_error | Degrees of freedom |
| p | P-value for the coefficient |
| fdr | False Discovery Rate (adjusted p-value) |

**Table S19.** Blood protein association with cell clusters in the acute data Association results for 1,463 circulating blood proteins with the abundance of all cell clusters from the acute scRNA-seq data.

| Column | Definition |
| --- | --- |
| protein | Name of the protein |
| celltype | Name of the celltype (cd8, cd4, bcell, myeloid) |
| cluster | Name of the cluster in that celltype |
| parameter | Name of the parameter from the linear model |
| coefficient | Coefficient for the parameter |
| se | Standard error for the parameter |
| ci_low | Lower bound of the 95% confidence interval |
| ci_high | Upper bound of the 95% confidence interval |
| t | t-statistic for the coefficient |
| df_error | Degrees of freedom |
| p | P-value for the coefficient |
| fdr | False Discovery Rate (adjusted p-value) |
| bonf | TRUE if the p-value is less than 0.05 / n (Bonferroni threshold) |

**Table S20.**
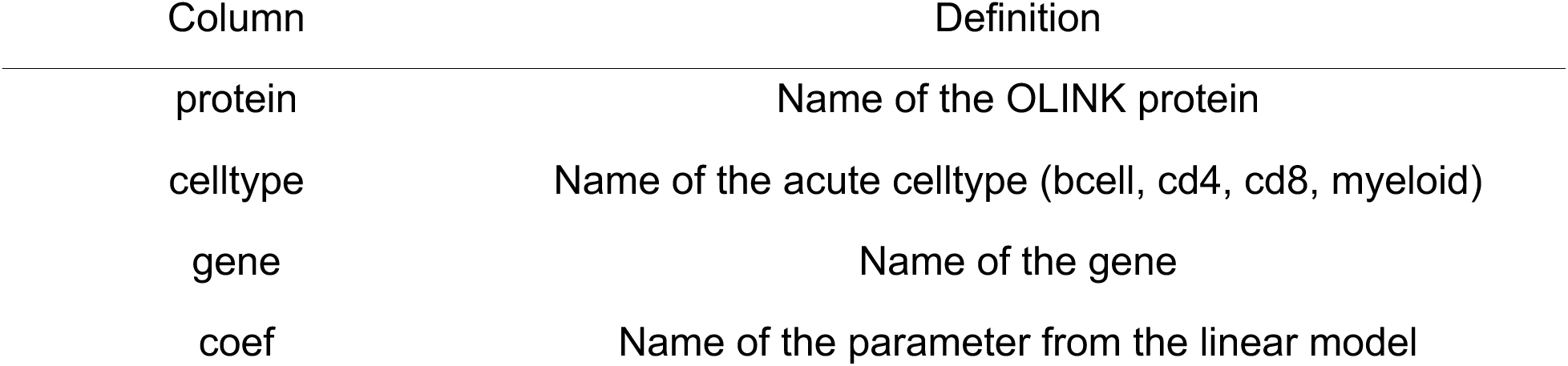
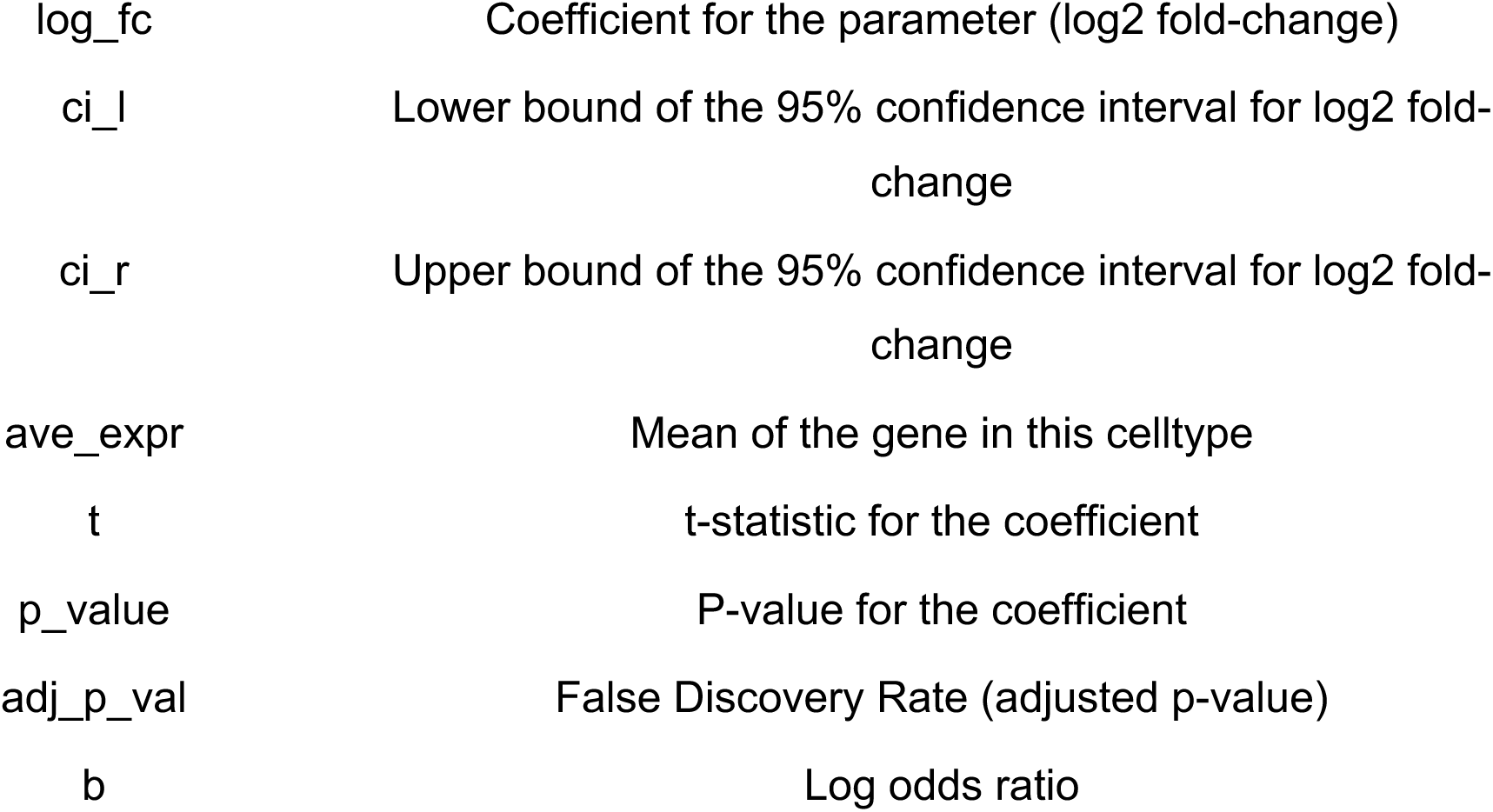
Blood protein association with gene expression in the acute data. A table (687,541,239 rows, 12 columns) with gene expression statistics for a linear model fit to each gene and each of the 1,463 circulating blood protein in the acute data. We provide results for gene expression at the level of cell lineages (cd4, cd8, bcell, myeloid).

**Table S21.** Blood protein association with acuity in the acute data. This includes association results for 1,463 circulating blood proteins with acuity.

| Column | Definition |
| --- | --- |
| name | Name of the protein |
| parameter | Name of the parameter from the linear model |
| coefficient | Coefficient for the parameter |
| ci_low | Lower bound of the 95% confidence interval |
| ci_high | Upper bound of the 95% confidence interval |
| AveExpr | Mean of the protein value |
| t | t-statistic for the coefficient |
| p | P-value for the coefficient |
| fdr | False Discovery Rate (adjusted p-value) |
| B | Log odds ratio for this protein and this variable |

**Table S22.**
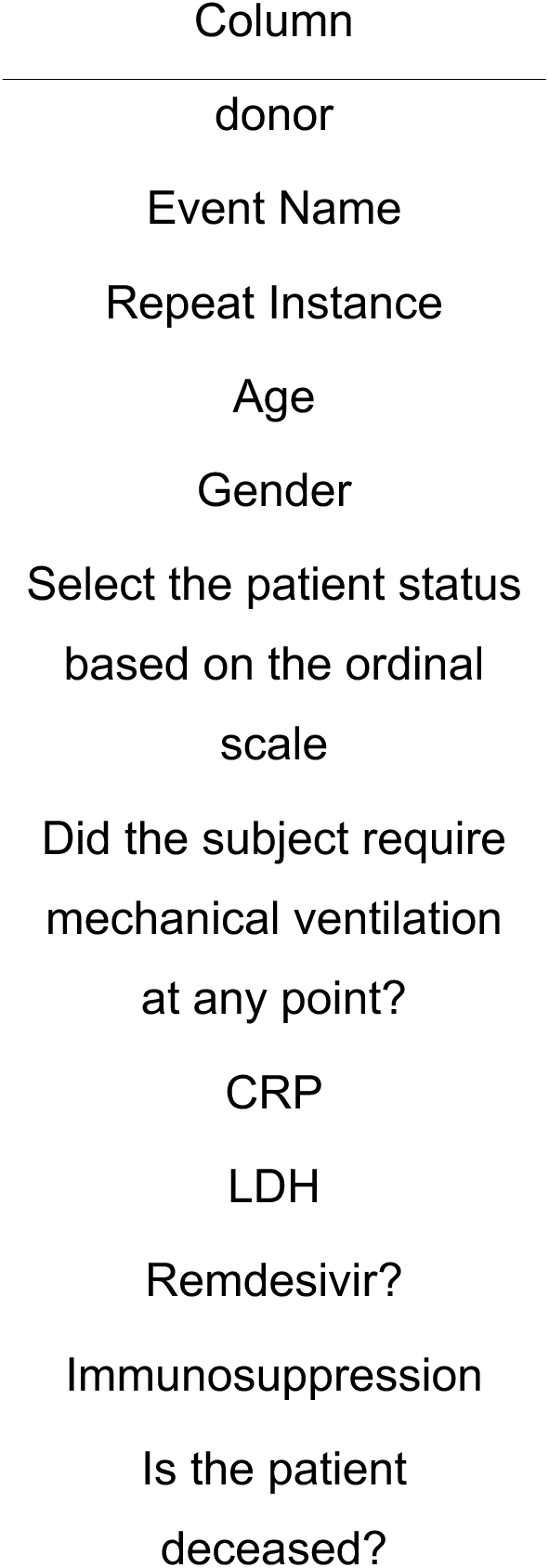
Clinical patient metadata for the tocilizumab data. Clinical patient metadata for 62 patients.

**Table S23.**
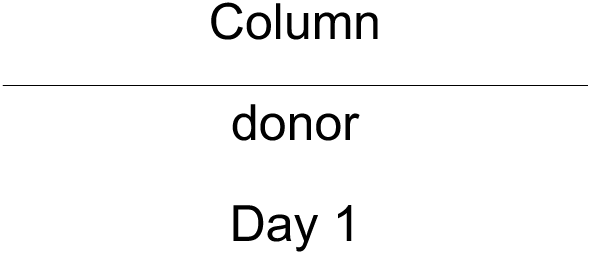
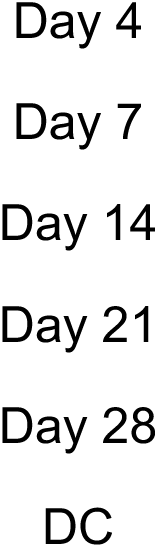
Patient IL-6 levels over time for the tocilizumab data. IL-6 measurements for 26 patients and 7 time points.

**Table S24.** Differential gene expression statistics for the tocilizumab data. A table with gene expression statistics for one linear model fit to each gene in the tocilizumab data. We provide results for cell lineages (cd4, cd8, bcell, myeloid) as well as cell clusters within each lineage.

| Column | Definition |
| --- | --- |
| celltype | Name of the toci dataset cell lineage (cd4, cd8, bcell, myeloid) |
| cluster | Number of the cell cluster in the cell lineage |
| coef | Name of the parameter from the linear model |
| gene | Name of the gene |
| ave_expr | Average log2CPM expression of the gene |
| quantile_expr | Quantile of the average expression |
| p_value | P-value for the coefficient |
| adj_p_val | False Discovery Rate (adjusted p-value) |
| log_fc | Coefficient for the parameter (log2 Fold-Change) |
| ci_l | Lower bound of the 95% confidence interval for the log2 FC |
| ci_r | Upper bound of the 95% confidence interval for the log2 FC |
| t | t-statistic for the coefficient |
| b | Log odds |
| ensembl_id | Ensembl ID for the gene |
| percent_cells | Percent of cells expressing the gene |

**Table S25.** Gene set enrichment results for the tocilizumab data. Association results for 2406 gene sets with tocilizumab treatment.

| Column | Definition |
| --- | --- |
| celltype | cell type (mnp, cd4, cd8, bcell) |
| cluster | cluster within the cell type, or all |
| coef | name of the linear model coefficient that is tested with fgsea |
| geneset | name of the gene set collection (e.g., MSIGDB_HALLMARK) |
| pathway | name of the pathway in the gene set collection |
| pval | p-value from fgsea |
| padj | adjusted p-value (FDR) |
| log2err | log2 error from fgsea |
| ES | enrichment score from fgsea |
| NES | normalized enrichment score from gsea |
| size | number of genes in this pathway |
| leadingEdge | the top genes with the strongest log2FC in this pathway |

**Table S26.** Differential gene expression statistics for the convalescent data. A table with gene expression statistics for one linear model fit to each gene in the convalescent data. We provide results for cell lineages (cd4, cd8, bcell, myeloid) as well as cell clusters within each lineage.

| Column | Definition |
| --- | --- |
| celltype | Name of the conva dataset cell lineage (cd4, cd8, bcell, myeloid) |
| cluster | Number of the cell cluster in the cell lineage |
| coef | Name of the parameter from the linear model |
| gene | Name of the gene |
| ave_expr | Average log2CPM expression of the gene |
| quantile_expr | Quantile of the average expression |
| p_value | P-value for the coefficient |
| adj_p_val | False Discovery Rate (adjusted p-value) |
| log_fc | Coefficient for the parameter (log2 Fold-Change) |
| ci_l | Lower bound of the 95% confidence interval for the log2 FC |
| ci_r | Upper bound of the 95% confidence interval for the log2 FC |
| t | t-statistic for the coefficient |
| b | Log odds |
| ensembl_id | Ensembl ID for the gene |
| percent_of_cells | Percent of cells expressing the gene |
| rank | Rank of the gene, based on p-value and log2FC |

**Table S27.**
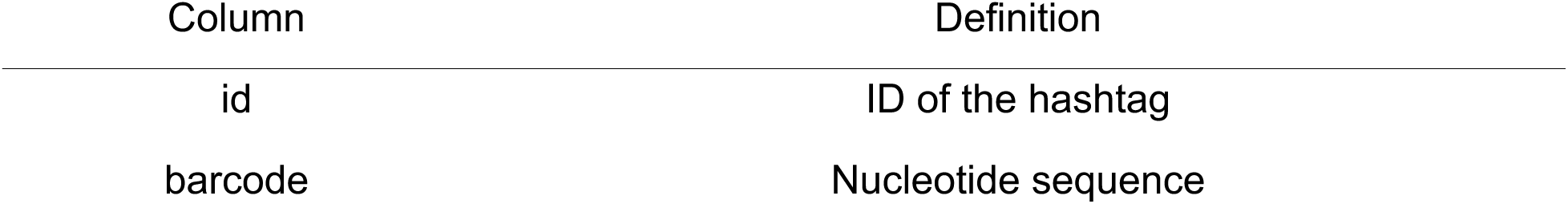
Hashing ids and nucleotide sequences. Names and barcode sequences for 10 hashtags.

**Table S28.** CITE-seq target proteins and barcode sequences. Names and barcode sequences for 197 CITE-seq features.

| Column | Definition |
| --- | --- |
| id | ID of the CITE-seq feature |
| antibody_description | Description of the CITE-seq feature |
| protein_target | Name of the protein target |
| barcode | Barcode sequence for the CITE-seq feature |
| clone | Name of the clone used |

**Table S29.** Multiplexed antigen array content for antibody data. Names, vendors, and catalog numbers for 77 antibodies.

| Column | Definition |
| --- | --- |
| Bead ID | Identifier of the bead for this antigen. |
| Class | General description of this group of antigens. |
| Antigen | Name of the specific antigen. |
| Vendor | Vendor who supplies the antibody for this antigen. |
| Catalog Number | Catalog number for ordering this antibody. |

